# Somatic mitochondrial DNA mutations are a source of heterogeneity among primary leukemic cells

**DOI:** 10.1101/2024.09.26.24314381

**Authors:** Kelly McCastlain, Catherine Welsh, Yonghui Ni, Liang Ding, Melissa Franco, Robert J. Autry, Besian I. Sejdiu, Ti-Cheng Chang, Wenan Chen, Huiyun Wu, Qingfei Pan, Veronica Gonzalez-Pena, Patrick Schreiner, Sasi Arunachalam, Joung Hyuck Joo, Benshang Li, Shuhong Shen, Samuel Brady, Jinghui Zhang, Charles Gawad, William E. Evans, Madan Babu, Konstantin Khrapko, Gang Wu, Jiyang Yu, Stanley Pounds, Mondira Kundu

## Abstract

Somatic mitochondrial DNA (mtDNA) mutations are prevalent in tumors, yet defining their biological significance remains challenging due to the intricate interplay between selective pressure, heteroplasmy, and cell state. Utilizing bulk whole-genome sequencing data from matched tumor and normal samples from two cohorts of pediatric cancer patients, we uncover differences in the accumulation of synonymous and nonsynonymous mtDNA mutations in pediatric leukemias, indicating distinct selective pressures. By integrating single-cell sequencing (SCS) with mathematical modeling and network-based systems biology approaches, we identify a correlation between the extent of cell-state changes associated with tumor-enriched mtDNA mutations and the selective pressures shaping their distribution among individual leukemic cells. Our findings also reveal an association between specific heteroplasmic mtDNA mutations and cellular responses that may contribute to functional heterogeneity among leukemic cells and influence their fitness. This study highlights the potential of SCS strategies for distinguishing between pathogenic and passenger somatic mtDNA mutations in cancer.

**Graphical Abstract:** 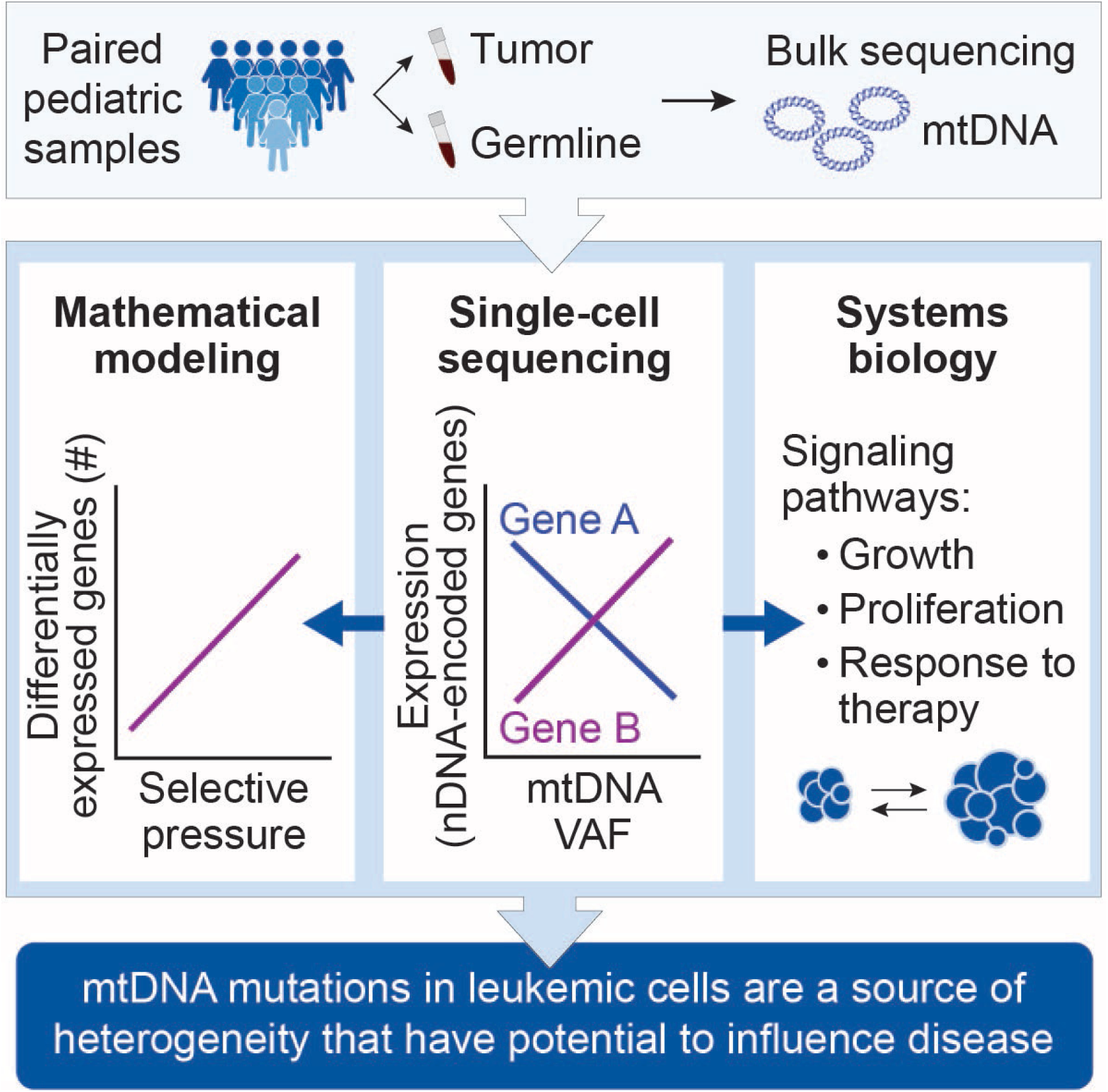

**Highlights:**

- Nonsynonymous and synonymous mtDNA SNVs are subject to different selective pressures among pediatric leukemias
- The distribution of mtDNA SNV VAFs among individual leukemic cells can provide evidence for selective pressure
- The impact of mtDNA SNVs on gene expression is correlated with selective pressure
- Somatic mtDNA SNVs associated with cell state changes are a source of heterogeneity among leukemic cells

## Introduction

The mitochondrial genome, which encodes key subunits of the electron transport chain (ETC) and the requisite RNA machinery for their translation, is critical for supporting oxidative phosphorylation (OXPHOS) and producing tumor-modifying metabolic signals [e.g., ATP, reactive oxygen species (ROS), NAD^+^, and FAD]^1–3^. OXPHOS and the tricarboxylic (TCA) cycle are also common targets of metabolic reprogramming driven by oncogenic mutations in nuclear DNA-encoded genes, including those that dysregulate MYC and/or MTOR signaling^4,5^. Given the intricate interplay between OXPHOS and the TCA cycle^6^, mutations in mitochondrial DNA (mtDNA) have the potential to enhance or impede tumorigenesis by influencing metabolic signaling and the availability of metabolite pools^3,7,8^. Nevertheless, although somatic mtDNA mutations have been reported in approximately 46% of pediatric^9^ and 60% of adult^10^ malignancies, investigating their contribution to tumor pathophysiology has been difficult.

Until recently, the mitochondrial genome has been resistant to modification by editing technologies^11,12^. Therefore, insights about the pathogenicity of mtDNA mutations have been limited to what can be inferred from primary mitochondrial diseases^13–21^ and modeling specific kindred^22–24^ or disease-associated mutations^25–27^ via cytoplasmic hybrid (cybrid) models. Cybrid cell lines are produced by fusing an enucleated cell (the mitochondrial donor) with a Rho0 cell (the nuclear donor), which is devoid of mtDNA ^28^. When all copies of mtDNA within cybrid cells harbor the same mutation, a state known as homoplasmy, the effects of clinically relevant mutations are often mediated by perturbations in respiratory capacity and ROS production, although the degree and direction of these changes are variant-specific^7,22,23,25,27^. However, the overlap between tumor-enriched (TU) mtDNA mutations and those for which cybrid models exist (e.g., maternally inherited mitochondrial disorders) is limited; therefore, most TU mtDNA mutations have not been modeled, and their functional consequences remain unknown.

The existence of multiple copies of mtDNA within a cell further complicates modeling efforts, as the ability of a given mtDNA mutation to influence cellular processes depends on the level of heteroplasmy, or the proportion of mutated mtDNA alleles present within a cell (i.e., variant allele fraction, or VAF). For instance, the clinically relevant m.3243A>G mutation in *MT-TL1*, a gene that encodes a mitochondrial transfer RNA (tRNA), is associated with various disease phenotypes that depend on the variant’s heteroplasmic range^13,29–31^. Notably, in a cybrid series harboring increasing heteroplasmic levels of the m.3243A>G allele, impaired OXPHOS was detected in cells with >50% mutated mtDNA, but altered levels of TCA cycle metabolites and concomitant changes in transcriptional programs and epigenomic markers were detected in cells with only 20%-30% mutated mtDNA^24,32^.

Given the paucity of appropriate model systems, it is not surprising that the functional consequences of TU mtDNA mutations––whether they confer a selective advantage that promotes oncogenesis (positive selection) or impede growth and are selected against (purifying or negative selection) ––remain controversial. Initial investigations of adult solid malignancies suggested that the accumulation of homoplasmic, tumor-specific mtDNA mutations is a consequence of positive selection^23,33–36^; however, with few exceptions^23,34^, those studies were associative and did not consider the functional significance of variants. More recently, bulk sequencing data from matched tumor and normal tissue samples in large-scale surveys have been used to comprehensively describe the mtDNA mutational landscape across tumor subtypes^9,10,37–41^. Although these studies reported increased numbers of missense mtDNA mutations among tumors, resolving which selective pressure is operative has been challenging. Differences in cohort composition, analytical approaches, and the extent of functional characterization of nonsynonymous mutations have complicated efforts to distinguish whether TU mtDNA mutations are subjected to purifying (negative) ^10,37,40,41^ or positive^9,37–39^ selection, or if they arise due to relaxation of purifying selection^38,41^ during development of the tumor.

In most adult tumors, deleterious truncating mtDNA mutations typically occur at lower VAFs than do missense or silent mutations^10,41^. This finding, combined with increasing evidence that tumors rely on OXPHOS for their growth^7,42–44^, has led to the notion that the propagation of deleterious mtDNA mutations is constrained by negative selection pressures. Although these observations appear to support the argument against the functional importance of mtDNA mutations in cancer, bulk DNA-sequencing strategies cannot determine heteroplasmy levels in individual tumor cells––information that is required to corroborate the effects of selective pressure on variant accrual and assess the impact of mtDNA mutations on cellular functions.

Single-cell sequencing studies have suggested the utility of heteroplasmic mtDNA variants as markers for identifying and tracking clonal populations^45–47^. However, single-cell sequencing technologies have not been systematically deployed to answer fundamental questions about the heteroplasmy levels and functional impact of pathogenic variants in tumor cells. Here, we present a multidimensional sequencing-based approach that circumvents technological barriers that preclude modeling of somatic mtDNA mutations at physiologically relevant levels of heteroplasmy. We then apply this strategy to address questions about selective pressure and the level and functionality of somatic mtDNA mutations in pediatric leukemia cells.

## Results

### Classification of mtDNA variants

We analyzed whole-genome sequencing (WGS) data from 637 paired pediatric tumor and germline (i.e., “normal” tumor-free tissue) samples sequenced under the auspices of the St. Jude/Washington University Pediatric Cancer Genome Project (PCGP)^48^. These tumors included 26 pediatric cancer subtypes comprising hematological (blood, 54%), central nervous system (brain, 19%) and solid tissue (solid, 27%) malignancies^48^ (**Table 1**). Samples were sequenced to an average depth of ∼30× for nuclear DNA–encoded genes, leading to a depth of ∼7,000× on the mitochondrial genome. Average sequencing coverage was then leveraged to estimate the mitochondrial DNA copy number (mtDNAcn) across all samples (**Supplementary Note 1, Table S1A**)^49^. To identify and classify mtDNA variants in pediatric samples, we mapped reads to the mitochondrial genome by using a stringent computational pipeline with a conservative mutation-calling threshold (**Figure S1A**), as was used for adult samples from The Cancer Genome Atlas (TCGA)^10^. This strategy minimized the risk of potential false-positive calls associated with nuclear sequences of mitochondrial origin (i.e., nuclear mtDNA sequences)^50^ (**Figure S1B**).

**Table 1.** PCGP tumor subtypes categorized by tissue type (blood, brain, or solid) and the number of subtypes sequenced per platform. **Abbreviations:** ACT, adrenocortical carcinoma; ALL, acute lymphoblastic leukemia; AML, acute myelogenous leukemia; AMLM7, acute megakaryoblast leukemia; CBF, core-binding factor AML; CPC, choroid plexus carcinoma; E2A, E2A–PBX1 B-lineage ALL; EPD, ependymoma; DUX4r, *DUX4*-rearranged B-lineage ALL; ETV, *ETV6–RUNX1* B-lineage ALL; EWS, Ewing sarcoma; HGG, high-grade glioma; HYPER, hyperdiploid B-lineage ALL; HYPO, hypodiploid B-lineage ALL; iAMP21, intrachromosomal amplification of chromosome 21 B-lineage ALL; INF, infant B-linage ALL; LGG, low-grade glioma; MB, medulloblastoma; MEL, melanoma; OS, osteosarcoma; mt, mitochondria; PH, Philadelphia chromosome B-lineage ALL; PHL, Philadelphia chromosome–like B-lineage ALL; PHL-Hyp, Philadelphia chromosome–like B-lineage ALL with hyperdiploid/hypodiploid features; RB retinoblastoma; RHB, rhabdomyosarcoma; RNAseq, RNA sequencing; TALL, T-lineage ALL; TCF3-HLF, *TCF3–HLF* B-lineage ALL; WGS, whole genome sequencing.

| Tumor Subtype | Category | WGS (n) | RNAseq (n) | Average mt reads (tumor WGS) | Average mt reads (germline WGS) | Average mt reads (RNA) |
| --- | --- | --- | --- | --- | --- | --- |
| ACT | Solid | 20 | 20 | 24566.76 | 3639.47 | 14465.81 |
| AML M7 | Blood | 4 | 4 | 5540.16 | 4462.12 | 10258.25 |
| CBF | Blood | 10 | 5 | 5935.20 | 3107.60 | 17260.78 |
| CPC | Brain | 4 | 1 | 22881.74 | 2268.52 | 9731.86 |
| DUX4r | Blood | 31 | 30 | 11344.13 | 3381.44 | 9995.29 |
| E2A | Blood | 30 | 8 | 7203.50 | 3229.68 | 9767.87 |
| EPD | Brain | 40 | 34 | 14521.80 | 3631.97 | 11141.20 |
| ETV | Blood | 50 | 49 | 6609.37 | 4040.52 | 8240.67 |
| EWS | Solid | 35 | 0 | 10458.44 | 3044.47 | - |
| HGG | Brain | 35 | 28 | 16934.64 | 9275.73 | 8392.90 |
| HYPER | Blood | 43 | 0 | 9025.76 | 2636.55 | - |
| HYPO | Blood | 19 | 0 | 8231.88 | 1883.20 | - |
| iAMP21 | Blood | 12 | 8 | 10177.23 | 3296.96 | 9431.98 |
| INF | Blood | 15 | 14 | 4077.39 | 3059.45 | 7159.17 |
| LGG | Brain | 38 | 27 | 20435.65 | 3485.47 | 14395.50 |
| MB | Brain | 22 | 3 | 6129.65 | 9511.11 | 8548.56 |
| MEL | Solid | 5 | 4 | 14325.60 | 3662.46 | 10883.70 |
| NBL | Solid | 55 | 0 | 5958.33 | 5748.98 | - |
| OS | Solid | 40 | 20 | 9335.58 | 3820.25 | 9677.88 |
| PH | Blood | 36 | 24 | 8405.07 | 3415.88 | 9882.28 |
| PHL | Blood | 23 | 14 | 7906.30 | 2431.99 | 10331.75 |
| PHL-Hyp | Blood | 10 | 2 | 9027.12 | 2975.16 | 8772.05 |
| RB | Solid | 11 | 1 | 8391.68 | 2708.87 | 5541.89 |
| RHB | Solid | 9 | 9 | 8033.02 | 3393.88 | 8273.89 |
| TALL | Blood | 14 | 2 | 5055.86 | 2711.23 | 6368.03 |
| TCF3-HLF | Blood | 5 | 0 | 8620.42 | 3628.37 | - |

Among the 616 paired samples that passed all quality-control (QC) thresholds, we identified 18,372 mtDNA single-nucleotide variants (SNVs) (**Table S1B**). These were sorted into the following four SNV classes based on the VAF in the tumor and germline samples (**Figures 1A** and **S1C**): (1) stably inherited (INH) mtDNA SNVs; (2) TU mtDNA variants, which were present in 38.1% of the samples in our cohort (**Supplementary Note 2**); (3) mtDNA SNVs that were heteroplasmic in the germline samples (GH); and (4) heteroplasmic variants that were shared (SH) between the tumor and the germline samples. The *de novo* mtDNA mutations (TU, GH, and SH) represent three classes of SNVs acquired during development or transformation^51^ that can be compared.

**Figure 1.**
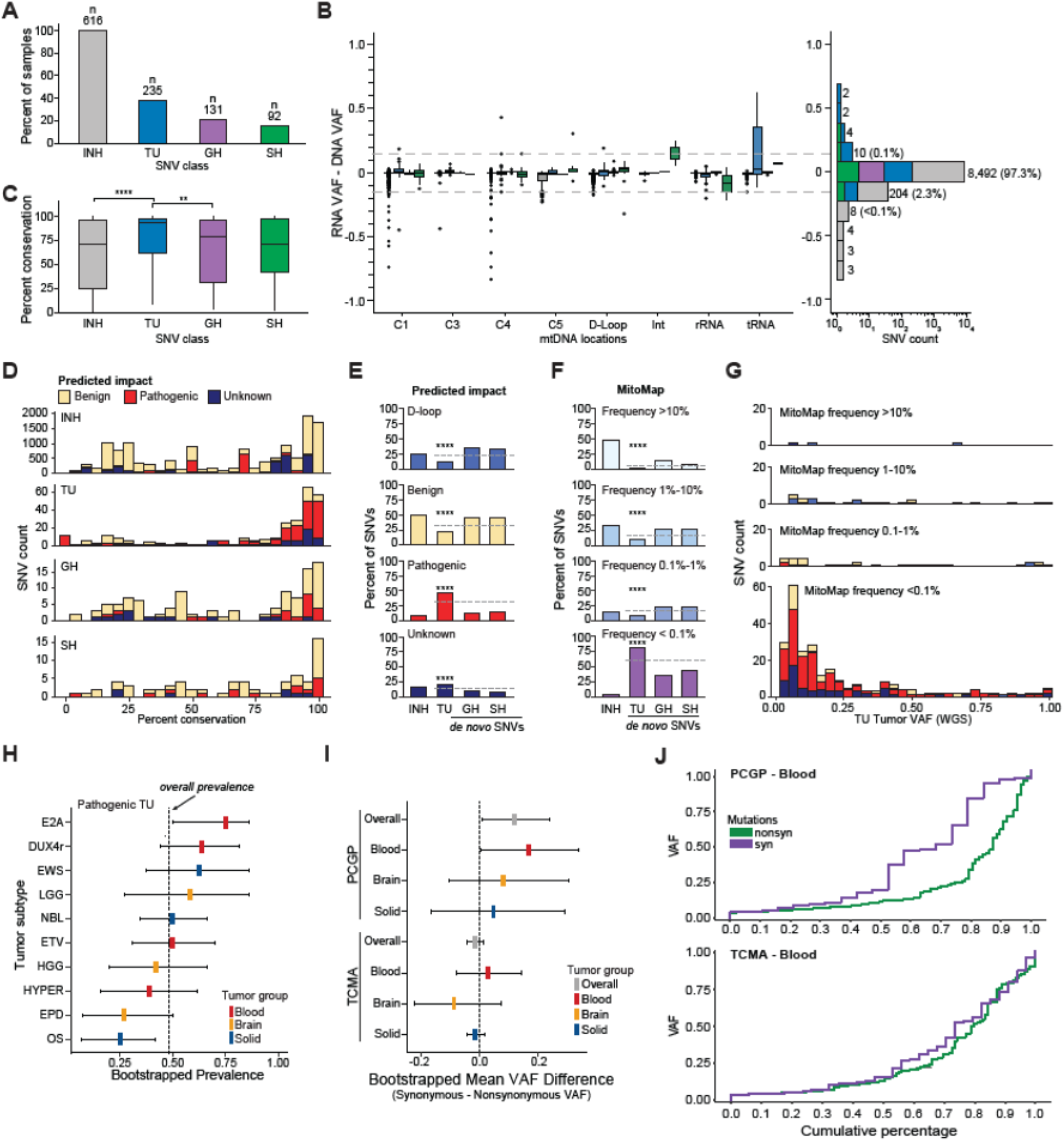
Tumor-enriched mtDNA variants exist at low levels of heteroplasmy and are more likely than other *de novo* mtDNA variants to be rare among the general population and of functional consequence. **(A)** Total number and percentage of samples carrying mtDNA variants in inherited or somatic SNV classes. SNV classifications are depicted by color and abbreviation. **(B)** RNAseq validation of 8,732 mitochondrial variants reveals a high degree of concordance between VAFs based on RNAseq (RNA VAF) and those based on WGS (DNA VAF). Left panel: Boxplots illustrating differences between RNA and DNA VAFs by gene groups. The gray dashed lines denote a 15% difference threshold. Right panel: Histogram showing mtDNA SNV counts binned by the difference between RNA and DNA VAFs. **(C)** Boxplot of mtDNA SNVs and percent evolutionary conservation (across all species) among inherited and somatic mitochondrial SNV classes. Total number of SNVs with conservation score, per class were INH, n = 13,274; TU, n = 277; GH, n = 110; SH, n = 69. Comparisons were made using two-sided Wilcoxon signed-rank tests. \*\**P* ≤0.01; \*\*\*\**P* ≤0.0001. **(D)** Stacked histograms integrating predicted functional impact (benign, pathogenic, or unknown) with percent evolutionary conservation and faceted by mtDNA SNV classification. Noncoding D-loop regions do not have conservation scores, thus are absent from the analysis. **(E)** Percentage of SNVs predicted by impact (D-loop/regulatory, benign, pathogenic, or unknown) among the SNV classes. *P*-values represent bootstrapped differences between observed and expected (dashed grey lines) prevalence for each predicted impact. The total number of SNVs per class were INH, n = 17,784; TU, n = 313; GH, n = 169; SH, n = 106. \*\*\*\**P* ≤0.0001. **(F)** Percentage of SNVs predicted by MitoMap^52^ population frequency, from common (pale blue) to extremely rare (purple) among the different SNV classes. *P*-values represent bootstrapped differences between the observed and expected (dashed grey lines) prevalence for each MitoMap frequency group. Total number of SNVs per class were INH, n = 17,784; TU, n = 313; GH, n = 169; SH, n = 106. \*\*\*\**P* ≤ 0.0001. **(G)** Stacked histograms integrating MitoMap frequency with WGS estimates of tumor VAF, revealing increased presence and clustering of rare mtDNA mutations that are predicted to be pathogenic at low allelic fractions. **(H)** Forest plot depicting the prevalence of TU SNVs predicted to be pathogenic among pediatric tumor subtypes with at least 30 samples. Confidence intervals (CI) are denoted by horizontal gray lines, and the overall prevalence across all subtypes is denoted by the vertical, dashed gray line. Bootstrapped *P*-values (OS, *P* = 0.0072; E2A, *P* = 0.0284). **(I)** Forest plot illustrating the mean difference in VAF between synonymous and nonsynonymous mutations. Overall mean VAF differences are depicted for each cohort and further categorized by tumor groups. The null value is depicted by a vertical, dashed gray line and CIs by horizontal gray lines. Bootstrapped mean difference with 95% CIs (PCGP overall, 0.119 (0.009, 0.236); PCGP Blood, 0.167 (0.006, 0.335)). **(J)** A comparison of cumulative VAF distributions between nonsynonymous (green) and synonymous (purple) mutations within the blood tumor groups of the PCGP (top panel) and TCMA (bottom panel) cohorts. Statistics presented in Figure 1I. **Abbreviations:** C1-5, complex I-V; DUX4r, *DUX4*-rearranged; E2A, *E2A–PBX1* translocation; EPD, ependymoma; ETV, *ETV6–RUNX1* translocation; EWS, Ewing sarcoma; GH, germline heteroplasmy; HGG, high-grade glioma; HYPER, hyperdiploid; INH, inherited germline; Int, intergenic; LGG, low-grade glioma; mtDNA, mitochondrial DNA; NBL, neuroblastoma; OS, osteosarcoma; PCGP, Pediatric Cancer Genome Project; rRNA, ribosomal RNA; SH, shared heteroplasmy; SNV, single nucleotide variant; TCMA, The Cancer Mitochondrial Atlas; tRNA, transfer RNA; TU, tumor-enriched; VAF, variant allele fraction; WGS, whole-genome sequencing.

We then examined the class-specific distributions of mtDNA SNVs across the mitochondrial genome and by major tumor groups (i.e., blood, brain, and solid) (**Supplementary Note 3**) and compared the mutational signatures among the SNV classes (**Supplementary Note 4**). These initial results were consistent with TU SNVs being under different selection pressures than the other SNV classes (**Supplementary Note 4**).

### RNAseq approximates mtDNA VAF calls from WGS

RNA sequencing (RNAseq) data were available for a subset of tumor samples that passed our QC thresholds (303/616) and were used to validate our mtDNA variant–calling pipeline. Using a sequencing depth criterion of ≥100 reads, we found that allelic RNA expression ratios (i.e., RNA VAFs) for mtDNA-encoded genes were highly concordant with their DNA analogs: only 0.7% (61/8,732) variants showed differences of >15% between RNA and DNA VAFs (**Figure 1B** and **Supplementary Note 5**). Among variants with discordant allelic fractions, 82% (50/61) had lower RNA VAFs and were predominantly INH variants (**Figure 1B** and **Supplementary Note 5**). Thus, the high concordance between WGS and RNAseq VAF estimates for *de novo* mtDNA variants validated our variant-calling approach.

### TU SNVs predicted to be pathogenic exist at low levels of heteroplasmy in bulk tumor samples

To assess the potential functional significance of *de novo* mtDNA variants, we first surveyed the evolutionary conservation scores of RNA- and protein-coding variants across all mitochondrial SNVs^52^ (**Figure 1C**). TU variants had significantly higher conservation scores (median, 93.3%) than the other SNV classes, including INH (median, 71.1%; *P* = 9.10 × 10^-12^, Wilcoxon rank sum test), GH (median, 78.9%; *P* = 0.004), and SH (median, 0.7110; *P* = 0.06), suggesting a greater predilection for pathogenic mutations among the TU SNVs^53^. When we integrated conservation scores with predicted functional impact, all four SNV classes exhibited peaks of highly conserved variants; however, only the TU SNV peak was enriched with variants predicted to be pathogenic (**Figure 1D**).

Next, we performed bootstrap analyses with subject-level resampling to explore whether the *de novo* mtDNA SNV classes exhibited differences in predicted impact and/or population frequency. We found that the mtDNA variants predicted to be pathogenic were significantly overrepresented among TU SNVs (bootstrap *P* <0.0001) but significantly underrepresented among the GH (bootstrap *P* <0.0001) and SH (bootstrap *P* <0.0001) SNVs (**Figure 1E**). Conversely, substitutions within the D-loop (a noncoding control region) and variants predicted to be benign were significantly enriched among the GH (*P* <0.0001, *P* = 0.0006) and SH (*P* = 0.0098, *P* = 0.0176) SNVs but significantly underrepresented among those in the TU group (*P* <0.0001 for both) (**Figure 1E**). In terms of population frequency, we found that whereas ∼82% of the INH variants (*n* = 14,495) were common variants (MitoMap frequency >1%), more than 90% of the TU variants (*n* = 283) were rarely observed (MitoMap^52^ frequency <0.1%) (**Figure 1F**). Integration of TU VAF distribution with population frequency and predicted impact tabulations revealed that most rare, functionally impactful variants in the TU class existed at VAFs of less than 0.25 (**Figure 1G**).

The relatively high proportion of pathogenic variants in the TU class was expected, given the mtDNA mutational signature, which suggests replication errors as a source of the mutations^10,54,55^. However, assuming that the source of mutations is the same for *de novo* and inherited variants^10^, the differences in the prevalence of benign *vs*. pathogenic mtDNA variants between the TU variants and the INH, GH, and SH variants suggested that different classes are under different constraints or selective pressures; namely, negative selection against pathogenic mtDNA mutations in nontumor cells and relaxation of this negative selection in tumor cells. This notion is also supported by a comparison of the mutational signatures among the different classes of *de novo* mtDNA variants (**Supplementary Note 4**).

Another way to ascertain selection pressure is to determine whether the ratio of nonsynonymous-to-synonymous substitutions (dN/dS) is greater than (positive selection) or less than (negative selection) neutrality^56,57^. Taking into consideration the mutational signature associated with mtDNA mutations, the calculated dN/dS^40^ was lower than the expected neutrality ratio among the INH, GH, and SH mtDNA variants (**Figure S2A**); however, it was higher than expected among TU variants in specific tumor subtypes (**Figure S2B**), including two B-cell acute lymphoblastic leukemia (B-ALL) subtypes (i.e., ETV6–RUNX1^+^ B-ALL and E2A/TCF3–PBX1^+^ B-ALL). Consistent with the dN/dS analyses performed on the pediatric tumor subtypes, the proportion of TU variants predicted to be pathogenic was significantly (bootstrap *P* = 0.028) higher in certain hematological malignancies (e.g., E2A/TCF3–PBX1^+^ B-ALL and DUX4r) as compared to other tumor types (**Figure 1H**, **Figure S2C**, and **Supplementary Note 6**). In contrast, lymphomas had the lowest dN/dS among adult cancers (**Figure S2D**). Collectively, these results suggest that unlike those in adult populations, pathogenic mutations in children are positively selected during the development of lymphoid malignancies and therefore may contribute to pathogenesis.

Using bootstrap analysis again, we next compared the cumulative VAF distributions between nonsynonymous and synonymous SNVs among pediatric and adult tumor samples. Among pediatric samples we noted a clear difference in cumulative VAFs between nonsynonymous and synonymous SNVs, which was most pronounced among hematological malignancies (**Figure 1I-J** and **Figure S2E**). By contrast, we observed overlapping profiles between synonymous and nonsynonymous SNVs in adult tumors (from TCMA, The Cancer Mitochondrial Atlas),^41,58^ including hematological malignancies (**Figure 1I-J**, lower panel; and **Figure S2E**). These results suggest that synonymous and nonsynonymous TU mtDNA SNVs in pediatric leukemias are subject to different selective pressures that may restrict their accumulation to varying degrees.

### Pathogenic TU variants accumulate to intermediate levels in subsets of cells

Bulk WGS estimates the overall burden of mutated mtDNA across a population of cells but fails to indicate the extent of VAF heterogeneity within individual cells – for example, populations in which all cells have a uniform VAF and populations with uneven VAF distributions may have the same average VAF by bulk WGS. Discerning between these two scenarios is crucial, as the cellular consequences and clinical phenotypes associated with pathogenic mtDNA mutations vary in a VAF-dependent manner^14,18,24,29,59^. Moreover, knowledge regarding the VAFs in individual cells may also clarify which selective pressures are at play. Therefore, using the C1 microfluidics-based platform (Fluidigm), we first explored whether single-cell mtDNA sequencing (sc-mtDNAseq) could capture the VAF distribution of established pathogenic mtDNA variants among individual leukemic cells.

To this end, we selected a B-ALL sample (*MLL*-rearranged infant leukemia, INF010), which based on our WGS analyses harbored a TU m.3243A>G mutation that has been observed in several adult tumor samples^60–63^ (**Supplementary Note 2**). When maternally inherited, this mutation can cause MELAS (mitochondrial myopathy, encephalopathy, lactic acidosis, and stroke-like episodes syndrome)^13,64,65^ and other primary mitochondrial disorders^13,31,66–69^, with compromised respiratory function and reduced OXPHOS being well-documented consequences of the mutation^24,70–72^. The INF010 sample also harbored a second TU mtDNA variant (m.72T>C), which is a recurrent D-loop variant seen in the PCGP and adult TCMA^41,58^ cohorts (**Supplementary Note 2**, and **Table S1C**) and in various noncancerous adult tissues^73^.

Our sc-mtDNAseq analyses revealed several differences between the VAF distributions of the two TU SNVs and other *de novo* mtDNA variants (i.e., 1 SH variant and 1 GH variant) identified in this B-ALL sample (**Figure 2A, Table S1D**, and **Supplementary Note 7**). Unlike the SH variant (m.4674A>G *MT-ND2*, bulk VAF=0.266), which was consistently present in all 19 cells at low allele fractions (interquartile range [IQR] = 0.053-0.248 VAF), the two TU variants were present at higher VAFs but only in subsets of leukemic cells (**Figure 2A**), suggesting that the SH and TU variants are under different selective pressures. Additionally, there were subtle differences in the VAF distributions between the two TU mtDNA variants. For example, sc-mtDNAseq detected the m.72T>C mutation (bulk VAF = 0.124) at high heteroplasmy levels among 5 of 19 blasts (0.55-0.93 VAF), and the m.3243A>G mutation (bulk VAF = 0.222) in 11 of 19 blasts at intermediate levels of heteroplasmy (0.20-0.70 VAF). Prior studies using cybrids demonstrated that the m.3243A>G mtDNA mutation not only impairs OXPHOS at VAFs >0.50 but also influences epigenetic marks and nuclear gene expression at VAFs as low as 0.20^24^. Thus, our sc-mtDNAseq results demonstrate that within individual leukemic cells, pathogenic variants can accumulate to intermediate levels at which they are predicted, based on data from cybrids^24^, to reduce (but not abolish) OXPHOS and incite compensatory cellular responses. In contrast, certain variants in the regulatory D-loop region may reach higher heteroplasmic levels due to the mtDNA replicative advantage that they confer^74,75^.

**Figure 2.**
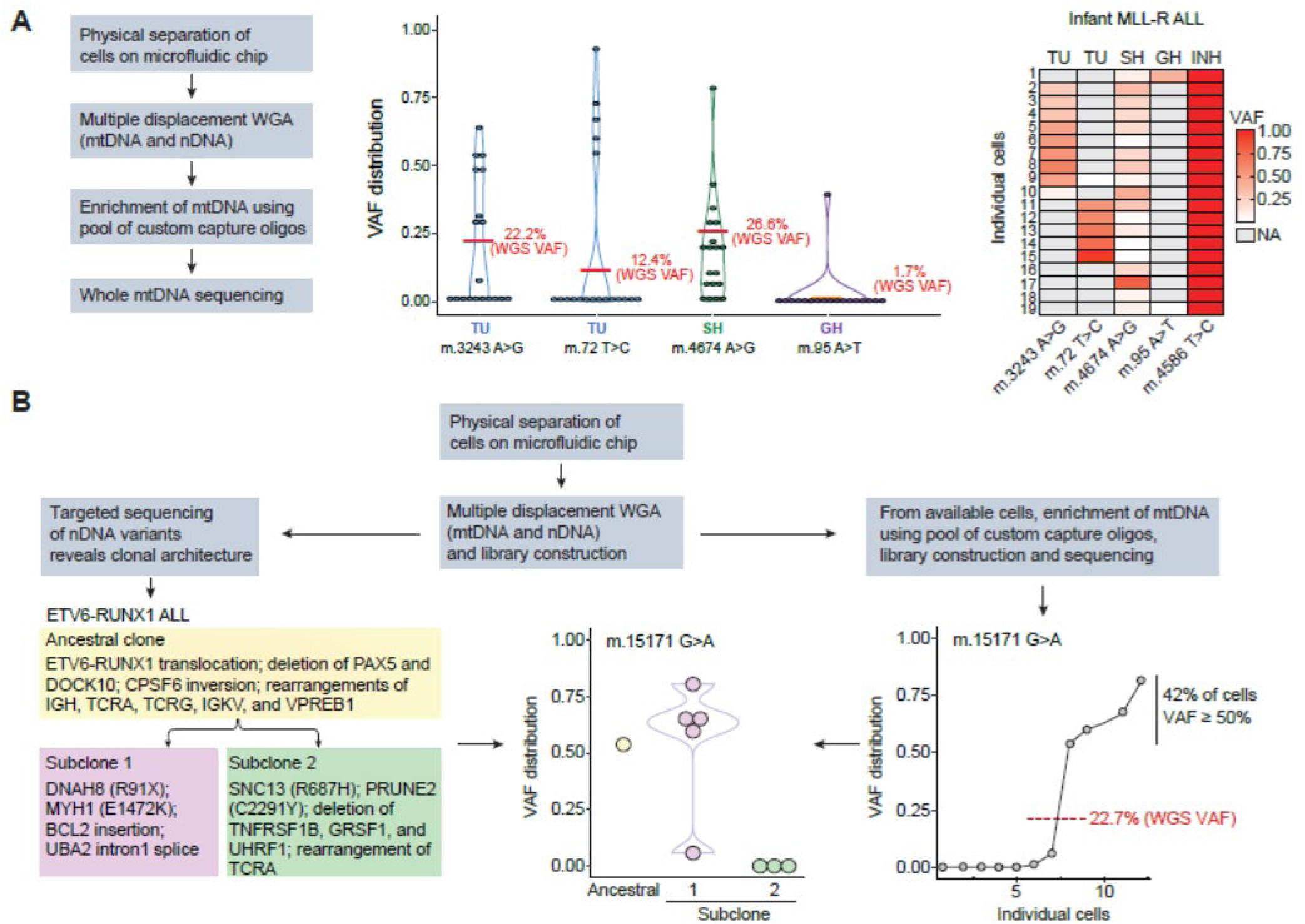
Single-cell DNA sequencing-based characterization of tumor-enriched, pathogenic mtDNA variants in two patients with B-ALL reveals intermediate VAFs in a subset of cells. Experimental sc-mtDNAseq overview and corresponding single-cell VAF results for select mtDNA variants. **(A)** Results from an infant *MLL*-rearranged B-ALL sample (INF010); a violin plot (middle panel) depicts VAF at single-cell resolution for each *de novo* mtDNA variant in the sample. Individual points represent single cells; red lines indicate WGS VAF estimates from the bulk tumor. The heatmap (right panel) shows the VAF for each *de novo* variant and one inherited variant on a cell-by-cell basis. White cells indicate an absence of mutation. Notably, TU variants were present at intermediate levels in mutually exclusive subsets of cells, and the SH variant was present in almost all cells. **(B)** Results from an ETV6–RUNX1^+^ B-ALL sample (ETV027); select cells that were originally sequenced for a clonal architecture study (left panel) were re-sequenced to detect a TU variant that impairs complex III activity. Nuclear SNVs and rearrangements identified from the original study are listed below each clone (ancestral, subclone 1, subclone 2). Violin plot (middle panel) illustrates TU VAF patterns of individual cells by clonal assignment; scatterplot (right panel) highlights the increased VAF among subsets of individual cells. The bulk WGS VAF estimate for this same variant (red line) was 22.7%. **Abbreviations**: B-ALL, acute B-lymphoblastic leukemia; GH, germline heteroplasmy; INH, inherited germline heteroplasmy; sc-mtDNAseq, single-cell mitochondrial DNA sequencing; SH, shared heteroplasmy SNV, single-nucleotide variant; TU, tumor-enriched; VAF, variant allele fraction; WGS, whole-genome sequencing.

### The expansion or loss of TU mtDNA variants contributes to subclonal diversity

In the INF010 sample, the two TU variants were distributed among a mutually exclusive subset of cells (**Figure 2A**, right panel), suggesting that the variants were acquired at different times during leukemogenesis and/or in distinct populations of leukemic cells. Thus, we next explored whether we could determine the timing of mtDNA mutation acquisition in a leukemic sample. Here, we leveraged data from a prior scDNAseq study that inferred the subclonal architecture of an ETV6–RUNX1^+^ ALL sample (ETV027) based on mutational cluster analysis of nuclear DNA-encoded variants sequenced from individual cells^76^. As previously described, this mutational cluster analysis identified two populations that arose from a common ancestral clone harboring the *ETV6–RUNX1* translocation^76^ (**Figure 2B**). Our re-analysis of the bulk WGS data revealed that this sample harbored the TU mutation m.15171G>A (VAF = 0.227). This mutation, which has been reported in adult tumors (**Supplementary Note 2**), occurs in the gene that encodes cytochrome b (*MT-CYB*) and results in a p.G142E substitution. Functionally, the mutation is predicted to disrupt ubiquinol ligand binding within the catalytic machinery of the cytochrome bc1 complex (complex III)^77–79^ and impair its activity^80^ as a consequence of increased hydration and reduced pocket volume at the ligand-binding site (**Supplementary Note 8**). Seeking to determine the allele fraction of the m.15171G>A mutation, we performed whole-mtDNA genome amplification and sequencing using DNA from a small subset of single cells (based on available material) whose clonal membership (i.e., ancestral clone, subclone 1, subclone 2) was previously established in the ETV027 sample^76^. The sc-mtDNAseq data were then used to calculate the VAF of the m.15171G>A mutation in individual leukemic blasts (**Table S1E**). Again, as observed with the m.3243A>G mutation in INF010, a subset of blasts (42%) harbored the m.15171G>A mutation at an intermediate VAF range (0.537-0.803; **Figure 2B**)––levels that are predicted to affect complex III activity^27^.

Grouping the re-sequenced blasts based on clonal membership revealed only one cell with the nuclear mutational signature of the ancestral clone. This cell harbored the mtDNA mutation at an allele fraction of 0.537 (**Figure 2B**). We also observed a difference in partitioning of the m.15171G>A mutation between the two derivative clones, with elevated VAFs in most of the cells in the first clone and complete elimination of the variant from the second clone. Given the low sampling of the ancestral clone, we cannot exclude the possibility that the mtDNA mutation was acquired within this population. Nevertheless, in the context of clonal evolution, these results suggest that the acquisition or expansion of TU mtDNA mutations can occur early in leukemogenesis. Moreover, the differential co-segregation of pathogenic mtDNA mutations in leukemic cells with related, but distinct, leukemia-associated nuclear genomes raises the possibility that the accrual of certain mtDNA mutations influences or is influenced by clonal evolution.

### Detecting mtDNA VAFs by scRNAseq

Our results suggest that pathogenic variants can accumulate to levels at which, based on prior studies^24,27,32,81–83^, they could incite cellular responses. Since estimates of the mtDNA VAFs were highly concordant between RNAseq and WGS, we next sought to determine whether single-cell RNA sequencing (scRNAseq) could be used to simultaneously estimate mtDNA VAF distributions in larger populations of cells and capture cell-state changes associated with pathogenic TU mtDNA variants in primary leukemic blasts.

Three B-ALL tumor samples harboring the t(1;19)(q23;p13) translocation (E2A–PBX1^+^)^84^ were selected for scRNAseq based on WGS results, which highlighted this subtype’s propensity for harboring one or more potentially pathogenic TU mtDNA mutations within a single sample (**Figure 1H**). Samples were chosen based on whether their mtDNA variants of interest were within the range of coverage imposed by the assay [i.e., maximal coverage within 300 bp of the poly(A) tail] (**Supplementary Note 9**). Two of the samples harbored variants reported in adult tumors (m.8172G>A and m.11889G>A) and one of them harbored a variant affecting a recurrently targeted amino acid (m.15657T>A, I304 MT-CYB) (**Supplementary Note 2**). After passing standard QC thresholds, which excluded cells with high mtDNA unique molecular identifiers (UMIs) (**Table S2A**), more than 1,500 cells (range: 1,591-2,622) from each of the three diagnostic B-ALL samples (E2A037, E2A015, E2A025) were analyzed. Based on the expression of nuclear-encoded RNA transcripts, individual cells from each sample were partitioned into transcriptionally distinct clusters and annotated with common lineage markers to distinguish leukemic blasts from residual normal hematopoietic elements present in the diagnostic sample (**Figures S3A-B**, **S4A-B**, and **S5A-B**). Blasts comprised ≥85% of cells in all samples, consistent with the flow cytometry–based enumeration of blasts at diagnosis (**Table S2A**).

A critical component of our downstream analyses was the ability to detect mtDNA variants previously identified by WGS. As exemplified by sample E2A037, which harbored two TU variants (i.e., m.11865T>C and m.8172G>A), UMIs covering each of the variant locations were detected in ≥97% of cells (IQR: 8-16 UMIs/cell for m.11865, and 13-30 UMIs/cell for m.8172), giving us high confidence in VAF assignments (**Figure S3C-D**, and **Table S2B**). Comparable coverage was observed for the m.15657T>A (E2A025; IQR, 3-10 UMIs/cell) and m.11889G>A (E2A015; IQR, 12-26 UMIs/cell) TU mtDNA variants present in the two remaining E2A–PBX1^+^ samples (**Figures S4C, S5C**, and **Tables S2C-D**). The m.1200G>A variant in sample E2A015 showed less coverage (IQR, 2-5 UMIs/cell) using the 10× sc-RNAseq strategy, but this was not unexpected given its position in the mitochondrial genome (**Figure S5D**).

Next, we projected the mtDNA VAFs and coverage onto the clusters of cells established by the initial t-distributed stochastic neighbor-embedding (tSNE) plots (**Figure 3A-D**, left panels and **Figure S5E**). These analyses showed the expected enrichment of TU variants (e.g., m.8172G>A and m.11865T>C in E2A037) among the blasts, as compared to normal cell populations (**Figure 3A-D**, middle panel and **Figure S5F**). Nevertheless, the presence of TU mtDNA variants in small populations of non-blast hematopoietic cells (i.e., T-cells, monocytes, or erythroid cells) suggests that leukemia-initiating hematopoietic stem/progenitor cells acquired the relevant somatic mtDNA mutation(s) either before or during the initiation of malignant transformation, such that derivatives of the cell could still differentiate into mature hematopoietic lineages. We also noted that the m.1200G>A and m.11889G>A mutations did not coexist in blasts from the E2A015 sample (**Figure S5G**), but 78% of blasts from the E2A037 sample harbored both TU mtDNA variants (m.8172G>A and m.11865T>C; **Figure S3E**).

**Figure 3.**
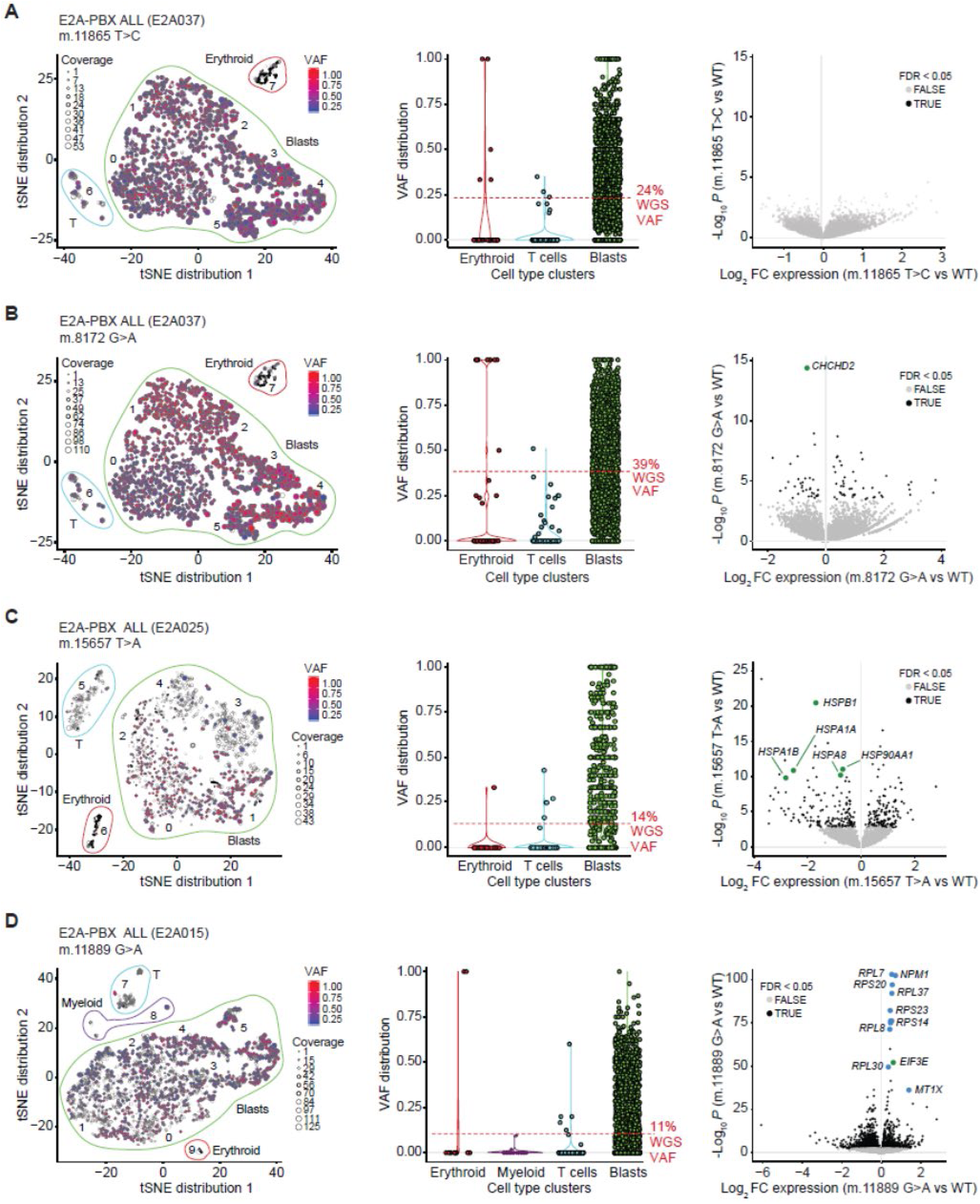
scRNAseq reveals that pathogenic, TU mtDNA mutations are subject to variant-specific forms of selective pressure that are commensurate with changes in gene expression. Single-cell RNA sequencing (scRNAseq) provides critical insight into mtDNA VAF distributions, operating selective pressure, and cell-state changes associated with pathogenic TU mtDNA variants in primary leukemic blasts. Each row represents a specific TU mtDNA variant. **(A-D, left panels)** Projection of variant allelic fractions (VAF) onto cells within discrete clusters identified by each sample’s initial tSNE analysis. Scatter point sizes reflect variant coverage. Cells are colored to reflect the magnitude of the VAF. **(A-D, middle panels)** Single-cell VAF distribution of TU variants among different cell-type clusters reveals early appearance of somatic mtDNA mutations; red dashed line indicates the bulk tumor WGS estimate. **(A-D, right panels)** For each TU mutation, a volcano plot examining differential gene expression between cells harboring the mutation (alternate UMI ≥ 20.0%) or WT allele only (reference UMI ≥ 5 and alternate UMI = 0.0%), with *P* values (-log_10_) on the y-axis and expression (log_2_ fold change) on the x-axis. Left side of plot denotes genes with reduced expression in mutant cells; right side of plot denotes increased expression of genes in mutant cells. Differentially expressed genes with FDR<0.05 are depicted in dark gray. Green dots represent genes where relationships between expression and heteroplasmy levels were explored further; blue dots represent notable differentially expressed genes. **Abbreviations**: *CHCHD2*, Coiled-Coil-Helix-Coiled-Coil-Helix Domain-Containing Protein 2; *EIF3E*, Eukaryotic Translation Initiation Factor 3 Subunit E; FC, fold change; FDR, false discovery rate; *HSP*, heat shock protein; *MT1X*, Metallothionein 1X; *NPM1*, Nucleophosmin 1; *RPL* or *RPS*, ribosomal protein; tSNE, t-distributed stochastic neighbor embedding; WGS, whole genome sequencing.

### scRNAseq reveals mtDNA variant–specific changes in the expression of nuclear DNA–encoded genes

After establishing a robust method for calling mtDNA VAFs at the single-cell level, and since pathogenic mtDNA mutations can influence transcriptional programs at VAFs as low as 20%^24,32^, we next compared differential gene expression patterns in leukemic blasts harboring TU mtDNA mutations (alternate UMI ≥20%) and those with WT mtDNA (reference UMI ≥5; alternate UMI = 0%) (**Tables S2E-I**). We did not detect any significant changes in gene expression associated with the m.11865T>C variant in sample E2A037, despite the moderate functional impact that was predicted (Mutation Assessor score, 3.06) for this p.L369S substitution within MT-ND4 (i.e., NADH: ubiquinone oxidoreductase, core subunit 4 of complex I) (**Figure 3A**, right panel). In contrast, we identified several genes whose expression was altered in leukemic cells harboring the m.8172G>A variant in *MT-CO2*, which encodes the second subunit of cytochrome c oxidase (COX; i.e., complex IV) (**Figure 3B**, right panel and **Table S2F**). A greater number of differentially expressed genes was identified in leukemic cells harboring the m.15657T>A (E2A025) variant that resides within *MT-CYB* (**Figure 3C**, right panel, and **Table S2G**). Variant-specific associations with gene expression were also evident within the E2A015 sample; in this case, the m.11889G>A variant (in *MT-ND4*) demonstrated the greatest number of significant gene expression changes among all TU mtDNA variants that were studied (**Figure 3D**, right panel, and **Table S2H**). Meanwhile, minimal changes in gene expression were detected in association with the m.1200G>A variant in *MT-RNR1* (**Table S2I**), perhaps because of the small number of cells harboring the variant.

Genes implicated in mitochondrial function and/or metabolism were among the most differentially expressed genes in leukemic cells from individual samples harboring WT or mutated mtDNA. For example, in blasts harboring the m.8172G>A *MT-CO2* variant, the gene *CHCHD2* (aka *MNRR1*) was dysregulated, as determined by negative binomial regression (false-discovery rate [FDR]-adjusted *P* = 4.01 × 10^-11^) and permutation-based testing (**Figure 3A**, right panel and **Table S2F**). This observation is interesting, given that *CHCHD2* encodes a protein that engages with COX and can translocate from the mitochondria to the nucleus during stress^85^. The negative correlation between *CHCHD2* expression and increasing heteroplasmy of the m.8172G>A variant (**Figure S3F**) is also reminiscent of the decreased *CHCHD2* expression reported in cybrid cell lines harboring the pathogenic m.3243A>G mutation^86^. As a result of the m.8172G>A mutation, a cysteine-to-tyrosine substitution occurs at position 196 (p.C196Y) that is predicted to disrupt copper binding (**Figure S3G**), which is required for the catalytic activity and maturation of COX^87,88^. To test whether disruption of COX activity due to copper displacement reduces *CHCHD2* expression, we treated a B-ALL cell line with increasing concentrations of ATN-224 (bis-choline tetrathiomolybdate), a high-affinity copper chelator that inhibits complex IV function^89^. After 24 h of ATN-224 treatment, we observed a significant (*P* = 0.024, ANOVA) dose-dependent decrease in *CHCHD2* mRNA (**Figure S3H**).

The m.15657T>A variant in *MT-CYB* replaces a hydrophobic isoleucine with a polar asparagine at position 304 in helix F, which is predicted to disrupt a network of hydrophobic interactions with surrounding residues from connecting helices. The mutation may also de-stabilize the protein, or–– since helix F forms the entrance to the Qo site––affect ligand entry (**Figure S4D**). Similar to what was reported for the disease-causing m.3243A>G mutation^24^, the m.15657T>A variant was associated with dysregulation of chaperone-encoding genes (**Figure 3C**, right panel, and **Table S2G**), many of which have been implicated in mitochondrial maintenance and function. For example, *HSP90AA1* and *HSPA8* encode HS90A and HSP7C, respectively, and are implicated in mitochondrial pre-protein import^90^. *HSPA1A* encodes HSP72, which translocates to depolarized mitochondria and is essential for proper function of Parkin, an E3 ubiquitin ligase involved in mitophagy^91,92^. *HSPB1* encodes the chaperone HSP27, which has been implicated in mitochondrial QC^93^. Thus, the reduced expression of *HSP* genes associated with the presence of m.15657T>A (**Figure 3C**, right panel) may constitute a compensatory response in which complex III dysfunction reduces mitophagy levels to increase mitochondrial content.

In the case of the E2A015 sample, the m.11889G>A variant was associated with significant (*P* < 10^-5^) changes in the expression of many nuclear DNA–encoded genes, including upregulation of genes encoding ribosomal proteins (e.g., *RPL7*, *RPS20*, *RPL37*, *RPS23*, *RPS14*, *RPL8*, *RPL30*) and those involved in targeted translational programs and accelerated protein synthesis (*EIF3E*, *NPM1*)^94–99^, mitigation of oxidative stress (*MTX1*, *NPM1*)^100–104^, and subversion of stress-induced apoptotic signaling (*NPM1*)^104–106^ (**Figure 3D**, right panel, and **Table S2H**). Induction of *EIF3E* was particularly intriguing, as its gene product enhances the synthesis of proteins with membrane-associated functions, including ETC components, and is critical for preserving mitochondrial function^107,108^. Further investigation of the relationship between *EIF3E* expression and increasing m.11889G>A levels revealed a positive correlation (**Figure S5H**), suggesting that m.11889G>A heteroplasmy alters MT-ND4 function and triggers a compensatory EIF3E response to regulate cellular respiratory activities^107,108^. Of note, the m.11889G>A variant results in a p.G377E substitution that may profoundly affect MT-ND4 function (Mutation Assessor score, 4.81) as the G377 residue is located within a discontinuous helix (**Figure S5I**) that forms a hydrophobic, protein–protein interface between MT-ND4 and MT-ND5 (NADH: ubiquinone oxidoreductase core subunit 5, complex I)^109^. Inward-facing residues, such as G377, tilt the lower half of the discontinuous ND4 helix at an angle so that helix–helix packing is favored; thus, the inability to place the charged glutamate (E377) residue in the required inward-facing conformation most likely disrupts helix packing. Together, these results highlight variant-specific changes in the expression of nuclear DNA-encoded genes associated with tumor-enriched mtDNA variants, affecting both the identity and the number of genes impacted.

### Statistical modeling suggests that selective pressures shape mtDNA VAF distributions in a variant-specific manner

We aimed to determine whether mtDNA variant-associated changes in gene expression were sufficient to influence cell state. We hypothesized that if these changes impacted cell state in ways that were either beneficial or detrimental to leukemia development, the variants would be subjected to positive or negative selection. Conversely, if the mtDNA mutations were functionally inert, we would expect no evidence of selection. To test this hypothesis, we developed Mitovolve, a statistical model that utilizes single-cell sequencing data to infer the evolutionary history of somatic mtDNA mutations.

Mitovolve assumes that an initiating leukemic cell has a fixed mtDNA copy number (derived from its WGS estimate) with initial VAFs (iVAF) ranging from 0 to 1.0. The probability distribution of mutant mitochondrial genomes in each daughter cell is sampled from a pool of duplicated mitochondrial genomes within the parent cell, a process reiterated for each successive generation of daughter cells (**Figure S6A-C**). Hypergeometric sampling is used for models without selection, whereas selection models sample a noncentral hypergeometric distribution with the log odds ratio (logOR) of selecting a mutant mtDNA defined as a cubic polynomial function of the VAF^110,111^. Given these initial parameters, the model (i.e., favoring either homoplasmic or heteroplasmic accumulation of mutant mtDNA, or no selection) that ultimately best fits the observed data can be identified.

For each of the four mutations shown in Figure 3, we first evaluated 30,351 – 62,913 selection-free models per variant (fixed mtDNAcn; iVAFs: 0.01-1 in increments of 1/mtDNAcn; generations: 0-200). The limit of 200 generations was determined empirically to capture the best selection-free model for each variant. Using our scRNAseq UMI-count data, we calculated the probability distribution and the negative log-likelihood (nLL) for each selection-free model^112^ and identified the best fit model with the minimum nLL. Since our null hypothesis assumed no selection, the best fit selection-free model was p.null ∼ 1. We expanded our analysis to include models that were not significantly different from this best fit model (p.null > 0.05) to yield a narrow range of models that closely approximated the observed data (**Table S2J** and **Figure S6D**). Next, unique starting parameters (i.e., iVAF and generation number) from each of these top selection-free models were used to perform selective pressure modeling. This allowed us to identify the selection beta coefficients associated with each best-fit selection model (**Table S2J**). Likelihood ratio testing was then utilized to identify which (if any) of the best-fit selection models outperformed the best-fit selection-free model (p <0.05) (**Table S2J**).

To visualize the nature of the selective pressures that may be shaping the observed VAF distributions for each of our variants, we generated plots for each variant showing the logOR cubic function for each of the best fit selection models (**Figure 4A-C**, top panel; **4D**, left panel). Lines depicting each model were color-coded based on p-values (p.alt) and illustrate the first notable difference among the variants – that with the exception of the m.11865T>C variant (p >0.07), all variant models with selection were predicted to fit the observed data better than their respective best selection-free models (p < 4.6×10^-5^ for m.8172G>A; p <1. 06×10^-22^ for m.15657T>C; p <2.89×10^-19^ for m.11889G>A (**Figure 4A-C**, top panel; **4D**, left panel). Although the magnitude and direction of the selection pressure predicted by the models can vary with different starting parameters (i.e., iVAF and number of generations; **Table S2K**), using the starting parameters from the top selection-free models (**Table S2J**), there were appreciable variant-specific differences in the logOR. With the exception of m.11865T>C, the plots suggest that the variants accumulate to intermediate VAF levels and then undergo negative selection at higher VAFs.

**Figure 4.**
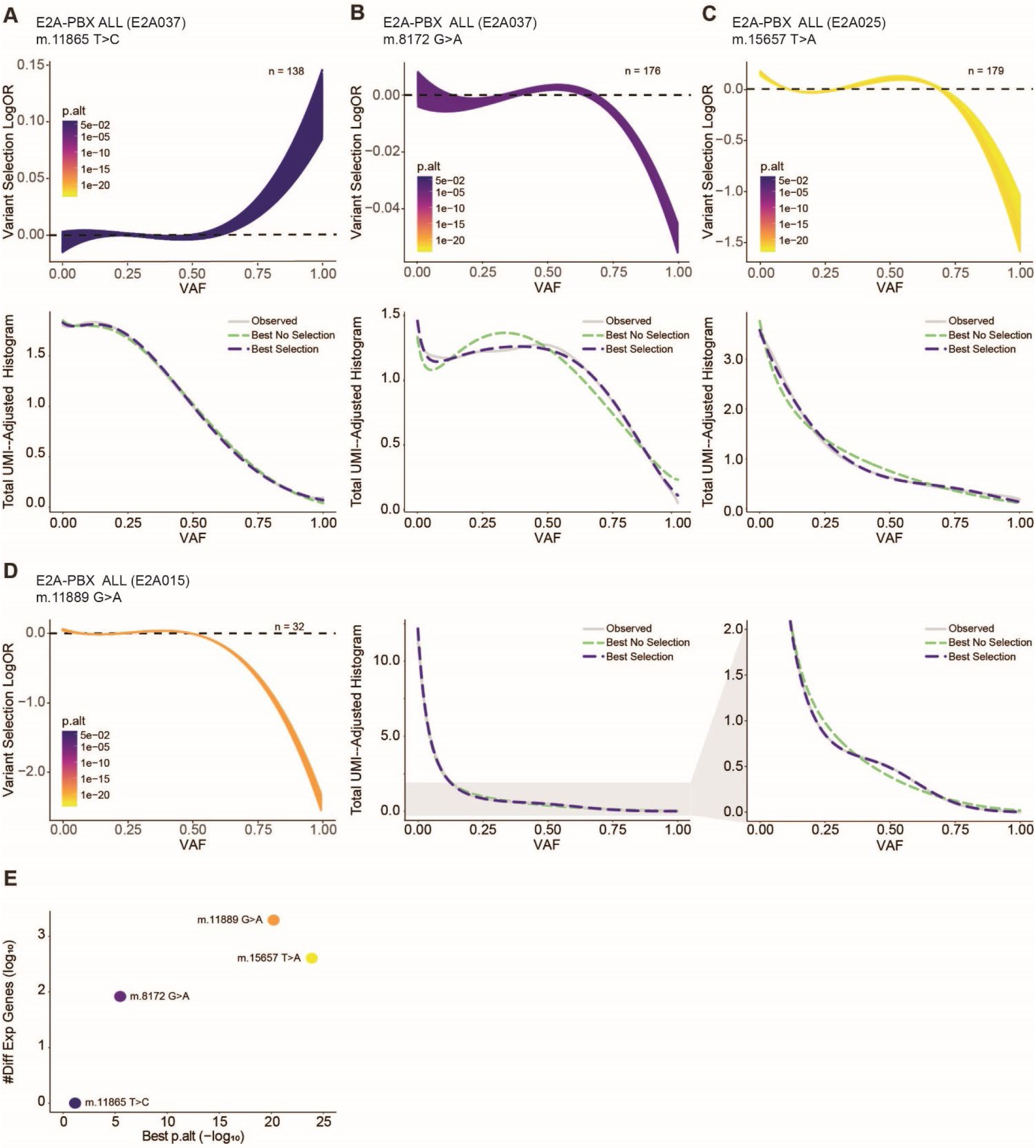
Statistical modeling suggests that selective pressures shape mtDNA VAF distributions in a variant-specific manner. **(A-C, top panel; D, left panel)**: For each variant, a series of best-fit selection models were identified using starting parameters defined by the best no-selection models (i.e. p.null >0.05). The models were projected onto a plot of logOR against VAF to gauge the magnitude and direction of selective pressure operating on each variant. Positive values for the logOR indicate positive selective pressure for the variant while negative values indicate negative selective pressure. **(A-C, bottom panel)** Smoothened total UMI-adjusted histograms show comparison of best selection model versus the best selection-free model and the observed data for each of the following variants: m.11865T>C, m.8172G>A, and m.15657T>C. **(D, middle and right panel)** For m.11889G>A, smoothened total UMI-adjusted histograms (middle) with expanded view (right) highlight the comparison between the best selection model versus the best selection-free model and the observed data. With the exception of m.11865T>C, the best selection model approximates the data better than the best no selection model. **(E)** Graph demonstrating the relationship between number of differentially expressed genes (FDR>=0.05) for each variant plotted and the strength of support (*i.e.* p.alt) for selection models approximating the observed data. **Abbreviations**: iVAF, initial variant allele fraction; logOR, log odds ratio; p.alt, statistical test comparing the best selection model with the best non-selection model.

We next generated smoothened histograms of the inferred frequency of cells with increasing VAFs (adjusted based on UMI counts) to compare the best-fit selection model with the lowest p-value (p.alt) to the best-fit selection-free model with the highest p-value (p.null) and to the observed data (**Figure 4A-C**, bottom panel; **4D**, middle and right panels). Here, we find that the models with selection fit the observed data better than the best selection-free model for all variants except for the m.11865T>C variant, where the lines were virtually indistinguishable. Finally, using the most significant p.alt-value as an indication of the likelihood that selection played a role in shaping the VAF distribution, we found a positive relationship between the number of differentially expressed genes (FDR ≥ 0.05) associated with each variant and the degree of support for its best selection model (**Figure 4E**). Thus, combining the evidence for selection with variant-associated gene expression changes may provide a means of distinguishing somatic mtDNA mutations that are likely to affect cellular function and/or growth properties (*e.g.*, m.11889G>A) from those that are functionally inert (*i.e.,* m.11865T>C).

### NetBID identifies hidden drivers and pathways associated with pathogenic mtDNA variants

We next further investigated the cell state changes associated with the three mtDNA mutations for which there was evidence of selection. To do this, we turned back to the expression data and addressed a common issue with single-cell transcriptomic data-the inconsistent or lost expression (i.e., “dropout”) of transcripts in cells^113^. Moreover, many signaling proteins and transcription factors that dictate cellular function are not differentially expressed at the mRNA level but are instead regulated by posttranslational or other modifications. Therefore, changes in their activities may be “hidden” by using conventional gene expression profiling approaches. To recover information about cell state despite lost transcripts and in recognition that important biological activities are often driven by “hidden” drivers, we deployed a network-based, systems biology approach, (i.e. Network-based Bayesian inference of drivers, or NetBID)^114^ to infer changes in the activity levels of these drivers in cells harboring somatic mtDNA variants.

For each TU mtDNA variant with significant (FDR <0.05) changes in gene expression (i.e., m.8172G>A, m.11889G>A, or m.15657T>A), blasts were dichotomized as being either mutant (alternate UMI ≥20%) or WT (reference UMI ≥5%. alternate UMI = 0%). Next, we used SJARACNe (v.0.2.1), an information theory–based algorithm for gene network reverse engineering^115^, to reconstruct a B-ALL–specific network of interactions between hub genes (1,643 transcription factors and 6,247 signaling factors) and their downstream targets from the bulk RNAseq-derived transciptomes of 185 pediatric B-ALL cases (TARGET cohort)^116^. The expression-based interactome encompassed 21,659 genes and 830,215 interactions, which were weighted based on the strength of the interaction between each hub-target pair^117^. The scRNAseq datasets were then projected onto the SJARACNe B-ALL network, and the activity of each hub gene in each cell was inferred based on the aggregate weighted expression of its targets^117^. Using NetBID’s Bayesian linear–modeling function on blasts within each sample that harbored WT or mutant mtDNA, we then calculated differential activity (DA) scores and identified genes with significantly (*P* <10^-5^) higher or lower activities in mutant *vs.* WT cells (**Figure 5A**).

**Figure 5.**
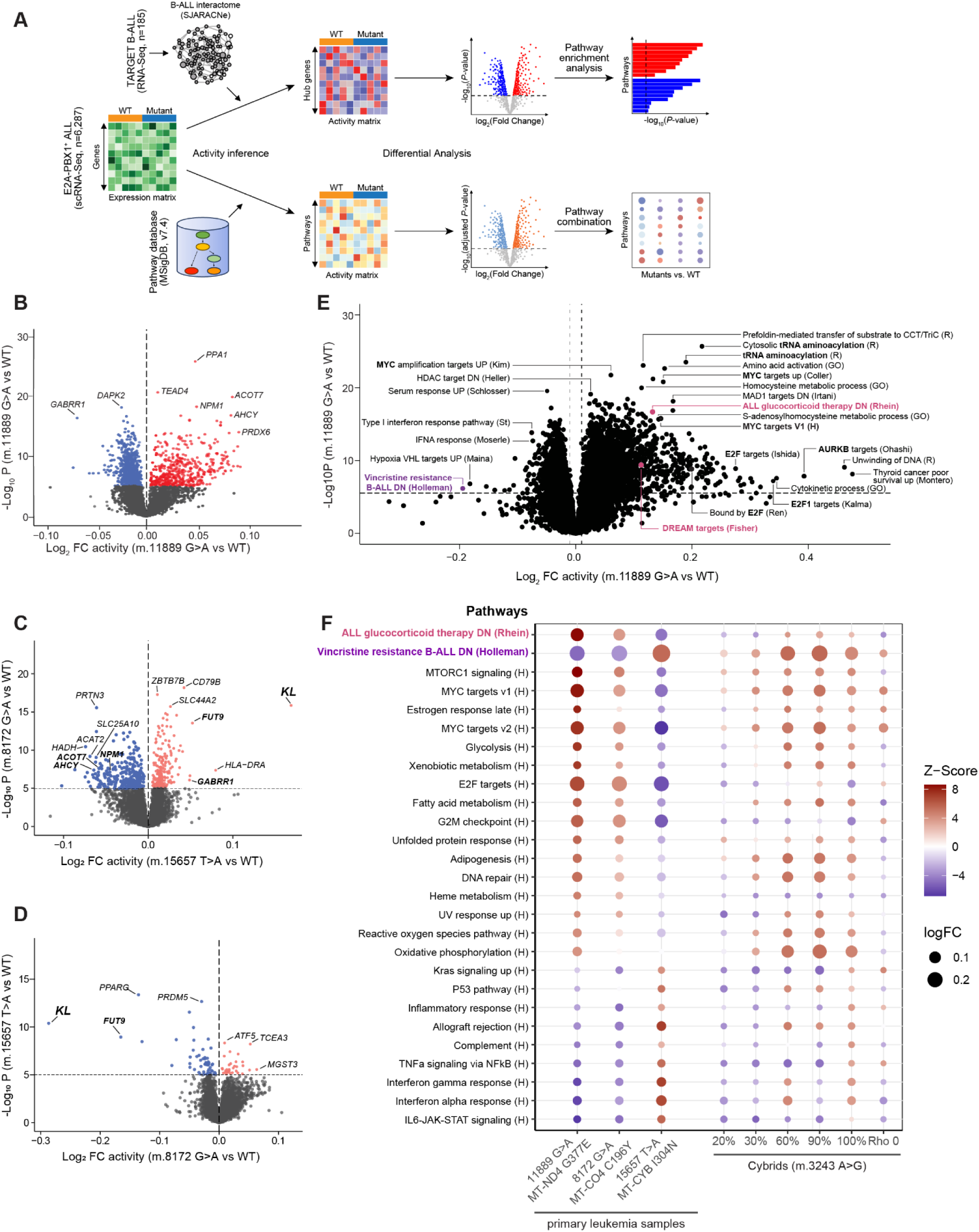
NetBID analyses highlight cell state changes associated with the presence of tumor-enriched mtDNA variants at intermediate levels of heteroplasmy. **(A)** Schematic for NetBID^114^. Using SJARACNe^115^, a B-ALL interactome was constructed from an RNAseq dataset comprising 185 pediatric B-ALL samples (TARGET cohort)^116^. Single-cell RNA-sequenced blasts from each TU mtDNA variant with significant (FDR <0.05) changes in gene expression (i.e., m.8172G>A, m.11889G>A, m.15657T>A) were dichotomized as either mutant (alternate UMI ≥20%) or WT (reference UMI ≥5 and alternate UMI = 0%). The scRNAseq datasets were then projected onto the SJARACNe B-ALL network, and the activity of each hub gene in each cell was inferred based on the aggregate-weighted expression of its targets. DA scores were then calculated and used to identify genes with significantly (*P* <10^-5^) higher or lower activities in mutant *vs.* WT cells. Similarly, the activity of each selected MSigDB^187^ pathway in each cell was estimated by averaging the expression of genes involved in the pathway. Then, the differentially activated hub genes (P < 10-5) and pathways (FDR-adjusted P < 10-5) were identified using NetBID’s Bayesian linear modeling function. The Fisher’s exact test-based pathway enrichment analysis of the hub genes with significantly higher or lower activities in mutant vs. WT cells were performed separately. The differentially activated pathways in mutant vs. WT cells across all mtDNA variants were combined and visualized with bubble plot. **(B-D)** Volcano plots reflecting differential activation of drivers in cells harboring specific somatic TU mtDNA mutations, as compared to those harboring only WT mtDNA. The left side of the plot denotes drivers with reduced activity in mutant cells; the right side denotes increased activity of drivers in mutant cells. The dashed black horizontal line denotes a conservative threshold for significance (*P* = 10^-5^). **(E)** NetBID’s linear modeling reveals significant (FDR-adjusted *P* <10^-5^) differences in pathway activities between the m.118889G>A mutant and WT cells. The volcano plot highlights pathways affected by the m.11889G>A variant; the top pathways identified by Fisher analysis are highlighted in dark pink, and the other pathways of interest are highlighted in magenta. Dashed black lines indicate significant *P*-value (horizontal line) and log_2_ FC (vertical lines) thresholds. **(F)** Comparison of differentially activated hallmark and drug pathways across the mtDNA variants that were predicted to be functionally impactful, contrasted with data from a cybrid series harboring progressive increases of the pathogenic m.3243A>G mutation^24^. Upregulation (red) and downregulation (blue) of pathways depended on mtDNA mutation context. For example, upregulation of glucocorticoid (e.g., prednisolone) resistance (dark pink) was most prominent in leukemic blasts harboring the m.11889G>A mutation, and vincristine sensitivity (magenta) was most prominent in blasts carrying the m.15657T>A mutation and in cybrids carrying the m.3243A>G mutation at VAFs >0.6. **Abbreviations**: B-ALL, B-cell acute lymphoblastic leukemia; DA, differential activity; DN, downregulated; E2F, E2F transcription factor 1; FDR, false discovery rate; G2M, gap 2 phase/mitosis; GO, gene ontology; H, hallmark; IL6, interleukin 6; JAK, Janus kinase; Kras, Kirsten rat sarcoma virus; mtDNA, mitochondrial DNA; MTORC1, mechanistic target of rapamycin (mTOR) complex 1; NetBID, network-based Bayesian inference of drivers; R, reactome; scRNAseq, single-cell RNA sequencing; STAT3, signal transducer and activator of transcription 3; TNFa, tumor necrosis factor alpha; TU, tumor-enriched; UMI, unique molecular identifier; UP, upregulated; UV, ultraviolet; VAF, variant allele fraction; WT, wild-type.

In accordance with our single-cell gene expression studies, cells harboring the m.11889G>A mutation had the most extensive portfolio of altered gene activities, many of which would not have been predicted based on gene expression alone (**Figure 5B-D**, **Figure S6A** and **Table S2L**). We observed significant mtDNA mutation–associated and VAF-dependent increases in the DA scores of many drivers (PPA1, TEAD4, ACOT7, NPM1, AHCY, and PRDX6). GABRR1, a γ-aminobutyric acid (GABA) rho-subunit receptor implicated in regulating hematopoiesis^118,119^, was among the drivers with significantly reduced activity (**Figure 5B** and **Figure S6B**). Many genes whose activities were altered in cells harboring the m.11889G>A mutation (e.g., *ACOT7*, *AHCY*, *NPM1, GABRR1*) were also dysregulated in cells harboring the m.15657 T>A mutation (E2A025), but in the opposite directions (**Figure 5C**). Similarly, there were several genes (e.g., *FUT9*, *KL*) whose activities were differentially altered in cells with the m.8172 G>A (E2A037 sample, downregulated) or m.15657 T>A (upregulated) mutations (**Figure 5C-D**). *KL* encodes the single-pass transmembrane protein Klotho, which inhibits the PI3K–AKT pathway, among others^120^. Additionally, Klotho protects normal cells from oxidative stress but alters the metabolism of cancer cells (i.e., it inhibits glycolysis and reduces mitochondrial activity and membrane potential), thereby inhibiting their growth^121^. Klotho also increases the efficacy of certain types of chemotherapy^122^. Thus, changes in Klotho activity associated with certain TU mtDNA mutations may influence the growth and/or response to chemotherapy of subpopulations of cells.

We next performed gene set/pathway enrichment analyses on driver genes associated with the relevant TU mtDNA mutations (i.e., m.8172G>A, m.11889G>A, m.15657T>A, **Table S2M**). As a point of comparison, we also performed gene set/pathway enrichment analyses on driver genes associated with m.3243A>G mutation in cybrids with either increasing levels of heteroplasmy or lacking mtDNA (Rho0) (**Table S2N**). Results from the m.11889G>A mutation analyses suggested significant (*P* <10^-5^) dysregulation of numerous cellular programs, including the upregulation of gene networks associated with mitosis (AURKB targets)^123^, proliferation (MYC targets V1)^124^, cytokinesis^125,126^, and protein synthesis (cytosolic tRNA aminoacylation)^127–129^ (**Figure 5E** and **Tables S2M, S2O-P**). Many of the constituent members of these networks are components of the DREAM complex, a master regulator of cell cycle progression (DREAM targets)^130^ that works with E2F-bound targets ^131^ to coordinate periodic expression of genes among cycling cells^132^. In addition to the deregulation of cell cycle genes, the profile of gene activities associated with the m.11889G>A mutation had features similar to those typically observed in aggressive malignancies (e.g., thyroid cancer poor survival [UP, upregulated genes])^133^, and those with poor response to therapy (vincristine-resistant B-ALL [DN, downregulated genes]^134^, ALL glucocorticoid therapy DN^135^). Notably, many of the most differentially activated drivers in cells harboring the m.11889G>A mutation were members of the ALL glucocorticoid therapy DN gene set^135^ (**FigureS6C**), comprising genes that are typically downregulated in ALL samples in response to glucocorticoid treatment but are upregulated in glucocorticoid-resistant blasts (**Supplementary Note 10** and **Tables S2Q-R**).

A comparison of selected pathway analysis results revealed that the enrichment of differentially activated drivers involved in glucocorticoid resistance was not a universal feature of the variants. We found only a modest upregulation in cells with the m.8172G>A variant (E2A037) and downregulation in cells with the m.15657T>A variant (E2A025) (**Figure 5F**, **Table S2M**, and **Supplementary Note 10**). Similarly, robust induction of mTORC1 (mTOR complex 1) signaling and associated anabolic programs (fatty acid metabolism, amino acid activation, tRNA acetylation) were more strongly associated with the *MT-ND4* variant than the other two variants (**Figure 5E-F**, **Table S2M**). MYC target genes, including those involved in glycolysis, were also enriched in blasts harboring either the m.11889G>A (*MT-ND4*) or m.8172G>A (*MT-CO2*) variant; however, only the *MT-ND4* mutation demonstrated a simultaneous increase in OXPHOS. Notably, the simultaneous activation of mTOR, MYC, OXPHOS, and glycolysis were also features of the proliferative metabolic phenotype seen in ALL cell lines and patients whose disease was characterized as glucocorticoid resistant^136^. Downregulation of innate immunoregulatory pathways and upregulation of pathways related to growth and proliferation were also associated with the m.11889G>A (*MT-ND4*) and m.8172G>A (*MT-CO2*) variants, but those pathways were inversely affected by the m.15657 T>A (*MT-CYB*) variant. Another notable difference associated exclusively with the m.15657 T>A variant was the upregulation of genes typically downregulated in B-ALL samples resistant to vincristine.

Together, these findings highlight potential differences in cellular responses (including pathways involved in cell proliferation, innate immunity, and sensitivity to chemotherapy) associated with the TU mtDNA mutations at physiologically relevant levels of heteroplasmy. Although the TU mutations were each associated with a unique constellation of changes, the magnitude and types of changes associated with these mtDNA mutations were comparable to those associated with the m.3243A>G in cybrids with 60%-90% heteroplasmy (**Figure 5F**).

### Characterization of TU mtDNA variants in relapsed ALL

Considering that certain somatic mtDNA variants underwent expansion in leukemic blasts, co-segregated with distinct subclonal leukemia genomes, and exerted changes in transcriptional programs that may support oncogenic processes and/or alter sensitivity to therapy, we next examined the fate of somatic mtDNA mutations in the context of recurrent acute leukemia. Towards this end, we collated WGS data from 123 matching diagnosis, relapse, and germline (i.e., tumor-free) samples obtained from two published cohorts of pediatric patients with ALL (Shanghai Children’s Medical Center ALL-2005 frontline treatment protocol^137^ and St. Jude Total Therapy studies XI-XVI^138^). We used a modified analysis pipeline that integrated information across trios (Methods and **Table S1F**) and stringent QC processes that eliminated 35 samples, resulting in a total of 3,031 mtDNA SNVs detected among the remaining 88 (70 B-ALL and 17 T-ALL) relapsed sample trios (**Table S1G**). Of these, we identified 108 TU mutations (i.e., present in the diagnosis and/or relapse samples but not in the germline sample) in 56 (64%) of the trios.

TU SNVs were further stratified according to whether variants were present only at diagnosis (“lost” in relapse), were maintained from diagnosis to relapse (“persistent”), or were present only at relapse (“gained”). Based on this classification schema, ∼60% of TU SNVs were lost (**Figure 6A**), and most (63%) were nonsynonymous mutations (**Table S1G**). Although the VAFs of the lost TU SNVs were often lower than the persistent TU SNVs (**Figure 6B**), the higher-than-expected dN/dS among lost SNVs (**Figure 6C**) suggests that these mtDNA mutations not only mark a minor clone with increased sensitivity to chemotherapy but also have the potential to contribute to the treatment response^139–141^. Among the lost variants, we also found a m.15657T>C mutation, which results in the substitution of the hydrophobic isoleucine at position 304 of MT-CYB with a polar amino acid (threonine) (**Figure 6D**). Based on our single-cell sequencing data, a similar mutation (m.15657T>A) resulting in the substitution of MT-CYB I304 with a polar amino acid (asparagine) was associated with a signature predictive of vincristine sensitivity^134^ (**Figure 5F**).

**Figure 6.**
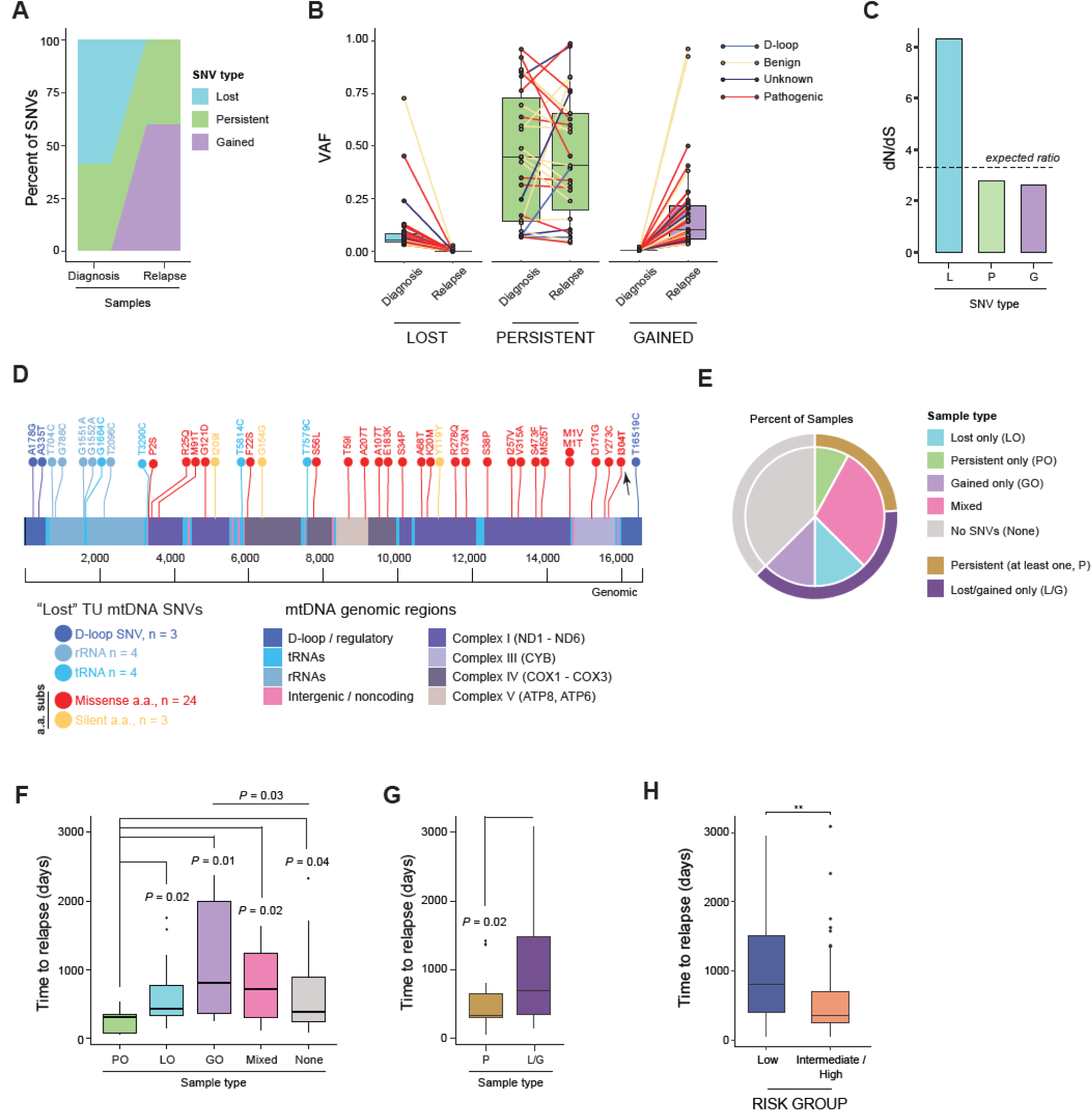
Characterization of mtDNA single-nucleotide variants in samples of relapsed acute lymphoblastic leukemia. Distribution and characteristics of 108 TU mtDNA SNVs, as determined by WGS of 88 matching diagnosis, relapse, and germline samples from two published cohorts of pediatric patients with ALL^137,138^. **(A)** The proportion and trajectory of TU mtDNA SNVs that persisted (green) from diagnosis to relapse or were lost (blue) or gained (purple) at relapse. **(B)** Paired boxplot illustrating changes in the mtDNA SNV variant allele fraction (VAF) from diagnosis to relapse among the variants that were either “lost” at relapse (blue), persisted from diagnosis to relapse (green), or were “gained” at relapse (purple). Variant trajectories are colored according to their predicted impact. **(C)** Barplot illustrating differences in the observed ratio of nonsynonymous-to-synonymous (dN/dS) substitutions among TU mtDNA mutations that are lost, gained, or persist at relapse. The expected dN/dS (3.3) for neutrality after considering the mtDNA mutational signature is denoted by a dashed line. For lost mutations, the bootstrapped *P*-value = 0.0708. **(D)** ProteinPaint^209^ depiction of the mitochondrial genome annotated with TU mutations lost at relapse (see color key). Notably, most lost mutations were nonsynonymous (in red), including the MT-CYB I304T substitution (arrow), which is predicted to be similar, in terms of structural consequences, to the I304N substitution in E2A025 from the discovery PCGP cohort. D-loop/regulatory, tRNA, and rRNA mutations are annotated as SNVs; all other mutations are annotated using amino acid alteration nomenclature. **(E)** Nested pie chart illustrating the proportion of samples with the indicated type(s) of TU mtDNA SNVs. Outer arc: samples with at least one TU mtDNA SNV (i.e., persistent, lost, and/or gained) were classified as harboring at least one persistent mtDNA SNV (gold) or only SNVs that were lost or gained (i.e., no persistent mtDNA SNVs, dark purple). Inner arc: samples with at least one TU mtDNA SNV were further stratified based on the number and type of SNVs. Samples with only persistent (green), lost (blue), or gained (purple) mtDNA SNVs are distinguished from those harboring multiple mtDNA SNVs of different classes, which were denoted as mixed (pink). Samples with no TU mtDNA mutations are depicted in gray. **(F)** Boxplots comparing median time to relapse among patients whose samples were categorized as in the inner arc of panel **e**. Patients whose samples harbored exclusively persistent (green) mtDNA SNVs relapsed sooner than all other groups. Comparisons were made using two-sided Wilcoxon signed-rank tests. **(G)** Boxplots comparing the median time to relapse between patients whose samples had at least one persistent mtDNA SNV (gold) and those harboring only lost or gained SNVs (dark purple). Median time to relapse was significantly shorter among samples with at least one persistent SNV (307 *vs.* 528 days, two-sided Wilcoxon signed-rank, *P* = 0.02). **(H)** Boxplot indicating that leukemia risk group is an independent predictor of time to relapse (in days). Comparisons were made using a two-sided Wilcoxon ranked sum test, **P ≤ 0.01. **Abbreviations:** a.a., amino acid; ALL, acute lymphoblastic leukemia; ATP, adenosine triphosphate; COX, cytochrome c oxidase; GO, gained only; LO, lost only; MT-CYB, mitochondrially-encoded cytochrome b; mtDNA, mitochondrial DNA; ND, NADH: ubiquinone oxidoreductase; PO, persistent only; rRNA, ribosomal RNA; SNV, single-nucleotide variant; tRNA, transfer RNA TU, tumor-enriched; WGS, whole-genome sequencing.

Next, patients were classified based on the composition of the TU variants (if any) that they harbored (**Figure 6E**) and whether there were associations between these classes and time to relapse (TTR) (**Table S1G**). The TTR was significantly shorter among patients who harbored only persistent mutations, as compared to those who had only lost mtDNA SNVs (median 307 *vs.* 436 days; *P* = 0.023; Wilcoxon rank-sum test), those who only gained mtDNA SNVs (median 307 *vs,* 814 days; *P* = 0.006; Wilcoxon rank-sum test), or harbored a mixture of TU variants (median 307 vs 714 days; *P* = 0.023; Wilcoxon rank-sum test) (**Figure 6F**). Similarly, we found that the TTR among patients who possessed at least one persistent TU SNV was significantly shorter than that of patients harboring only lost or gained TU SNVs (median 307 *vs.* 528 days, *P* = 0.01; Wilcoxon rank-sum test) (**Figure 6G**). Patients were also categorized into 3 groups according to their clinical risk, which was based on molecular subtyping^142,143^: ETV6–RUNX1, hyperdiploid, and *DUX4r* B-ALL were considered low risk, and all other disease subtypes were considered intermediate/high risk. Although subtype risk group (low *vs.* intermediate/high) was an independent predictor of TTR (*P* <0.01; Wilcoxon rank-sum test) (**Figure 6H**), we detected no interaction between TU variant type and B-ALL risk group (*P* = 0.520; two-way ANOVA). Together, these analyses suggest that certain pathogenic mtDNA mutations present at diagnosis undergo negative selection and are eliminated during relapse, while others that persist may simply serve as biomarkers of relapse.

## Discussion

Large-scale sequencing studies have facilitated the improved detection of somatic mtDNA mutations across diverse tumor subtypes. Despite the growing body of evidence supporting oncogene-driven changes in mitochondrial metabolism as drivers of disease, the oncology field has struggled to define the contribution of somatic mtDNA mutations to cancer pathophysiology. Here, we address this gap in knowledge by presenting conceptual and methodological advances that elucidate the level and functionality of somatic mtDNA mutations in pediatric ALL. Our approach involved the development of new pipelines for analyzing data from existing WGS datasets and from our own exploratory single-cell sequencing experiments.

First, we demonstrated that pediatric cancers are enriched with mtDNA variants that are distinct from other *de novo* mtDNA variants that arise during normal development. Through a comprehensive investigation of population variation, conservation scores, and predicted functional impact, we found that TU variants are rare in the general population and are more likely than other *de novo* variants to disrupt protein function. This observation aligns with prior data showing that although mutational origins may be equivalent, pathogenic variants in tumor cells are not subject to the same purifying selective pressure that operates in nontumor tissues and contexts (i.e., relaxation of negative selection)^9,37–39,144^.

Second, our initial analyses of bulk sequencing data from pediatric malignancies indicated that pathogenic mtDNA SNVs tend to exist at relatively low VAFs, consistent with previous reports^9,10,40,41^. However, unlike in adult malignancies, where nonsynonymous and synonymous variants showed similar cumulative VAF distribution patterns, nonsynonymous variants in pediatric malignancies accrued with greater restriction than their synonymous counterparts. It is plausible that significant disruption of OXPHOS in malignant cells harboring high levels of a pathogenic mtDNA mutation impedes their proliferation^43,145,146^, thereby limiting variant accrual in bulk WGS assays. Indeed, the application of Mitovolve to data from our single-cell sequencing studies, which provide insights into VAF heterogeneity not evident through bulk WGS, suggests that functionally impactful mutations can accumulate to intermediate levels before undergoing negative selection at high VAFs. This pleiotropic relationship between selection pressure and VAF is not surprising given that mtDNA mutation-associated alterations in metabolism, transcription and epigenetics also vary in a VAF-dependent manner^24,29,59,147^. Moreover, the positive correlation between the degree of selection and the associated gene expression changes suggests that Mitovolve may be used to distinguish functionally impactful variants from those that are inert.

Our results demonstrate the utility of combining scRNAseq with systems-based computational biology approaches (e.g., NetBID) to identify cell-state changes associated with somatic mtDNA mutations in primary cells. For example, we identified mtDNA mutation-specific changes in the activities of pathways related to mTORC1 signaling and energy metabolism (e.g., glycolysis, fatty acid metabolism, OXPHOS). As a master regulator, mTORC1 integrates environmental cues with the metabolic needs of the cell^148^, thereby serving as a molecular fulcrum that shifts metabolic activities in response to changing bioenergetic and stress-related stimuli. Alterations in mitochondrial function are sufficient to provoke such shifts^149–152^; however, their impact on mTORC1 signaling appears to be contingent upon the degree and/or mode of disruption of OXPHOS, as well as the cellular context. For example, pharmaceutical inhibition of ETC components or ATP synthase impairs mTORC1 signaling in cell lines^149,151^, whereas mtDNA deletions in a mouse model of mitochondrial myopathy hyperactivate mTORC1 *in vivo*^150^. In patient-derived cells and muscle biopsies harboring the pathogenic m.3243A>G mutation, increases in PI3K–Akt–mTORC1 pathway activity have been coupled to changes in metabolite composition associated with the mutation^152^. Given that mtDNA integrity and ETC fitness are important determinants of mTORC1 function, it is not surprising that the three TU mtDNA mutations of interest were all associated with changes in mTORC1 activity. Similarly, as MYC is involved in retrograde-signaling pathways, linking mitochondrial function with the expression of genes involved in growth, metabolism, and mitochondrial biogenesis^153–155^, the observed mtDNA mutation–associated changes in MYC activity are consistent with alterations in mitochondrial function in leukemic cells bearing mtDNA mutations. Only the m.11889G>A variant was associated with dual induction of both OXPHOS and glycolytic programs. This metabolic reprogramming, in conjunction with the increased MTORC1 and MYC activities, may help to explain the persistence of this mutation at intermediate VAFs ^136,156,157^.

Notably, we observed that the direction and magnitude of expression changes in many common genes and associated pathways were inconsistent across the variants studied. Although all genes encoded by the mitochondrial genome ultimately impact OXPHOS, these results suggest that different mutations and the resulting changes in the structure of mtDNA gene products can elicit distinct types of cellular responses. While this variability may be attributed to many causes (e.g., the specific effects of different mutations on mitochondrial gene products, varying levels of heteroplasmy, distinct compensatory mechanisms activated by cells, interactions between mitochondrial and nuclear genomes, and/or differing levels of oxidative stress and ROS production), investigations of additional mutations may help to identify groups of mtDNA mutations that are associated with similar changes in cell state, which may enable functional categorization of mtDNA mutations.

Therapeutic resistance, disease recurrence, and poor prognosis are commonly attributed to subclonal variation^158^. Understanding clonal evolution may depend on our ability to reconstruct the subclonal acquisition of nuclear mutations; however, recent studies^45–47^ have suggested using mitochondrial SNVs as stable markers for clonal progression. Here, through sc-mtDNAseq of a sample with established clonal architecture^76^, we determined the clonal trajectory of a TU mtDNA mutation (m.15171G>A in *MT-CYB*) and not only confirmed its early appearance in the ancestral clone but also found evidence for its propagation in one of the two clonal derivatives. Given that our scRNAseq studies revealed cell-state changes associated with pathogenic TU mtDNA mutations in primary leukemic blasts and considering that the m.15171G>A mutation occurs at a highly conserved residue that might hinder complex III function, the early enrichment and subsequent loss of this mutation in subclones with distinct leukemia genomes suggests that the mtDNA mutation (1) contributed to the evolution of the clones and/or (2) affected the fitness of clones throughout therapy and disease progression. Therefore, somatic mtDNA mutations may not be merely markers of clonal progression but may help shape the divergence of clones (**Figure 7A-C**).

**Figure 7.**
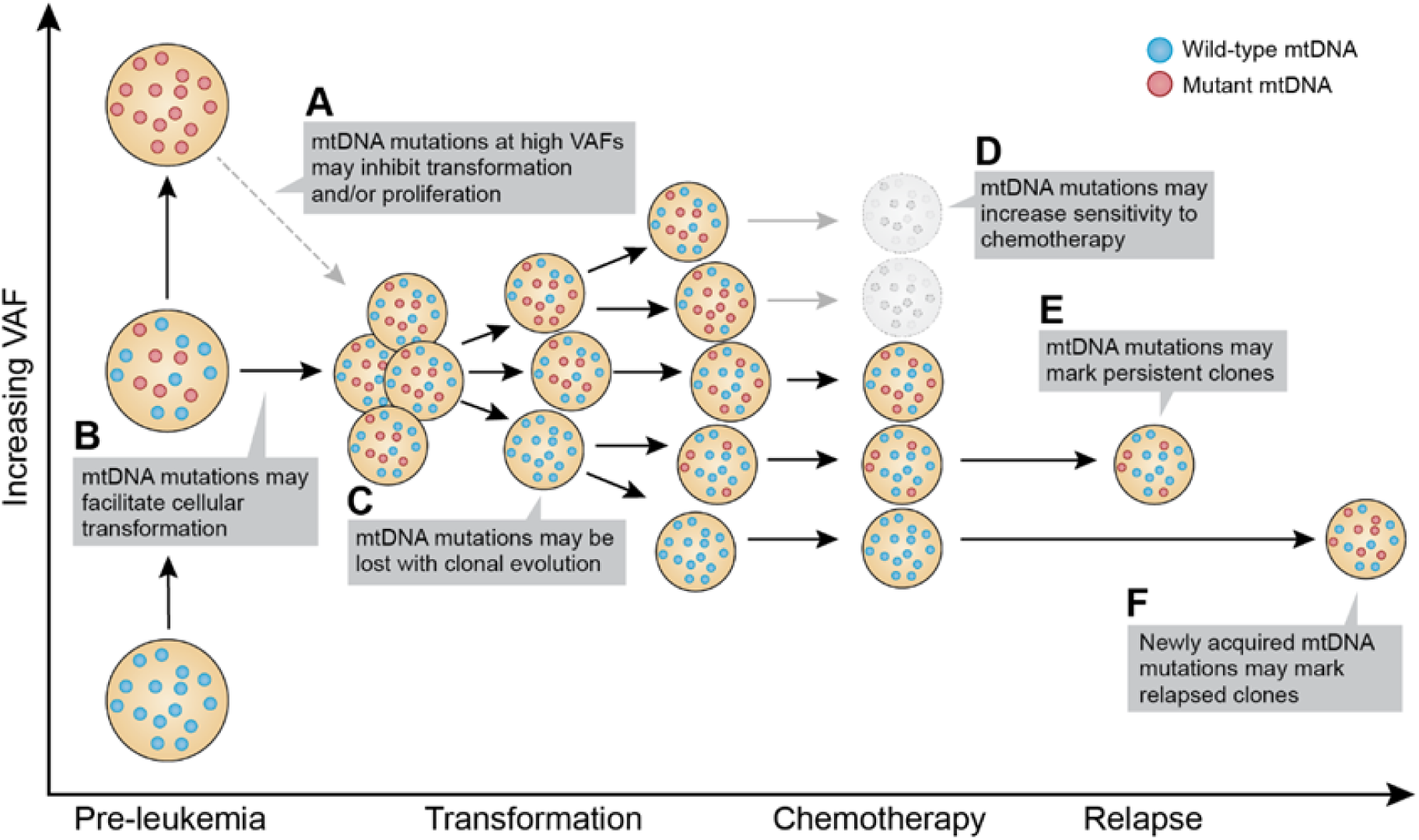
Contributions of somatic mtDNA mutations fluctuate throughout pediatric leukemia progression. **(A)** Homoplasmic or near-homoplasmic levels of pathogenic somatic mtDNA mutations may disrupt mitochondrial bioenergetic/biosynthetic functions in a manner that is prohibitive of enhanced self-renewal, transformation, and/or clonal expansion of leukemia-initiating stem cells. This may help explain the decreased representation of leukemic cells harboring high levels of mutated mtDNA. **(B)** In contrast, intermediate levels of pathogenic somatic mtDNA mutations may render pre-leukemic hematopoietic stem cells more permissive to transformation by specific oncogenes. Founding oncogenic translocations and mutations in nuclear DNA–encoded oncogenes or tumor suppressors can alter metabolic homeostasis within pre-leukemic hematopoietic stem cells, thus requiring mitochondrial adaptation. Clones with intermediate levels or certain mtDNA mutations may be metabolically rewired to rapidly replenish amino acids, fatty acids, nucleotides, metabolites, and energy substrates in response to mTORC1 and MYC signaling and may even stimulate these pathways through metabolic signaling (e.g., ATP, reactive oxygen species, NAD+, FAD). Thus, pre-leukemic hematopoietic stem cells exposed to intermediate levels of functionally impactful mtDNA mutations may adapt to oncogenic stimuli more effectively than do cells without such mutations (or those with near-homoplasmic levels of somatic mtDNA mutations). **(C-F)** In leukemia-initiating stem cells, the differential acquisition of mutations in nuclear DNA–encoded genes and their derivatives often results in an assortment of diverse clones at diagnosis, each with a distinct leukemic genome. These clones compete for fitness at various stages of disease progression and throughout treatment. Therefore, depending on the functional alteration and context, mtDNA mutations that were advantageous early in disease may be selected against later in disease [e.g., the acquisition of additional mutations in nuclear DNA–encoded genes **(C)** or upon exposure to certain chemotherapeutic agents **(D)**]. Other somatic mtDNA mutations may become inconsequential and serve as markers of resistant clones **(E,F)**.

In addition, certain TU mtDNA mutations (perhaps a different subset than those promoting leukemogenesis) may sensitize leukemic cells to chemotherapy (**Figure 7D**). In a study that investigated the subclonal properties of diagnostic samples collected from patients with relapsed B-ALL, Dobson et al.^159^ discovered that, at diagnosis, relapse-fated subclones are uniquely poised to survive chemotherapy based on transcriptional alterations related to mTOR activation, stress response, and mitochondrial metabolism. Although these same pathways appeared to be upregulated in blasts harboring the m.11889G>A mutation, which were predicted to be less sensitive to glucocorticoids than their WT counterparts, the small degree of change in prednisone sensitivity may not be sufficient to influence disease outcome. In contrast, a mutation at position m.15657T>A (E2A025) was predicted to sensitize cells to vincristine; a similar mutation (m.15657T>C, also causing the replacement of I304 in MT-CYB by a polar amino acid) was among the lost variants in the relapse cohort. The overall decrease in synonymity (elevated dN/dS) among lost variants, compared with that of persistent or gained variants, further supports the notion that certain somatic mtDNA mutations sensitize cells to chemotherapy (**Figure 7A-D**).

Although the results of dN/dS analyses argue against a major role for persistent or gained variants, the TTR study results suggest that they are useful as markers of the relapsed clone (**Figure 7E-F**). Additional studies of larger patient cohorts and samples will be needed to determine whether there are common cell-state changes associated with lost mtDNA mutations, compared with those of persistent or gained mutations. In particular, the repeated occurrence of somatic mtDNA mutations impacting the I304 residue among four separate leukemia samples warrants further investigation.

While it is commonly believed that critical determinants of tumor pathophysiology reside primarily within the nuclear genome, we anticipate that single-cell approaches, such as those described here, will help shift the focus to the mitochondrial genome. These methods provide a means to stratify mtDNA variants based on evidence of selection and associated cell-state changes, thereby streamlining strategies to assess their contributions in multiple disease settings.

## Methods

### Patient samples and datasets

WGS datasets for 637 diagnostic pediatric tumors and their matched germline samples, along with 307 matched RNAseq datasets, were extracted from the PCGP^48^. Datasets from 123 WGS relapsed pediatric ALL samples were collated from 102 Chinese genomes provided by Li et al.^137^ and from 21 PCGP genomes provided by Waanders et al.^138^. The prednisolone-sensitivity prediction (PSP) score analysis utilized RNAseq datasets from 161 pediatric patients with newly diagnosed B-ALL (discovery cohort) and 179 adult and pediatric patients with B-ALL (validation cohort) provided by Autry et al^160^. Study approval was granted by the St. Jude Institutional Review Board, and informed consent was obtained from the parents/guardians of each child.

### mtDNA WGS analysis

WGS reads were aligned to the human reference genome build 37 (GRCh37)^161^ and to the revised human mitochondrial reference (NC_012920)^162^ using Burrows-Wheeler Aligner (BWA) v.0.7.12^163^. Sequencing reads and depths were calculated using SAMtools v.1.2 (http://samtools.sourceforge.net/)^164^. Due to the exceedingly high number of mtDNA molecules in each cell, the average mtDNA-sequencing depth (7,204×) was considerably higher than the typical 30× nuclear genome coverage, which afforded increased confidence in calling lower VAFs. Furthermore, WGS coverage availed estimates of the mtDNAcn, which were calculated using mitoCalc^49^.

### mtDNA variant detection and classification

To align sequence reads, an established “double-alignment” algorithm^49^ was used to compensate for the circular nature of the mitochondrial genome. Variants with >1 read supporting the alternate allele were extracted using Bambino^165^, followed by elimination of variants with <100 read coverage, variants that were the product of strand bias (>90% of the mutant-allele–supporting reads from only 1 strand) and variants in which <10 reads supported the mutant allele. Variants were then classified according to the following criteria (tVAF, tumor VAF; gVAF, germline VAF):

- **INH** (inherited): tVAF ≥0.97 AND gVAF ≥0.97
- **TU** (tumor-enriched): (tVAF ≥0.03 AND gVAF <0.01) OR (tVAF/gVAF >3)
- **GH** (germline heteroplasmy): (gVAF ≥0.03AND tVAF <0.03) OR (tVAF ≥0.97 AND gVAF <0.97 AND tVAF/gVAF ≤0.03)
- **SH** (shared heteroplasmy): all other SNVs

SNVs in which both the germline and the tumor VAFs were <0.03 were excluded from further analyses.

Relapse samples were subjected to the same pipeline (Supplementary Fig. 14) as the PCGP-paired tumor–germline samples but with an additional paired run (i.e., relapse–germline). The same variant-detection algorithm was also used, with the following modified classifications to account for the relapse samples (rVAF, relapse VAF):

- **INH** (inherited): tVAF ≥0.97 AND rVAF ≥0. 97 AND gVAF ≥0.97
- **RP** (TU, persistent): (tVAF ≥0.03 AND rVAF ≥0.03 AND gVAF <0.01) OR (tVAF/gVAF >3 AND rVAF ≥0.03)
- **RL** (TU, lost): (tVAF ≥0.03 AND rVAF <0.03 AND gVAF <0.01) OR (tVAF/gVAF >3 AND rVAF <0.03)
- **RO** (relapse-only): tVAF <0.03 AND rVAF ≥0.03 AND gVAF <0.01
- **GH** (germline-specific): tVAF ≤0.03 AND rVAF ≤0.03 AND gVAF ≥0.03
- **SH** (shared): all other SNVs

### Elimination of false positives

Although the PCGP dataset has been carefully curated^166^, we supplemented our pipeline with several metrics to minimize the likelihood of calling false positives. First, the minor cross-contamination (C-value) was estimated in a manner like that of Ju et al.^10^, in that the VAF of autosomal-homozygous SNPs genotyped from common SNP sites was used to gauge the level of adulteration. On the basis of this criterion, we removed seven samples that had >2% autosomal contamination in the tumor (SJE2A021, SJINF020032, SJNBL006, SJE2A064, SJNBL039, SJNBL021) or the germline (SJCBF013). We also disposed of known false-positive variants that were symptomatic of either misalignment triggered by mtDNA homopolymers or common sequencing errors (A302C, C308T, C309A, C309T, T310C, C314T, G513A, C516T, A517T, A567C, C3106G, A16183C, T16189C, T16189A, C16192T). Well-characterized germline polymorphisms were used to identify false somatic substitutions (SJBALL020625) and detect germline back-mutations, both of which are also indicative of contamination. Thirteen samples had ≥2 germline back-mutations and were removed (SJMB031, SJBALL021170, SJHYPER084, SJPHALL020040, SJERG020051, SJHYPER119, SJE2A006, SJHYPER010, SJBALL021058, SJBALL021516, SJBALL021893, SJHYPER123, SJHYPER095). Finally, we used C-values to calibrate VAFs and eliminated SNVs with an adjusted tumor VAF or an adjusted germline VAF <0.03.

To test the ability of our pipeline to detect true mtDNA variants with confidence while filtering out potential background signal arising from nuclear mtDNA sequences^50^, we performed WGS on a human osteosarcoma cell line (143B) and its mtDNA-deficient derivative Rho0 (or ρ°)^167^. The overall coverage depth across the nuclear genome was comparable between the two cell lines (∼37× and ∼39×, respectively), yet none of the 33 mtDNA variants detected in the parental cell line were identified in the Rho0 sample.

We used a simulation to evaluate the impact of tumor purity on our pipeline’s ability to detect TU variants. As a point of reference using the PCGP dataset^48^, acute leukemia samples consisted of at least 70% blasts (i.e., leukemic cells); tumor purity exceeded 50% in brain tumor samples and ranged from 48% to 96% in solid tumor samples. First, we generated a grid of theoretical coverage depth (100×-10,000×) with a step size of 100 and variant allele read support for both tumor and germline samples. For tumor samples, the grid was generated using the full range of VAFs (0.01-1.0) with a step size of 0.01. For the germline samples, since the germline VAFs for >92% of the TU SNVs was <0.02, the grid was generated using integer-based alternative read counts as large as the number supporting VAFs of 0.02. We then applied our pipeline to the 101 million theoretically possible combinations of tumor and germline read counts. Assuming a model of 100% tumor purity, we found that 97 million combinations passed our pipeline’s detection for TU mutations.

Next, we generated new grids for the tumor samples to model a range of tumor purity percentages (50%-99%) using a step size of 1% and adjusting the tumor VAF at each step. The simulated data were then submitted through our pipeline. With each 1% decrease in tumor purity, less than 0.1% of all possible simulated combinations were lost. Mutations with tumor VAFs close to 0.03, which were at the lower bound of our filtering threshold, were the ones most likely to be lost as tumor purity decreased. Overall, the results suggested that the main impact of low tumor purity is a small decrease in our ability to detect TU variants at thresholds near our VAF cutoff of 0.03. Similarly, a small percentage of variants classified as GH might be SH variants in cases with low tumor purity.

### RNAseq pipeline validation

Tumor VAF estimates from WGS were validated using matched RNAseq data from 307 pediatric tumor samples. For mitochondrial variants detected by WGS, the corresponding supporting reads for the reference and mutant alleles in the RNAseq BAM files were extracted to compute the VAF. Reads flagged as optical or PCR duplicates and those with a base quality score <15 were excluded from the estimation of the VAF. Samples with VAF differences >0.99 in one or more variants were also eliminated (SJRHB012, SJMEL001004, SJTALL002, SJTALL012).

### Variant annotation

Successful candidate SNVs were annotated with population-frequency tabulations, and predicted impact scores were based on information from databases of mtDNA common polymorphisms^168,169^, MITOMAP^52^ (accessed July 11, 2018), and Mutation Assessor (v3.0)^170^. For simplicity, all synonymous and neutral-impact variants were classified as “predicted benign,” whereas all other protein-encoding sequence variants were classified as “predicted pathogenic.” MitoTIP^171^ was used to predict the pathogenicity of mitochondrial tRNA variants, and rRNA variants were assessed using heterologous inferential analysis (HIA)^172,173^. ANNOVAR^174^ was used to append location and functional annotation (nonsynonymous, synonymous, stop gain/loss) to each variant and to translate amino acid changes for all coding nonsynonymous SNVs. Any variants occurring in intergenic regions or lacking information from the above sources were categorized as having an unknown predicted impact. Conservation scores were determined based on the percentage of residues that matched the rCRS derived from all species and are provided for all protein-coding, tRNA, and rRNA variant loci.

### Bulk nuclear DNA variant calls and detection for correlational analyses

WGS data were processed and analyzed according to previous methods^48^, and somatic nuclear SNVs and structural variants were identified and sorted into tiers as previously described^175,176^.

### Mutational signature and recurrence

Mutational signature comparisons between TU mitochondrial and nuclear genomes were based on a suite of COSMIC mutational signatures developed by Alexandrov et al.^177^. For each SNV class, strand-specific substitution rates were derived from tabulating the occurrence of 192 possible trinucleotide contexts (96 each on the L-strand and the reverse-complemented H-strand) among SNVs. Assuming an equal probability of allelic substitutions, the expected value of each triad was calculated based on its frequency in rCRS^162^, and its substitution rate was determined by the number of observed/expected mutations. Comparisons between TCGA and PCGP TU mutational signatures and signature comparisons among pediatric SNV classes were tested by cosine similarity^178^. Because different patients harbor varying numbers of mutations, the individual probability of a mutation at a given genetic locus will also vary by individual. Therefore, we used GRIN analysis^179^ to adjust for this variation and determined the *P*-value by computing the probability distribution for the number of individuals with a mutation in each given region by chance.

### scDNA-seq of primary leukemic blasts

Based on previously developed protocols^76^, primary leukemic blasts were isolated from the bone marrow of two pediatric ALL cases (INF010, ETV027); each was processed separately and subjected to individual cell-capture and multiple displacement-amplifications using the C1 Single-Cell Auto Prep System (Fluidigm). Briefly, individually captured primary leukemic blasts were counted, evaluated for singlets, and assessed for viability (on chip LIVE/DEAD stain, Invitrogen) using phase contrast and GFP/Y3 fluorescence filters on a Leica microscope. On the Integrated Fluidic Circuit (IFC) chip, cells were then lysed and subjected to whole-genome amplification (REPLI-g Single Cell Kit, Qiagen), followed by enrichment of mitochondrial genomes with a pool of custom-capture oligos (**Table S2Q**) purchased from IDT.

Bulk DNA from all three samples were also re-sequenced with the same strategies used to interrogate single cells. Libraries from enriched mitochondrial genomes were constructed using the KAPA HyperPlus kit (Roche). All libraries were then subjected to 2 × 150-bp paired-end sequencing on a MiSeq (Illumina). For ETV027, mtDNA variants were then connected to the clonal nuclear variants identified in those cells^76^. Sequenced data were then subjected to quality trimming and adapter removal using Trimmomatic^180^. Using BWA v.0.7.12^163^, reads were subsequently aligned to hg19(GRCh37)^161^ and the revised human mitochondrial reference genome (NC_012920)^162^. Then they were sorted, compressed, and indexed using Picard (https://broadinstitute.github.io/picard/). Variants with >1 read supporting the alternate allele were extracted using Bambino^165^ and annotated with ANNOVAR^174^. We next focused on variants previously identified by WGS and excluded cells that either did not have an average of 10 reads per selected loci and coverage greater than 1.5× the upper boundary of the IQR (potential doublets).

### scRNAseq of primary leukemic cells

Viable single-cell suspensions from three pediatric E2A–PBX1^+^ ALL tumor samples were processed using the Chromium v.2 Single Cell 3′ Library and Gel Bead Kit, in conjunction with the 10X Genomics Chromium single-cell platform (10X Genomics). Libraries were paired-end sequenced (26-bp read 1, 8-bp I7 index, and 98-bp read 2 configuration) on an Illumina NovaSeq 6000 genome analyzer; 100-bp reads were aligned against the human genome (hg19) using the 10X Cell Ranger pipeline (ver. 1.2.0). QC, cell clustering, and expression analyses were enabled by the R Seurat toolkit 3.0.1^181^. Cells were retained for subsequent analyses (see also Supplementary Dataset 3) based on the following criteria: number of genes per cell was ≥200 and ≤6000, the fraction of mitochondria UMIs per cell was ≤0.5, and the minimal number of expressed cells (UMI >0) for each gene was 3. For example, the gene × cell after QC for each diagnostic leukemia sample were as follows: 14413 × 2074 (E2A037), 14002 × 1591(E2A025), and 15467 × 2622 (E2A015).

For cell clustering, we first removed mitochondrial and ribosomal genes and then normalized (*NormalizeData*) and scaled (*ScaleData*) the single-cell data. Principal component analysis (PCA) was then performed (*RunPCA*), and cell clusters were inferred (*FindClusters*) based on the normalized expression values and markers (*FindMarkers*) in each cluster. After identifying the cell types of each cluster, we merged clusters of blast cells into a single cluster and identified the top differentially expressed genes among different clusters (*FindMarkers*). The tSNE results (*RunTSNE*) were generated using the top 10 principal components.

Aligned BAM files were parsed to enumerate the number of reads containing reference or alternative nucleotides at each locus of interest (i.e., harboring a variant previously identified by WGS) by using the SAMtools pileup method implemented in Pysam (https://github.com/pysam-developers/pysam)^164^. The cell barcodes and UMIs were retrieved for each read by using the tag CB (cell-barcode) and UB (UMI-barcode), respectively. Low-quality bases/reads (i.e., base quality <15, mean read quality <15, and 10× adjusted mapping quality <255) were discarded, and only the reads in cells that passed Seurat QC were retained for VAF calculations.

### Differential expression analysis

For each TU mutation under investigation, cells were classified as mutant if the alternate UMI was ≥20%, whereas those with an alternate UMI = 0% and reference UMI counts ≥5 were classified as WT. After exclusion of potential doublets, differential expression (DE) analysis was performed using the widely accepted negative binomial distribution to model UMI counts^182–184^. Specifically, the *edgeR* package^185^ was used and parameter configurations were adjusted to accommodate scRNAseq data (e.g., likelihood ratio test; set prior.count = 0), while the *scran* package^186^ was utilized to estimate the library size^186^. Log_2_-fold change (log2FC) was reported to describe the effect size and the FDR was reported to account for multiple-comparisons testing. With regards to the association between m.8172G>A and *CHCHD2*, we tested for robustness by using the nonparametric Wilcoxon rank-sum test, which yielded a similar significance value (P = 3.8 × 10^-15^). To further confirm that the significance of the association was not by chance, we performed DE analysis on permuted VAF-based phenotypes. The permutation-based analysis was repeated 1,000 times, resulting in 12,859 × 1,000 p-values), and as a result, the *P*-value of *CHCHD2* from the observed data was ranked the second smallest among all 12,859,000 permutated tests (substantially less than *P*-value = 0.05).

### Mitovolve: inference of the evolutionary history of somatic mtDNA mutations

*Mitovolve* starts with an initiating parent cell with a given number of WT and mutant copies of mtDNA that, for simplicity, are doubled prior to division of the parent cell. Hypergeometric sampling of parent cells generates the partitioning of mitochondrial genomes into daughter cells, resulting in a probability distribution for the number of mutant mtDNA in the daughter cells. For each subsequent generation, the process is reiterated for a given number of generations to obtain a final probability distribution for the number of mutant mtDNA per cell. This theoretical probability distribution is defined by parameters representing the number of observed mtDNA UMIs per cell that yield an expected probability distribution, and results in a histogram representing the expected mutant allele fraction in the dataset.

To model this process with evolutionary selection, *Mitovolve* uses a noncentral hypergeometric distribution with the log odds ratio of selecting a mutant mtDNA defined as a cubic polynomial function

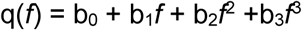

of the mutant allele frequency (MAF) *f*. To model the process without evolutionary pressure, *Mitovolve* uses a central hypergeometric distribution obtained by setting b_0_ = b_1_ = b_2_ = b_3_ = 0 in the above equation so that there is no preference for a mutant mtDNA over a WT mtDNA (the log odds ratio q(*f*) = 0). Positive values for q(*f*) indicate a selective preference for the mutant mtDNAs whereas negative values for q(*f*) indicate a selective preference for the WT mtDNAs.

*Mitovolve* uses a likelihood framework to evaluate the fit of a series of theoretical models to the observed distribution of WT and mutant mtDNA UMIs that cover a particular base-pair locus across individual cells from the single-cell sequencing data. The negative log likelihood (nLL) of a given model (i.e., based on initial VAF and generation number, with or without selection) is then computed by comparing the theoretical distributions of mutant and WT mtDNA genomes per cell (weighted based on UMIs) to the observed distribution. For a user-specified range of starting VAFs and generation numbers, *Mitovolve* uses a likelihood ratio test to compare the best nLL among models with selection to the best nLL among models without selection to determine whether there is statistically significant evidence that the mtDNA mutation was subject to selective pressure. *Mitovolve* also uses a series of likelihood ratio tests to find models with fits that are not significantly worse than that of the model with the best nLL.

### NetBID identification of drivers and pathways associated with pathogenic mtDNA mutations

To systematically infer drivers of both gene and protein expression from the scRNAseq data, we deployed NetBID (v.2.0, https://jyyulab.github.io/NetBID/)^114^. Single cells were first classified as either mutant or WT for each mtDNA mutation by using the criteria described above, with each variant considered separately. Within the NetBID environment, SJARACNe (v.0.2.1, https://github.com/jyyulab/SJARACNe)^115^ was utilized to construct the B-ALL interactome from an RNAseq dataset representing the transcriptomes of 185 pediatric B-ALL cases (TARGET cohort)^116^, integrate scRNAseq expression data from WT and mutant cells with the B-ALL interactome, and generate networks of transcription factors and signaling molecules, from which driver activity was inferred for each of the 7,890 hub genes in each cell using NetBID’s *cal.Activity* function. Bayesian linear analysis was then used to identify differentially expressed drivers between mutant and WT cells, at both the inferred gene expression (DE) and activity (DA) levels. The m.1200G>A SNV (in E2A015) and m.11865T>C SNV (in E2A037) were excluded from further analyses due to minimal effects on gene expression/activation (data not shown). STRING database analysis was used to initially characterize the biological implication of the top 50 significant DA drivers by using the normal gene set analysis settings and inclusion of all active interaction sources.

Pathway activity was computed (*cal.Activity.GS*) based on each cell’s inferred hub gene activities, with the activity of a pathway *P* in cell *c* was defined by the following equation:

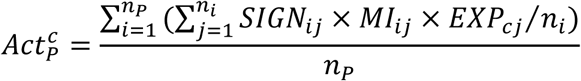

The gene expression matrix was z-normalized in each cell. The *n*_*p*_ and *n*_*i*_ are the number of hub genes in pathway *P* and the number of target genes of hub gene *i*, respectively. *EXP_cj_* is the expression value of gene *j* in cell *c*. *MI_ij_* is the mutual information between hub gene *i* and its target gene *j.sign_ij_* is the sign of Spearman correlation between gene *i* and its target gene *j*. After MSigDB (v7.4)^187^ gene sets were curated to exclude irrelevant positional (C1), computational (C4), and cell-type signature (C8) gene sets. Bayesian linear modeling was used to calculate differential pathway activity (*getDE.BID.2G*).

Independently curated gene sets from MSigDB database (collection H, C2, C3, C5, C6, C7) were also queried for enrichment of drivers by using significantly differentially activated drivers of variants m.8172G>A, m.11889G>A, and m.15657T>A. To include a similar number of top drivers of each variant in the analysis, different P-value cutoffs were used for the three variants (m.11889G>A: *P* <6.7 × 10^−6^; m.15657T>A: *P* <6.7 × 10^−6^; m.8172G>A: *P* ≤0.05). Gene sets with <30 or >500 genes were excluded from the analysis, and Fisher’s exact test (*funcEnrich.Fisher*) was used to evaluate statistical significance.

### Ex vivo prednisolone sensitivity in primary leukemia samples

Primary leukemia cells were isolated from the bone marrow or peripheral blood of patients with newly diagnosed ALL and tested for prednisolone sensitivity (Solu-Medrol, Pfizer) by 3-(4,5-dimethyl-2-thiazolyl)−2,5-diphenyl-tetrazolium bromide (MTT) assay, as previously described^160^.

### NetBID estimation of biomarker-based PSPs

Each patient’s RNAseq profile (log_2_-transformed FPKM) was first standardized by z-transformation so that comparisons could be made across samples. Activity scores were individually calculated for each of the 200 hub genes (top 100 upregulated hub genes and top 100 downregulated hub genes) by using the B-ALL–specific interactome and NetBID’s weighted mean function (*cal.activity*). For a patient *p*, we used a weighted (+1 for positive drivers and −1 for negative drivers) mean of activities across all 200 drivers as the PSP:

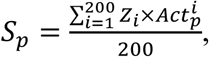

where *Z_i_* is the z-score of hub gene *i* between sensitive and resistant samples, and 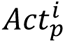 is the activity of the hub gene *i* in sample *p*. The above procedure was also used to estimate PSPs in cell lines.

### Rotenone treatment of B-ALL cells

Drug sensitivity was tested by treating the B-ALL cell line RS;411 with either prednisolone (Solu-Medrol, Pfizer; 0.05, 0.015, or 0.15 µM) alone or in combination with various concentrations of rotenone (R&D Systems; 0.1 pM–0.8 µM). Viability assays were performed using CellTiter-Glo (Promega). Synergistic or additive changes were assessed via response surface modeling^188,189^, which was implemented in MATLAB vR2016a (MathWorks). RS;411 cells were also treated with either 0.019 μM or 0.038 μM rotenone (R&D Systems), harvested at 72 h posttreatment, and then RNA was extracted with TRIzol (Invitrogen). Transcriptomes were interrogated on a Clariom S human array (Affymetrix), which was scanned using a GeneChip Scanner 3000 7G System (Affymetrix).

### Quantitative real-time PCR (ΔCt method)

Endogenous *CHCHD2* expression was measured in the 697 B-ALL cell line (gift from Dr. Charles Mullighan, St. Jude Children’s Research Hospital) treated with either increasing concentrations of ATN-224 (Cayman Chemical) or DMSO vehicle (Sigma-Aldrich). After 24 h, samples from each treatment (n = 3) were collected; RNA was extracted using TRIzol (Invitrogen); and cDNA was synthesized using SuperScript IV VILO Master Mix with ezDNase enzyme (Invitrogen). Standard curves were derived from 10-fold serial dilutions of the DMSO-treated 697 cell line. Quantitative real-time PCR was performed using TaqMan assays HS03043895_g1 (CHCHD2) and Hs99999901_s1 (18s) (Applied Biosystems), TaqMan Fast Advanced Master Mix (Applied Biosystems) and fast cycling conditions (initial incubation: 50°C for 2 minutes; polymerase activation: 95°C for 20 seconds; 40 cycles of denaturation at 95°C for 1 second and anneal/extension at 60°C for 20 seconds) on a QuantStudio7 real-time PCR system (Applied Biosystems). Cycle thresholds and standard curve information were calculated using the QuantStudio7 software, and then each sample’s replicate (n = 3) had its log input normalized to standard curves per gene. Normalized ratios (*CHCHD2*/*18s*) were compared, and mean values were reported (Tukey’s post hoc pairwise testing, n = 9 replicates from 3 experiments).

### Structural analysis and functional modeling of amino acid substitutions

Structural information was derived from human respiratory complex I (PDB IDs: 5XTC, 5XTD) and human respiratory complex III (5XTE)^190^. Structures solved by Esser et al.^79^ were used to model the functional effect of the p.G142E (MT-CYB) substitution, with specific focus on the structure bound to stigmatellin A (PDB ID: 1SQX) as a starting point for docking and molecular dynamics (MD) simulations. The Schrodinger software suite was used to model MT-CYB and to perform docking (*Glide*)^191^, and *LigPrep* and *Prime*^192^ were used for ligand and protein preparation, respectively. Docking was done to confirm ubiquinol binding to cytochrome b and to understand the interactions between the bound ligand and cytochrome b residues. The configurations observed for ubiquinol, along with structural information available from other crystal structures (e.g., PDB ID:1EZV), confirmed that Gly142 occupies a critical area in the ligand-binding pocket of cytochrome b.

MD simulations started with the 1SQX structure, using CHARMM-GUI webserver (*Membrane Builder*)^193,194^ to build a system that utilized either pure phosphatidylcholine (POPC) lipids or binary mixtures of POPC and ubiquinol lipids at different ratios. MD simulations of WT and G142E-mutant MT-CYB were also performed, with and without bound ligand, to test the potential effect of the p.G142E mutation on ligand binding. The final system was solvated in water (TIP3P model)^195^, and the charge was neutralized with 150 mM NaCl ions. CHARMM36m force-field^196^ with a 2 fs time step was used to integrate the forces, and long-range electrostatics were treated using Particle Mesh Ewald^197^ with a real space cutoff of 1.2 nm and 0.12 nm Fourier grid spacing (0.12 nm cutoff was also applied to van der Waals interactions). Temperature was kept at 310K by using a Nosé-Hoover thermostat^198,199^; a Parrinello-Rahman barostat^200^ kept the pressure at 1 bar, using a coupling constant of 5 ps and compressibility of 4.5 × 10^-5^ bar^-1^. Covalent bonds to hydrogen atoms were constrained using the LINCS algorithm^201^. All simulations were performed using GROMACS software (v2021)^202,203^, where simulation lengths varied based on the setup employed (100 ns for WT and p.G142E mutant proteins only; 300 ns for WT and p.G142E mutant with bound ligand; up to 500 ns for unbiased ligand binding).

Cavities were measured using mdpocket^204^ with default settings. The software identified several pockets; therefore, a pocket that coincided with the ligand-binding cavity was manually selected and, for consistency, the same coordinates were applied for both WT and the p.G142 mutant. ProLint^205^ was used to analyze the number of water molecules. Raw data (transparent plots) were plotted with their rolling mean (averaged >50 samples), and protein visualizations were done using VMD^206^ and Mol*^207^. Finally, hydrophobic contacts were calculated by using the default setting in Mol*.

### Statistical analyses

Statistical analyses were performed using the R statistical package (www.r-project.org). Associations between categorical variables were evaluated using a two-sided Fisher’s exact test and Poisson-regression modeling was used to examine the associations between variant frequency and disease subtypes. Correlations between nuclear SNVs and mtDNA SNVs were analyzed with Spearman rank-order and Pearson’s product-moment where appropriate. The mtDNAcn differences were analyzed using log_2_-transformed values. Differences in mtDNAcn among different tumors and subtypes and differences in TTR among relapsed groups were determined using Wilcoxon signed-rank testing. Two-way ANOVA was used to test interactions between relapse groups and risk by using log_10_-transformed TTR. PSP score comparisons excluded patients with intermediate PSP scores, so that 92 (Total Therapy XV), 69 (Total Therapy XVI), and 179 (n=115 pediatric and n = 64 adult) B-ALLs were compared (Supplementary Fig. 8b-c and Supplementary Dataset 8).

### Bootstrap procedure

A bootstrapping procedure^208^ was used to rigorously evaluate the association of variants’ properties, disease characteristics, and patient outcomes. Our procedure generated 10,000 bootstrap datasets by resampling patients with replacement. This process mimics 10,000 replications of a study that selects patients and collects data similar to those of our cohort. We then computed an estimate of an association parameter for the scientific question of interest with the original data and each of the 10,000 bootstrap datasets. A 95% bootstrap confidence interval for the association parameter was defined and computed as the 2.5 percentile and 97.5 percentile of the estimates from the bootstrap samples. All the association parameters estimated by our bootstrap procedure were defined on the real line, and a value of zero corresponded to the null hypothesis. Therefore, a bootstrap *P-*value was defined and computed as the proportion of bootstrap samples giving an estimate with sign opposite to that obtained for the original dataset; this definition is motivated by inverting the bootstrap interval (opposite process as inverting a test to obtain a confidence interval).

We defined and computed several association parameters, as appropriate for the scientific question under evaluation. One association parameter was the difference in prevalence of a categorical property of a variant across two groups of variants. The difference in prevalence was used to evaluate the associations of variant properties, such as the predicted impact of a variant or base pair substitution category (C>T *vs.* T>C), with one another or with characteristics of the patient’s disease (such as diagnosis or disease subtype). Another association parameter was the difference of the mean or median of a quantitative characteristic of a variant across two groups of variants. For cross-tabulations, we computed the differences between the observed cell count data and the expected null cell count.

## Data Availability

PCGP WGS and RNA-seq datasets are available through St. Jude Cloud (https://platform.stjude.cloud/data/cohorts), accession code SJC-DS-1001. Datasets from WGS of Chinese pediatric relapse tumors may be obtained from the corresponding author of Li et al., and WGS of relapsed PCGP genomes may be accessed from the European Genome-Phenome Archive (accession code EGAS00001003975). RNA-seq datasets used for PSP analysis can be found at the Gene Expression Omnibus (GEO) under accession codes GSE115525 and GSE124824. Otherwise, data in present work contained within manuscript.

https://ega-archive.org/datasets/

https://www.ncbi.nlm.nih.gov/gds

## DATA AVAILABILITY

PCGP WGS and RNA-seq datasets are available through St. Jude Cloud (https://platform.stjude.cloud/data/cohorts), accession code SJC-DS-1001. Datasets from WGS of Chinese pediatric relapse tumors may be obtained from the corresponding author of Li et al.^137^, and WGS of relapsed PCGP genomes may be accessed from the European Genome-Phenome Archive (accession code EGAS00001003975). RNA-seq datasets used for PSP analysis can be found at the Gene Expression Omnibus (GEO) under accession codes GSE115525 and GSE124824.

## CODE AVAILABILITY

Source code for single-cell analyses can be found at https://github.com/disonchang/Mito_scRNA. Codes for NetBID (v.2.1.1) analysis are available at GitHub (https://github.com/jyyulab/NetBID). SJARACNe (v.0.2.1, https://github.com/jyyulab/SJARACNe) was utilized within the NetBID environment. The Mitovolve package can be found at https://github.com/yonghui-ni/Mitovolve.

## ACKNOWLEDGMENTS

We thank the patients and families who donated specimens for research purposes and are grateful for the invaluable contributions from our clinicians and research staff. We thank the Genome Sequence Facility (Dana Roeber, Melanie Loyd, and Scott Olsen) of the Hartwell Center for Bioinformatics & Biotechnology, the Flow Cytometry and Cell Sorting core facility (Richard Ashmun and Stacie Woolard), and the Pharmacotyping Resource of the Department of Pharmacy and Pharmaceutical Sciences at St. Jude Children’s Research Hospital (St. Jude) for their exceptional service. We also thank Mikhail Alexeyev (University of South Alabama) for providing parental and mtDNA-deficient 143B cell lines. Special thanks to Ilaria Iacobucci and Yunchao Chang for excellent technical guidance regarding single-cell RNA sequencing, to Brandon Smart for assistance with the rotenone-treatment studies, to Carl Panetta for use of his RSM MATLAB program (all St. Jude); and to Judy Hirst (MRC Mitochondrial Biology Unit, University of Cambridge) and Zhaowen Luo (St. Jude) for expertise and assistance with protein structures. We also thank Natalia Nedelsky for assisting with manuscript preparations. This work was supported in part by funds from NIH grants (R01GM132231 [M.K.], R01GM134382 [J.Y], R01CA36401 [W.E.E], P50GM115279 [W.E.E.], U01 GM92666 [W.E.E.], the St. Jude Comprehensive Cancer Center grant CA21765 from the National Cancer Institute, and by the American Lebanese Syrian Associated Charities. The content is solely the responsibility of the authors and does not necessarily represent the official views of the National Institutes of Health.

## SUPPLEMENTARY FIGURES

**Figure S1.**
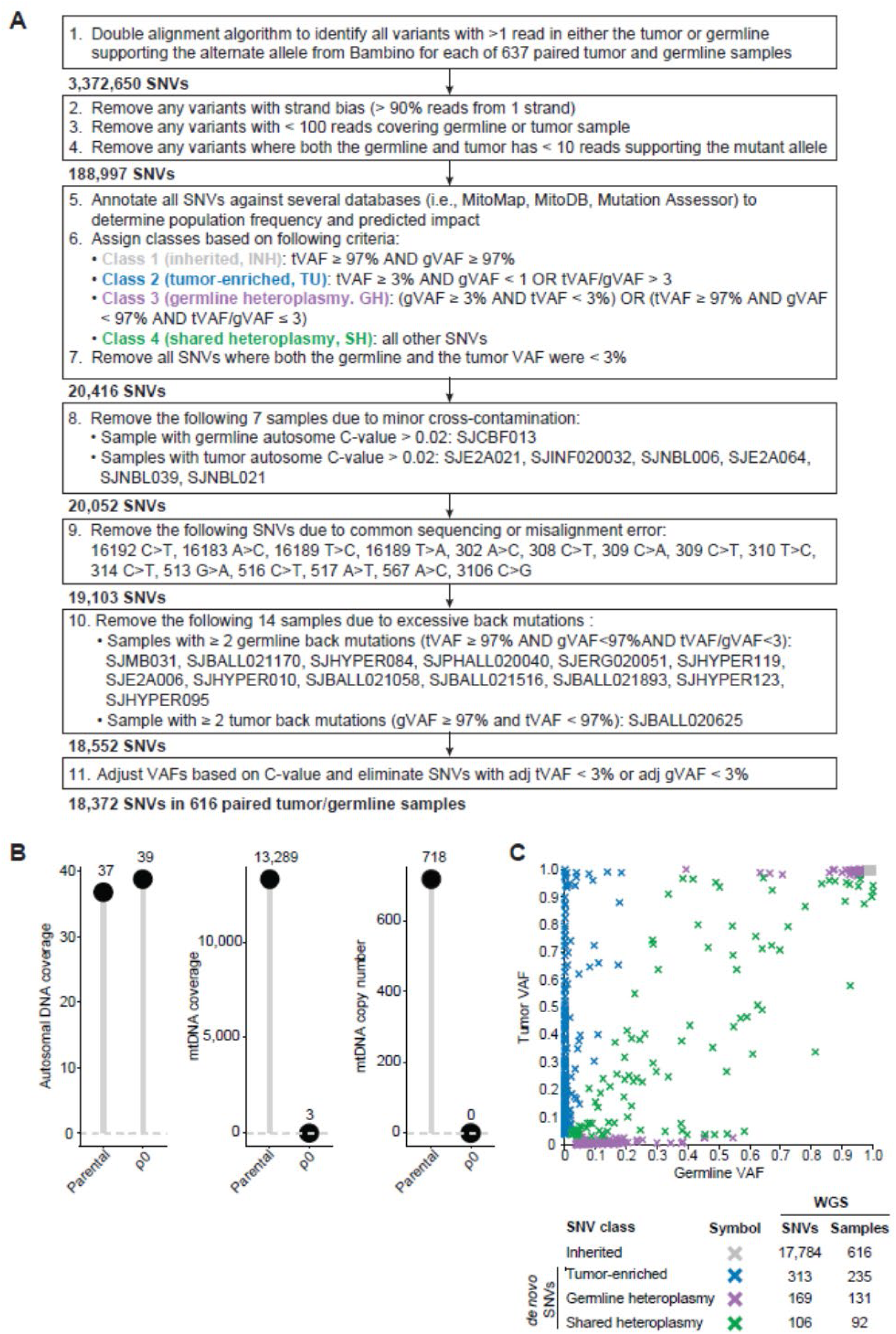
Pipeline for the identification and classification of mtDNA variants. **(A)** Curation of mtDNA variants from paired tumor–germline samples collected under the auspices of the PCGP^48^. **(B)** WGS pipeline validation results for a human 143B TK-osteosarcoma cell line (parental) and its mtDNA-deficient derivative (Rho0, ρ0). Lollipop barplots indicate equivalent autosomal coverage between cell lines (left), yet negligible mitochondrial genome coverage (middle) and absence of mtDNAcn in the ρ0 cell line (right). **(C)** Distribution of 18,372 mtDNA SNVs based on tumor and germline (nontumor) VAFs. SNV classification (depicted by color) is based on the ratio of VAFs estimated by WGS. SNV and sample counts per SNV class are shown in the bottom panel. **Abbreviations**: gVAF, germline variant allele fraction; mtDNAcn, mitochondrial genome copy number; PCGP, Pediatric Cancer Genome Project; SNV, single nucleotide variant; tVAF, tumor variant allele fraction; WGS, whole-genome sequencing.

**Figure S2.**
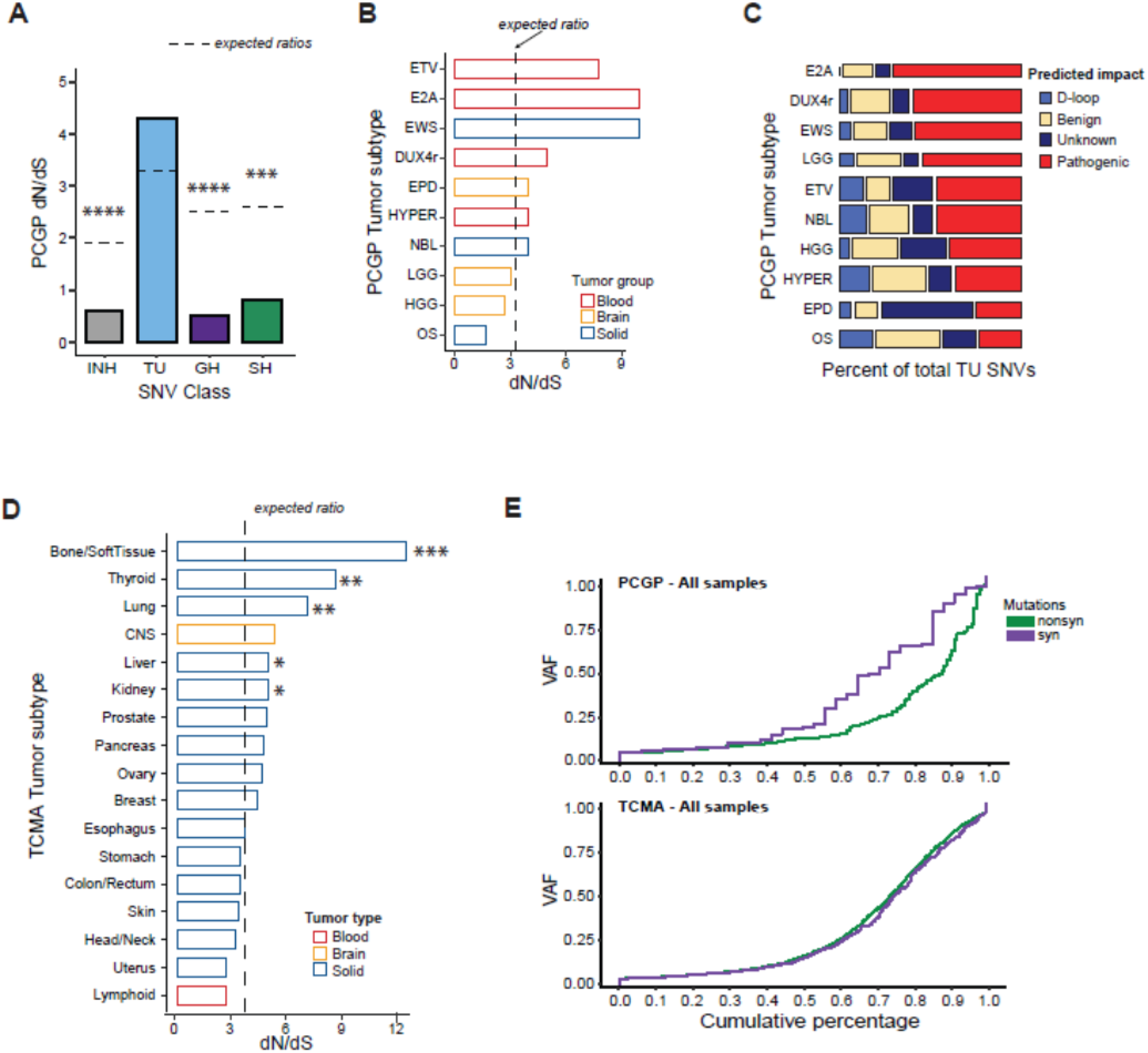
Characterization of TU mutations in PCGP and TCMA cohorts. **(A)** Barplot illustrating observed dN/dS ratios among different classes of pediatric mtDNA SNVs. SNV classifications are depicted by color and abbreviation. \*\*\**P* ≤0.001; \*\*\*\**P* ≤0.0001. Dashed gray lines show the expected dN/dS ratio for each SNV class after considering the mutational signature. **(B)** Observed ratio of non-synonymous to synonymous substitutions (dN/dS) among TU SNVs in pediatric tumor subtypes with at least 30 samples. The expected dN/dS ratio is denoted by a dashed line. Bootstrapped P-values (ETV, P = 0.0954; E2A, P = 0.0864; EWS, P = 0.0638). **(C)** Mosaic plot illustrating the proportion of total TU SNVs distributed among each subtype (width of horizontal, stacked bar), and the proportion of SNVs that are classified by predicted functional impact. Subtypes with fewer than 30 samples were excluded from the analysis. **(D)** Barplot depicting observed dN/dS ratios among adult tumors from the TCMA cohort, grouped by tumor type (blood, brain, or solid). The expected dN/dS ratio (3.7) for adult tumors is denoted by a dashed line. \**P* ≤0.05; \*\**P* ≤0.01; \*\*\**P* ≤0.001. **(E)** A comparison of overall cumulative VAF distributions between nonsynonymous (green) and synonymous (purple) mutations, within the PCGP (top panel) and TCMA (bottom panel) cohorts. **Abbreviations**: dN/dS, ratio of nonsynonymous-to-synonymous substitutions; DUX4r, *DUX4*-rearranged ALL; E2A, *E2A*–*PBX1* ALL; EPD, ependyoma; ETV, *ETV6–RUNX* ALL; EWS, Ewing sarcoma; GH, germline heteroplasmy; HGG, high-grade glioma; HYPER, hyperdiploid; INH, inherited germline heteroplasmy; LGG low-grade glioma; mtDNA, mitochondrial DNA; NBL, neuroblastoma; OS, osteosarcoma; PCGP, Pediatric Cancer Genome Project; SH, shared heteroplasmy; SNV, single-nucleotide variant; TCMA^41,58^, The Cancer Mitochondrial Atlas; TU, tumor-enriched; VAF, variant allele fraction.

**Figure S3.**
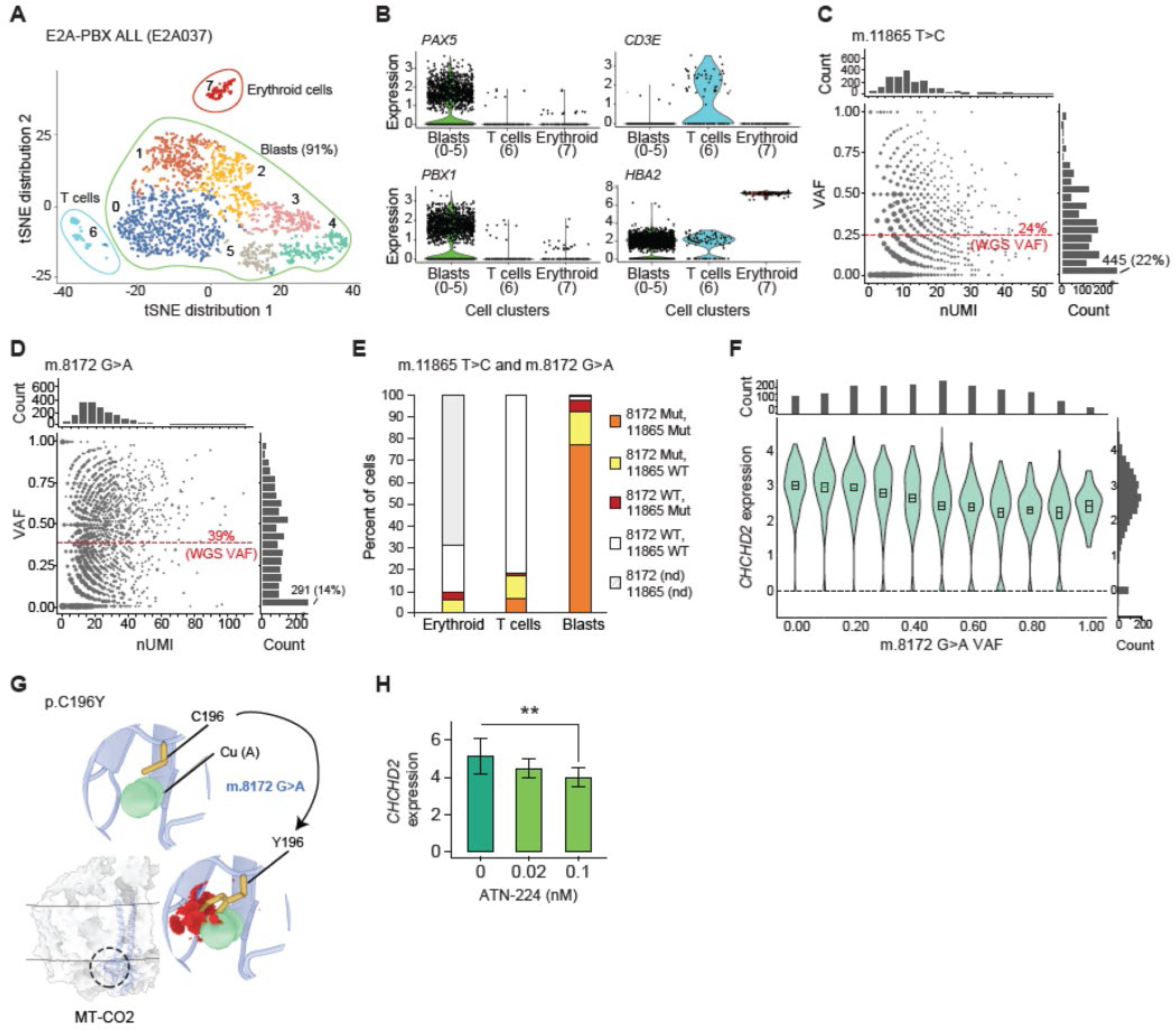
Single-cell RNA sequencing reveals that m.8172G>A is the dominant tumor-specific variant among E2A037 leukemic blasts. **(A)** A tSNE plot illustrating discrete clusters of cell populations and blast subpopulations within the E2A037 sample, with colors and numbers delineating clusters. **(B)** Representative violin plots of select cell-lineage markers with their expression and distribution among tSNE clusters. **(C)** Bubble plot depicting the allelic fraction of the m.11865T>C variant (VAF) as a function of the number of unique molecular identifiers (UMIs) among 2,074 analyzed cells. Dashed red line indicates the bulk tumor WGS estimate. Marginal plots indicate the number of cells binned per nUMI (top) and per VAF (right). VAF counts are binned in 5% increments (e.g., the lowest VAF bin includes cells with 0 to <0.05 VAF). **(D)** Bubble plot depicting the VAF of E2A037’s second TU variant, m.8172G>A, among 2,074 analyzed cells. The plot is as described in panel c. **(E)** Stacked bar plots detailing the percentage of cells harboring each of the E2A037 variants and their dissemination among cell clusters. **(F)** Relation between log-normalized *CHCHD2* expression and VAF (binned) of the m.8172G>A mutation. The bins are 0, 0-0.1, 0.1-0.2…, 0.9-1.0, and boxplots inside each violin show the bootstrapped 95% CI of the mean. Marginal plots indicate the number of cells per binned VAF (top) and per normalized gene expression value (right) intervals. **(G)** Structural modeling of the cysteine-to-tyrosine conversion at position 196 (p.C196Y) that results from the m.8172G>A mutation. The mutation is predicted to alter copper binding within the binuclear copper A center (CU_A_) of MT-CO2 (mitochondrially encoded cytochrome c oxidase II). **(H)** Pharmacological sequestration of copper results in attendant reduction of *CHCHD2* expression, as measured by real-time PCR (ΔCt method). The B-ALL cell line 697 was subjected to increasing concentrations of ATN-224 (bis-choline tetrathiomolybdate), a high-affinity copper chelator that inhibits complex IV function. At 24 h post-treatment, significant (*P* = 0.024, ANOVA) dose-dependent decreases in *CHCHD2* expression were observed, with the largest difference between vehicle only and 0.1 nM ATN-224 (Tukey’s post-hoc pairwise testing, n = 9 replicates from 3 experiments, adjusted \*\**P* = 0.00897). **Abbreviations**: ALL, acute lymphoblastic leukemia; *CD3E*, CD3 epsilon subunit of T-cell receptor complex; *CHCHD2*, coiled-coil-helix-coiled-coil-helix domain containing 2; *HBA2*, hemoglobin subunit alpha 2; Mut, mutant; nd, not determined; nUMI, number of unique molecular identifiers; *PAX5*, paired box 5; *PBX1*, PBX homeobox 1; tSNE, t-distributed stochastic neighbor embedding; VAF, variant allele fraction; WGS, whole-genome sequencing; WT, wild-type.

**Figure S4.**
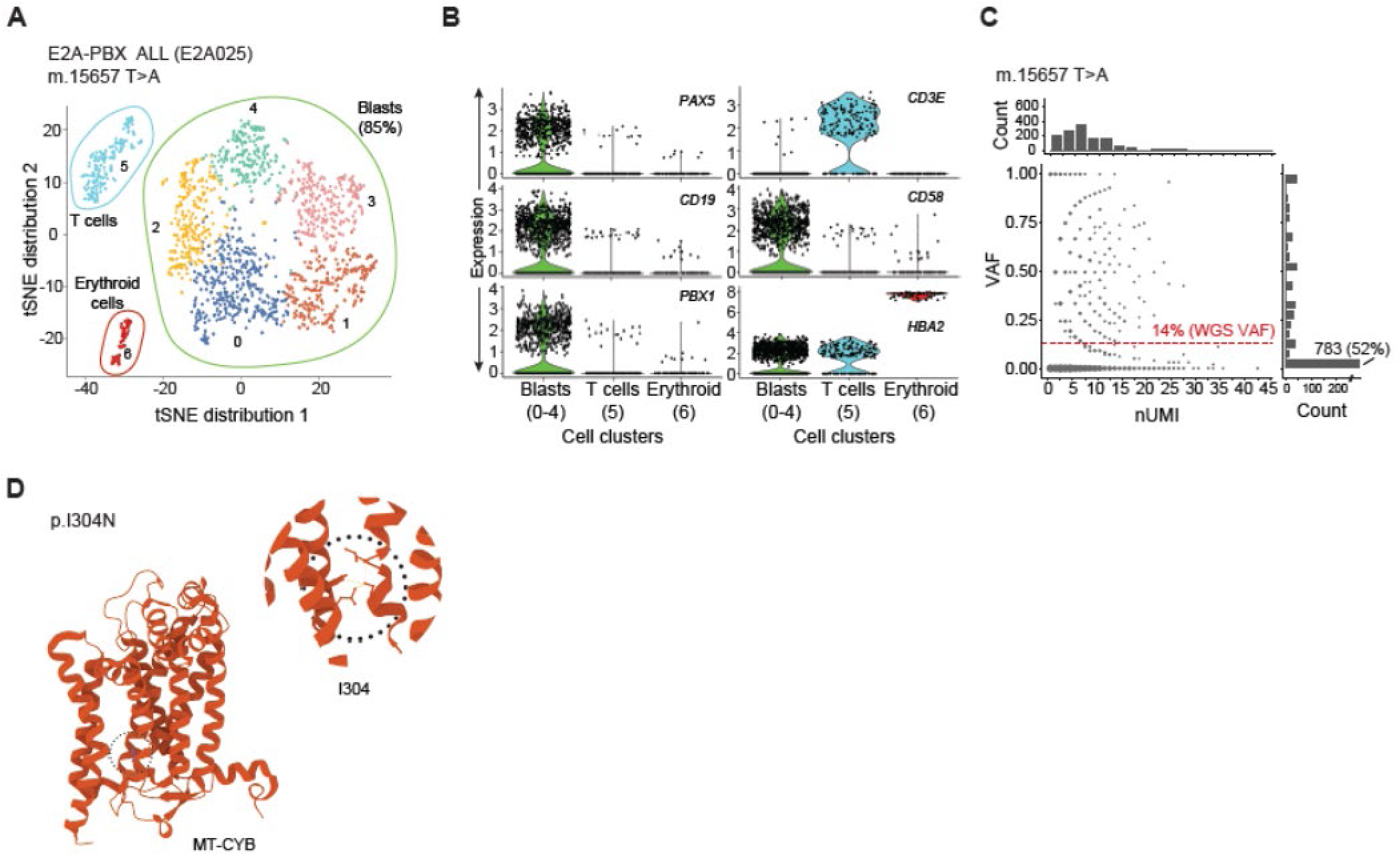
Single-cell RNA sequencing of E2A-PBX1^+^ sample E2A025 reveals VAF distribution pattern for m.15657 T>A variant, which impacts MT-CYB. **(A)** A tSNE plot illustrating discrete clusters of cell populations and blast subpopulations within the E2A025 sample, with colors and numbers delineating clusters. **(B)** Representative violin plots of select cell-lineage markers and their gene expression and distribution among tSNE clusters. **(C)** Bubble plot depicting the VAF of the m.15657T>A as a function of nUMIs among 1,591 analyzed cells. Red line indicates the bulk tumor WGS estimate. Marginal histogram plots indicate the number of cells binned per nUMI (top) and per VAF (right). VAF counts are binned in 5% increments (e.g., lowest VAF bin includes cells with 0 to <0.05 VAF). **(D)** Position of the I304 residue in the secondary structure of MT-CYB, and the hydrophobic interactions (inset, yellow lines) between I304 and the surrounding residues (i.e., L102, L301, Y107, and I362) in human MT-CYB. **Abbreviations**: ALL, acute lymphoblastic leukemia; *CD19*, B-lymphocyte antigen CD19; *CD3E*, CD3 epsilon subunit of T-cell receptor complex; *CD58*, lymphocyte function-associated antigen 3; *HBA2*, hemoglobin subunit alpha 2; I, isoleucine; L, leucine; MT-CYB, mitochondrially encoded cytochrome b; N, asparagine; nUMIs, number of unique molecular identifiers; *PAX5*, paired box 5; *PBX1*, PBX homeobox 1; tSNE, t-distributed stochastic neighbor embedding; VAF, variant allele fraction; WGS, whole-genome sequencing; Y, tyrosine; WT, wild-type.

**Figure S5.**
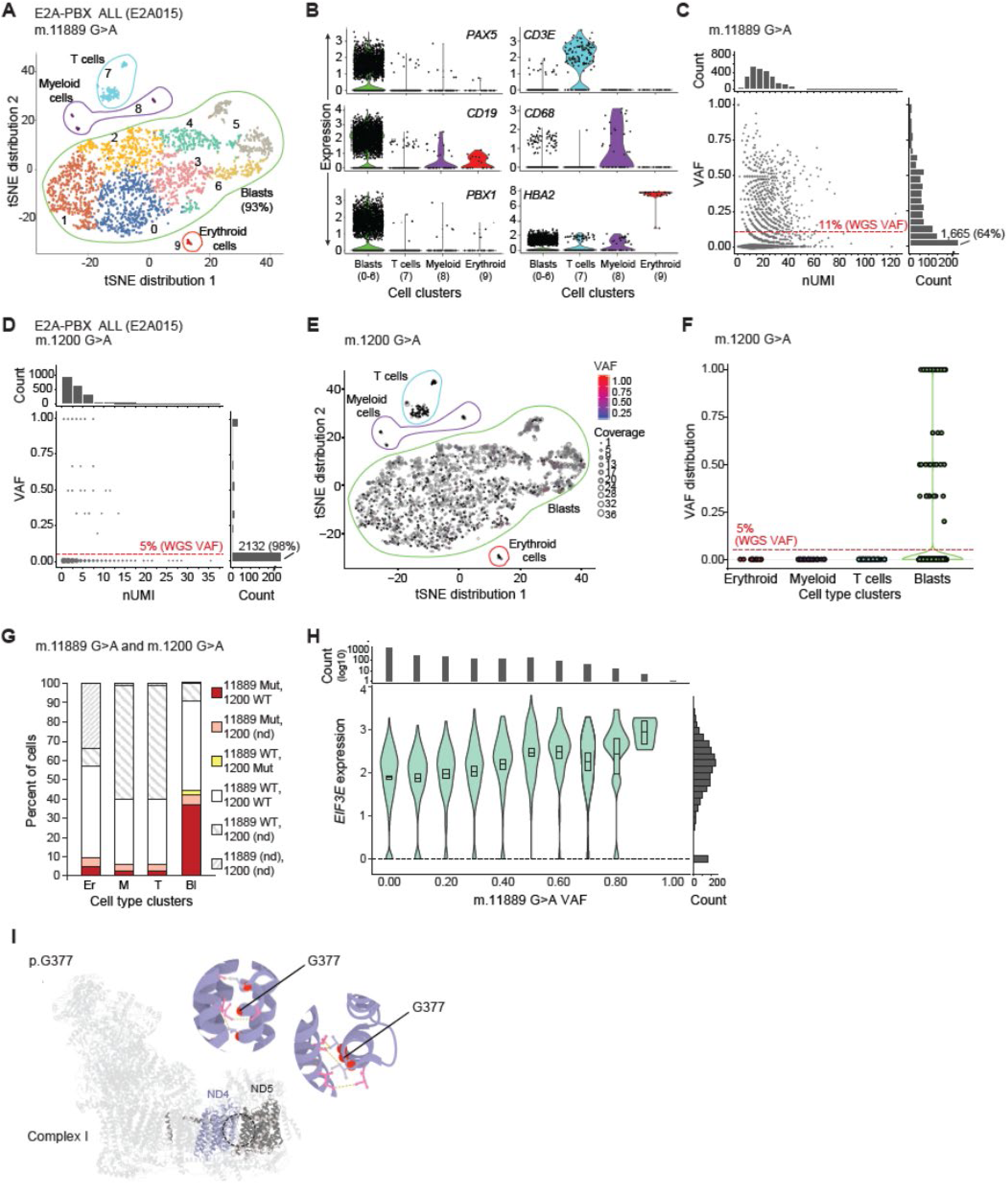
The m.11889 G>A variant in E2A-PBX1^+^ sample E2A015 is the dominant tumor-enriched variant. **(A)** A tSNE plot illustrating discrete clusters of cell populations and blast subpopulations within the E2A015 sample, with colors and numbers delineating clusters. **(B)** Representative violin plots of select cell-lineage markers with their expression and distribution among tSNE clusters. **(C)** Bubble plot depicting the allelic fraction of the m.11889G>A variant (VAF) as a function of the number of unique molecular identifiers among 2,622 analyzed cells. Dashed red line indicates the bulk tumor WGS estimate. Marginal plots indicate the number of cells binned per nUMI (top) and per VAF (right). VAF counts are binned in 5% increments (e.g., the lowest VAF bin includes cells with 0 to <0.05 VAF). **(D)** Bubble plot depicting the VAF of E2A015’s second TU variant, m.1200G>A, among 2,180 cells with coverage at this locus. The plot is as described in panel c. **(E)** Projection of m.1200G>A VAFs onto cells within discrete clusters identified by the tSNE analysis. Scatter point sizes reflect variant coverage. Cells are colored to reflect the magnitude of the VAF. **(F)** Distribution of the m.1200G>A VAF among different cell clusters, demonstrating the paucity of cells harboring the variant; red dashed line indicates the bulk tumor WGS estimate. **(G)** Stacked barplots detailing the percentage of cells harboring each of the E2A015 variants and their dissemination among clusters. **(H)** Increased *EIF3E* expression associated with increased VAF (binned) of the m.11889G>A variant. The bins are 0, 0-0.1, 0.1-0.2…, 0.9-1.0, and the boxplots inside each violin show the bootstrapped 95% CI of the mean for each bin. Marginal plots indicate the numbers of cells per binned VAF (top) and per normalized gene expression value (right). **(I)** Complex I structure (PDB ID: 5XTD) in light grey, with MT-ND4 (light blue) and MT-ND5 (darker grey) proteins highlighted. The p.G377E substitution, which is induced by the m.11889G>A mutation, occurs within a discontinuous helix (TMH12) and is predicted to profoundly alter ND4 function (Mutation Assessor score: 4.81). Expanded view (inset left) and rotated view (inset right) of inward-facing I373, G377, and V381 residues of ND4. Residues that form the protein–protein interface are shown as pink sticks, and hydrophobic interactions are shown as dashed, yellow lines. Red spheres are used to show the location of alpha carbons. **Abbreviations**: ALL, acute lymphoblastic leukemia; *CD19*, B-lymphocyte antigen CD19; *CD3E*, CD3 epsilon subunit of T-cell receptor complex; *CD68* CD68 antigen; *HBA2*, hemoglobin subunit alpha 2; E, glutamic acid; *EIF3E*, eukaryotic translation initiation factor 3 subunit E; G, glycine; MT, mitochondrially-encoded; Mut, mutant; ND4, NADH: ubiquinone oxidoreductase core subunit 4; ND5, NADH: ubiquinone oxidoreductase core subunit 5; nd, not determined; nUMI, number of unique molecular identifiers; *PAX5*, paired box 5; *PBX1*, PBX homeobox 1; PDB, Protein Data Bank; TMH, transmembrane helix; tSNE, t-distributed stochastic neighbor embedding; TU, tumor-enriched; VAF, variant allele fraction; WGS, whole-genome sequencing; WT, wild-type.

**Figure S6.**
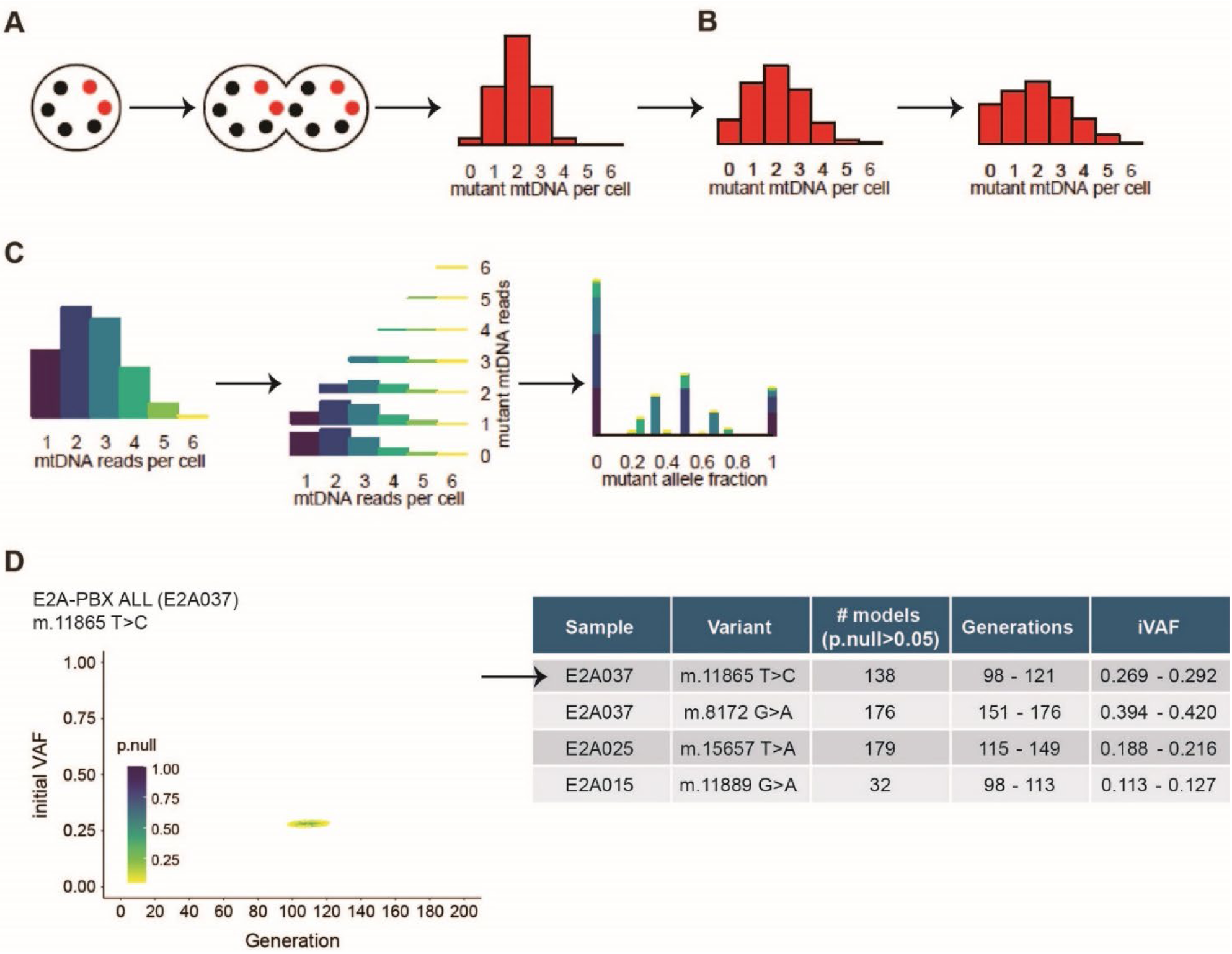
Overview of selection-free modeling process using Mitovolve. **(A)** An initial cell with four wildtype mtDNAs (black) and two mutant mtDNAs (red) followed by duplication of its mtDNA. The red histogram represents the probability distribution for the number of mutant mtDNAs inherited by the daughter cell and is defined by hypergeometric sampling from the duplicated mtDNA. **(B, left panel)** The same process is repeated for the next daughter cell, effectively rendering a convolution of hypergeometric models. Each histogram bar represents the number of inherited mutant mtDNAs by the next daughter cell. **(B, right panel**) The number of mutant mtDNAs in subsequent generations is modeled by iterative convolution of hypergeometric models. **(C, left panel)** Single-cell sequencing generates a distribution of mtDNA reads per cell. **(C, middle panel)** For a given number of total mtDNA reads, the number of mutant mtDNA reads is modeled by hypergeometric sampling of reads from mtDNAs. **(C, right panel)** The model produces a theoretical distribution for the fraction of mutated reads across cells that can be compared to the observed empirical distribution. **(D)** Scatter plot illustrating the range at which selection-free models best approximated the observed scRNA-seq data (p.null ≥ 0.05) for the m.11865T>C variant. Selection-free model testing parameters: iVAFs in 0.01-1 in 0.01 increments; fixed mtDNAcn = 312; generations: 1-200. mtDNAcn was derived empirically from the patient’s WGS data. **Abbreviations**: mtDNAcn, mitochondrial DNA copy number; p.null, p-value that compares each selection-free model with the observed data; iVAF, initial variant allele fraction; WGS, whole genome sequencing.

**Figure S7.**
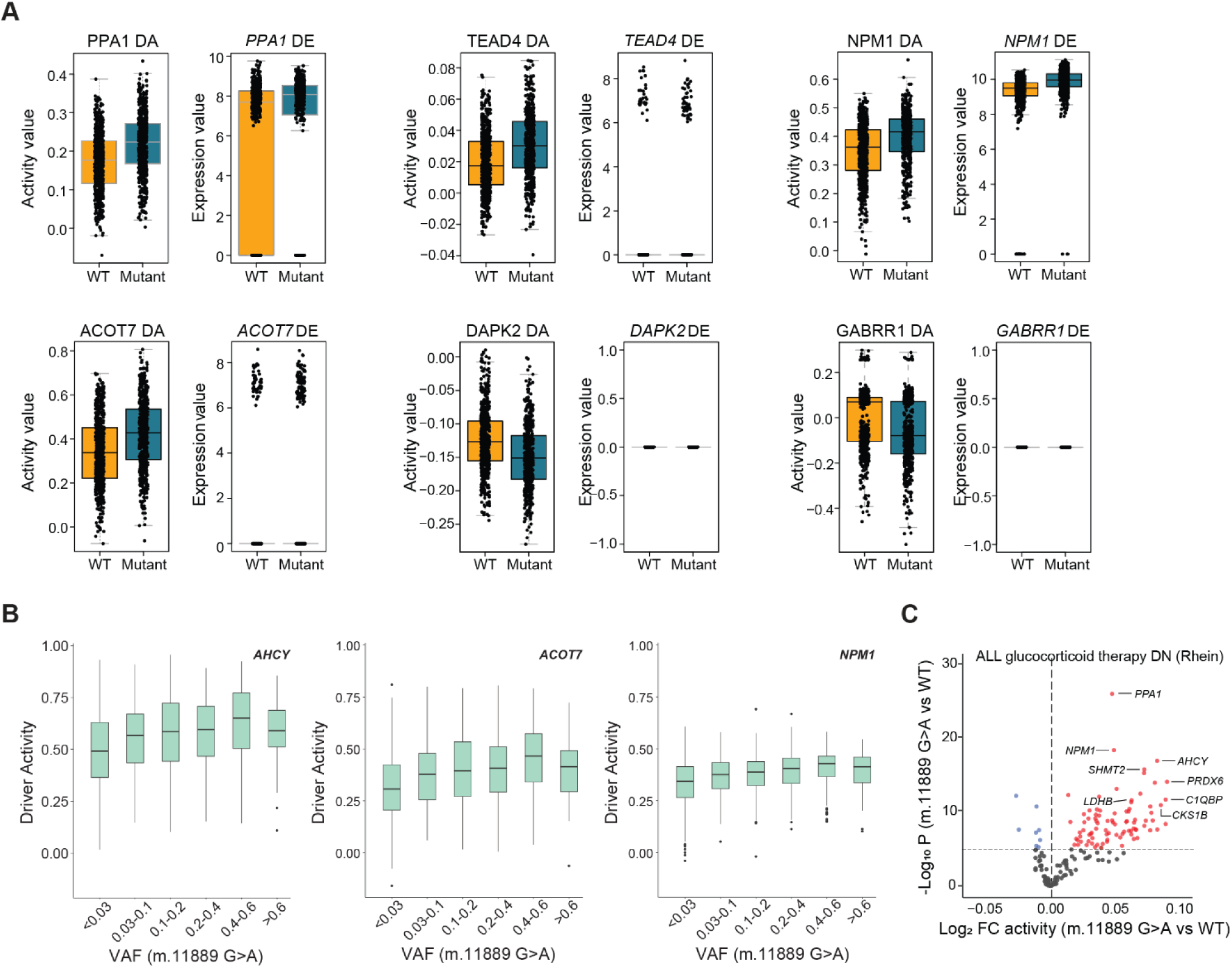
NetBID infers the differential activity of drivers in cells with or without mtDNA variants. **(A)** Paired boxplots comparing driver activity (left panel) and gene expression (right panel) for select genes from the volcano plot in Figure 4B. Comparisons are made between WT cells (gold) and cells harboring the m.11889G>A mutation (Mutant, turquoise). **(B)** Examples of VAF-associated changes in activity levels of drivers that were altered in cells harboring the m.11889G>A mutation. VAFs are binned as < 0.03, 0.3-0.1, 0.1-0.2, 0.2-0.4, 0.4-0.6, and > 0.06. **(C)** Volcano plot illustrating differential activity of drivers from the ALL-Glucocorticoid therapy DN (Rhein) gene set in cells harboring the m.11889G>A mutation as compared to WT cells. **Abbreviations**: *ACOT7*, acyl-CoA thioesterase 7; *AHCY*, adenosylhomocysteinase; ALL, acute lymphoblastic leukemia; DA, differential activity; *DAPK2*, death associated protein kinase 2; DE, differential gene expression; DN, downregulated; FC, fold change; *GABRR1*, gamma-aminobutyric acid type A receptor subunit Rho1; *NPM1*, nucleophosmin 1; *PPA1*, inorganic Pyrophosphatase 1; *TEAD4*, TEA Domain Transcription Factor 4; VAF, variant allele fraction; WT, wild-type.

