## Supplementary Notes for "Somatic mitochondrial DNA mutations are a source of heterogeneity among primary leukemic cells"

- Supplementary Notes (with figures) 1-10
- Supplementary Note Tables 1-2
- Supplementary References

**Supplementary Note 1. Mitochondrial DNA copy number (mtDNAcn) analyses by tumor subtype.**

Tissue-type analysis revealed a significantly greater mtDNAcn in tumors than in the germline samples (*P* <0.0001 for blood, brain, and solid tumors; Wilcoxon signed-rank test,). With a few exceptions, this trend was maintained across cancer subtypes (**Table S1A**). Subtype-specific differences in mtDNAcn were also observed within each tissue type (**Figure SN1**). Adrenocortical carcinomas had the greatest mtDNAcn (median: 832 copies/cell) of any subtype ($P = 2.95 \times 10^{-11}$, Wilcoxon rank sum test), a finding that was not unexpected, given the intrinsic relation between mitochondria and steroidogenesis and that excessive hormone production and oncocytic features are hallmarks of these tumors[1,2]. Higher copy numbers were also observed more frequently among brain tumors (median range: 500-674 copies/cell), whereas leukemias tended to predominate at the lower end of the mtDNAcn spectrum (median: 275 copies/cell). Infant B-ALL cases possessed the fewest copies (median: 182 copies/cell) among all subtypes and had significantly lower mtDNAcn than all other blood cancers ($P = 7.55 \times 10^{-5}$, Wilcoxon rank sum test).

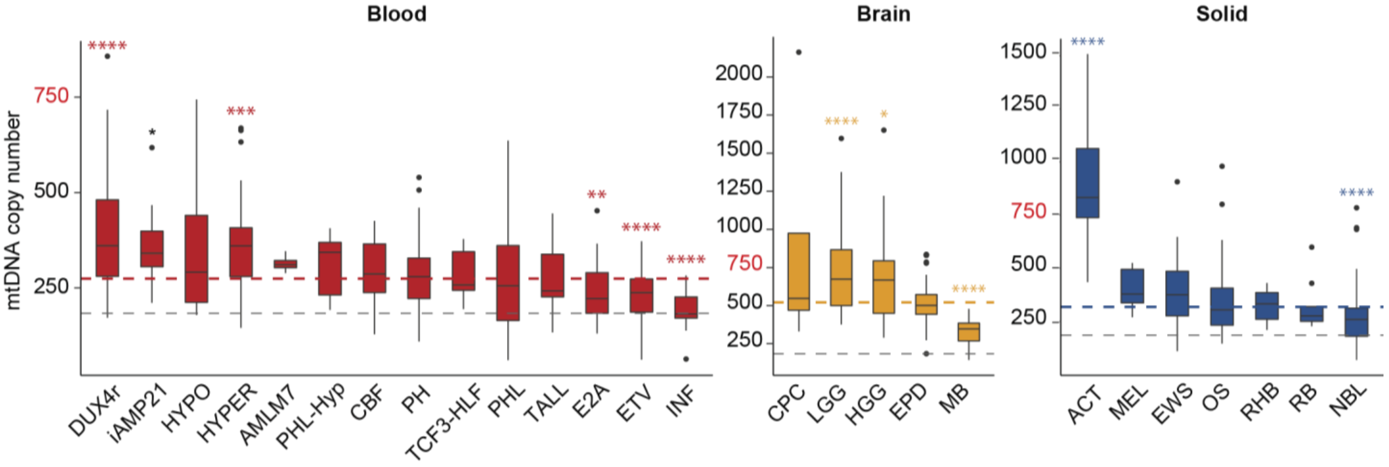


**Figure SN1. Mitochondrial DNA copy number across tumor subtypes.** Subtype-specific distribution of mtDNAcn among blood (red), brain (orange) and solid (blue) tumors. Dashed grey lines represent the median germline mtDNAcn within each tumor group; dashed colored lines represent the median tumor mtDNAcn within each tumor group; red font on the y-axis (750) is a benchmark mtDNAcn to highlight changes in scale across tumor groups. Significant differences are indicated between each subtype and all other subtypes within the same tumor group. Comparisons were made using two-sided Wilcoxon signed-rank tests. **P* ≤0.05; ***P* ≤0.01; ****P* ≤0.001; *****P* ≤0.0001. **Abbreviations:** ACT, adrenocortical carcinoma; AMLM7, acute megakaryoblastic leukemia; CBF, core-binding factor; CPC, choroid plexus carcinoma; DUX4r, *DUX4*-rearranged; E2A, *E2A–PBX1* translocation; EPD, ependymoma; ETV, *ETV6–RUNX1* translocation; EWS, Ewing sarcoma; HGG, high-grade glioma; HYPER, hyperdiploid; HYPO, hypodiploid; iAMP21, intrachromosomal amplification of chromosome 21; INF, infant ALL with/without *MLL* rearrangements; LGG, low-grade glioma; MB, medulloblastoma; MEL, melanoma; NBL, neuroblastoma; OS, osteosarcoma; PH, Philadelphia chromosome/*BCR–ABL1* translocation; PHL, Philadelphia chromosome–like; PHL-Hyp, PHL with either hyperdiploid or hypodiploid features; RB, retinoblastoma; RHB, rhabdomyosarcoma; TALL, T-cell acute lymphoblastic leukemia; TCF3-HLF, *TCF3–HLF* translocation.

**Supplementary Note 2. Characterization of TU mtDNA SNVs.**

Compared to adult malignancies, significantly fewer pediatric malignancies harbored at least one TU mtDNA SNV (*P* <0.0001, Fisher's exact test; **Figure SN2A**). In addition, samples with multiple TU SNVs, which comprised 31.1% of the TCGA cohort, were observed less frequently (10.7%) among pediatric samples. These differences may reflect the distinct tumor-type compositions of the adult and pediatric cohorts, tissue-specific somatic mutation rates[3], and/or an increased propensity to acquire somatic mutations with age[4,5]. Among pediatric samples, there was a significant association (*P* = 0.04, Poisson regression) between tumor group and the proportion of samples with TU SNVs, which was highest among the hematological malignancies (**Figure SN2B**). Rhabdomyosarcomas harbored the fewest TU SNVs, and T-cell ALL harbored the most (**Figure SN2C**).

We also explored the data for any relation between the numbers of TU mtDNA SNVs and annotated nuclear DNA SNVs among individual samples with nuclear SNV data available (**Figure SN2D** and **Table SN1**). We identified a positive correlation between mtDNA SNVs and Tier 1 nuclear DNA SNVs (i.e., coding synonymous, nonsynonymous, splice-site, and noncoding RNA variants) across all three cancer groups (Rs = 0.17, *P* = 0.005 for blood; Rs = 0.23, *P* = 0.007 for brain; and Rs = 0.23, *P* = 0.003 for solid; Spearman's rank; **Figure SN2D**). This correlation was strongest among samples of Ewing sarcoma (Rs = 0.53, *P* ≤0.001) and DUX4r B-ALL (Rs = 0.43, *P* = 0.02), in which positive correlations were also observed between the number of TU mtDNA and Tier 2 (regulatory regions) and/or Tier 3 (intergenic and intronic) nuclear DNA SNVs (**Table SN1**). Because >60% of the TU SNVs in these tumor subtypes were predicted to be pathogenic and mostly affect complex I, III, or IV function (**Table S1B**), such mtDNA mutations might promote nuclear genome alterations (e.g., by increasing reactive oxygen species [ROS] production)[6-10] and/or render cells more permissive to their accumulation.

We next sought to examine the incidence of truncating and recurrent mtDNA SNVs. Truncating mutations were rare (n=11) (**Figure SN3A**) and primarily found in solid tumor samples (n=8) (**Table SN2**). These premature termination events were often the result of G>A substitutions (n=9) in either glycine or tryptophan, and many were localized to complex I (n=6). The appearance of G>A (C>T) mutations was not limited to stop–gain mutations and constituted 48% (n=70) of all deleterious mutations.

Excluding synonymous variants, we found 11 recurrent mtDNA SNVs (3 SNVs in D-loop, and 8 missense mutations in coding sequences), with each occurring in 2-3 patients (**Figure SN3A** and **Table SN2**). Three samples harbored substitutions within MT-CYB, at position 304 (I304F, I304N, I304T), which conferred various degrees of functional impact based on mutation assessor scoring[11], and all of which were rare (<0.001%) in the general population. Three pediatric tumor samples harbored the m.72T>C TU variant, which has been not only frequently observed as a TU mtDNA SNV in adult malignancies[12] but also reported as a somatic substitution in various noncancerous adult tissues[13]. In comparison, this variant has an allelic frequency of 1.8% in MitoMap[14] and was detected as an INH SNV in 2.4% (n = 15/616) of patients in our pediatric cancer cohort (**Table S1B**).

In addition to the aforementioned m.72T>C variant, several recurrent TU SNVs were identified in both pediatric and adult tumor samples. The clinically relevant m.3243A>G mutation (in *MT-TL1*) and variants profiled in this study (i.e., m.8172G>A, m.11889G>A, and m.15171G>A) are among the most notable of recurrent variants predicted to be pathogenic (**Figure SN3B** and **Table S1C**). We also noted the recurrence of the m.11711G>A variant that is uncommon in the general population but has been previously reported[15,16]. The variant occurs within a highly conserved residue of *MT-ND4* and is predicted to be pathogenic.

We next sought to determine whether mtDNA variants were concentrated in specific genes, complexes, or regulatory loci. We performed a GRIN analysis[17], which assumed a random distribution of variants, with equal probability of distribution across all loci (**Figure SN4A** and Methods). This analysis revealed significantly (*P* ≤0.001) more than expected mtDNA variants in the regulatory D-loop region among GH and SH SNVs but no deviations among the TU variants. When the analysis was restricted to variants predicted to be pathogenic, we observed higher-than-expected mutation frequencies among the TU variants in *MT-ND1*, *MT-COX1*, *MT-CYB*, and 9 of 22 MT-tRNAs; however, only *MT-TM* reached statistically significant levels (*P* <0.01, GRIN) (**Figure SN4B**).

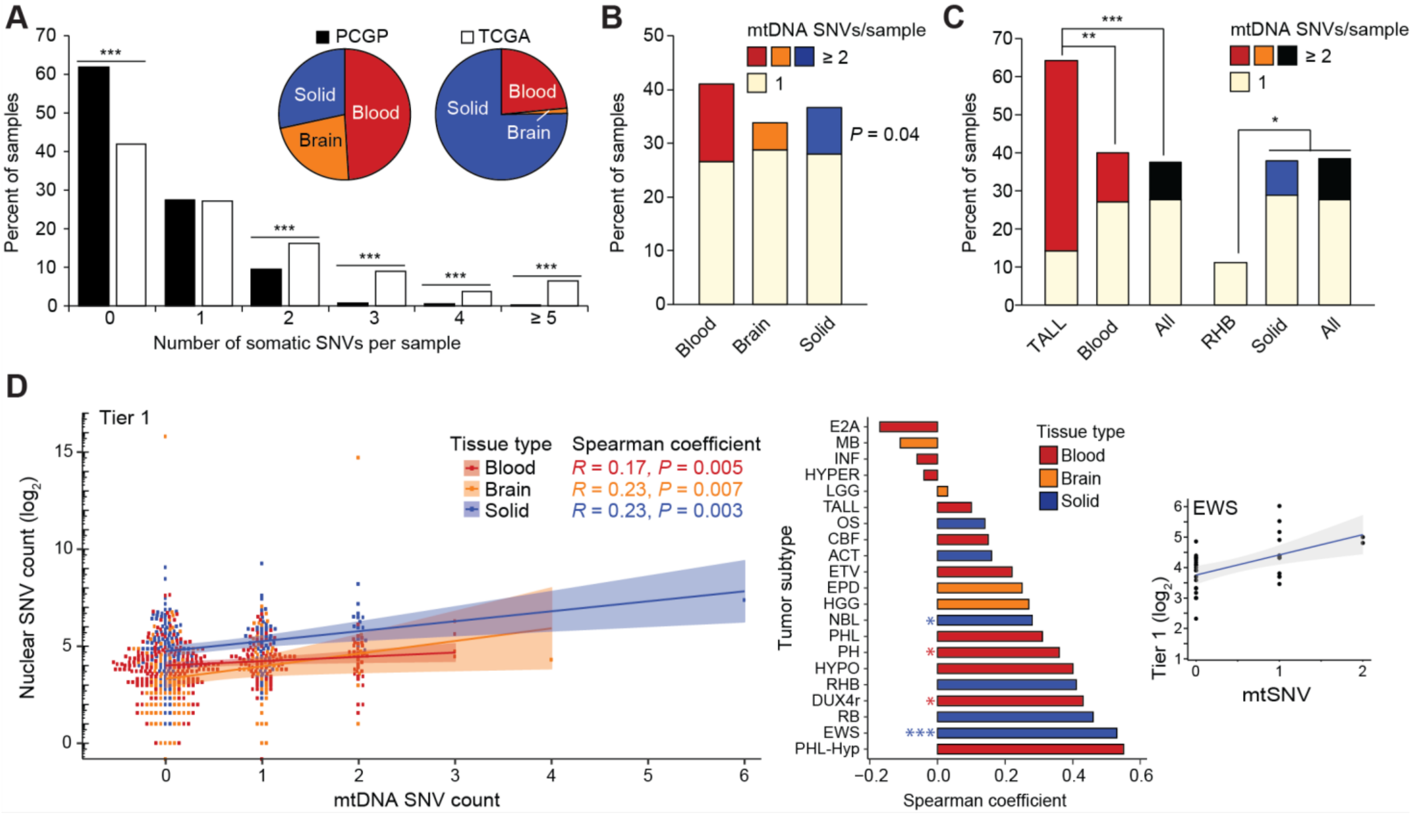

**Figure SN2. TU mtDNA SNVs and correlations with annotated Tier 1 nuclear genes.** (A) Bar plot showing significant differences (****P* ≤0.001, Fisher's exact test) in the percentage of samples harboring discrete numbers of SNVs between pediatric (PCGP[36]) and adult (TCGA) cancer cohorts. Differences in tumor group composition (blood, brain, solid) between the two cohorts are also presented (inset, pie charts). (B) Percentages of samples categorized by tumor group (blood, brain, solid) with either 1 or ≥2 mtDNA substitutions per sample (*P* = 0.04, Poisson regression). (C) Subtype-specific analysis considering the percentage of samples with either 1 or ≥2 mtDNA SNVs per sample. The significant subtypes are highlighted *vs*. their respective tumor group and *vs.* all (All) other tumors (**P* ≤0.05; ***P* ≤0.01; ****P* ≤0.001, Poisson regression). (D) Left panel: correlational analysis between the frequency of Tier 1 nuclear-coding SNVs (i.e., coding synonymous, nonsynonymous, splice-site, and noncoding RNA variants) and mtDNA SNVs stratified by tumor group. Middle panel: subtype-specific correlational analyses between nuclear-coding and mtDNA SNV counts, with Spearman rank coefficients of each subtype plotted in ascending order. **P* ≤0.05; ****P* ≤0.001. Right panel: example of a correlation between substitutional frequency among nuclear-coding genes and mitochondrial SNVs in Ewing sarcoma (EWS) cases. **Abbreviations**: ACT, adrenocortical carcinoma; CBF, core-binding factor; DUX4r, *DUX4*-rearranged; E2A, *E2A–PBX1* translocation; ETV, *ETV6–RUNX1* translocation; EPD, ependymoma; HGG, high-grade glioma; HYPER, hyperdiploid; HYPO, hypodiploid; INF, infant ALL with/without *MLL* rearrangements; LGG, low-grade glioma; MB, medulloblastoma; mtDNA, mitochondrial DNA; NBL, neuroblastoma; OS, osteosarcoma; PCGP, Pediatric Cancer Genome Project; PH, Philadelphia chromosome/*BCR–ABL1* translocation; PHL, Philadelphia chromosome–like; RB, retinoblastoma; RHB, rhabdomyosarcoma; PHL-Hyp, Philadelphia chromosome–like with either hyperdiploid or hypodiploid features; SNV, single-nucleotide variant; TALL, T-cell acute lymphoblastic leukemia; TCGA, The Cancer Genome Atlas.

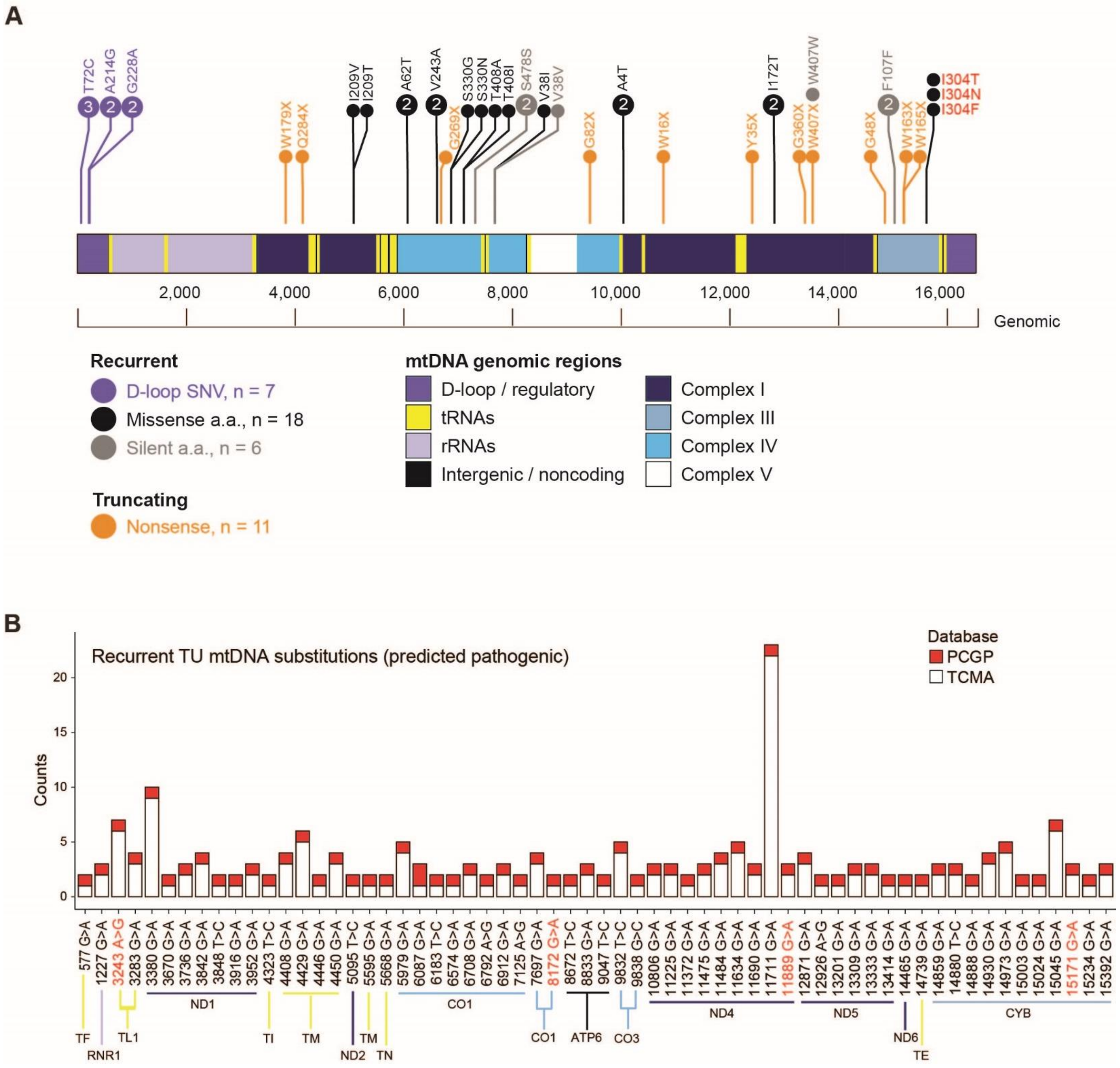


**Figure SN3. Recurrent and truncating TU mtDNA variants.** (A) Protein paint[37] depiction of the mitochondrial genome (1-16,569 base pairs), with annotation of recurrent (see color key) and truncating (orange) mutations. Recurrence is defined as occurring in two or more pediatric samples. D-loop/regulatory mutations are annotated as SNVs; all other mutations are annotated using amino acid–alteration nomenclature. Recurrent mutations affecting the I304 residue in complex III are shown in red. (B) Distribution and frequency of recurrent adult (white bar) and pediatric (red bar) TU mtDNA variants with predicted pathogenic impact across the mitochondrial genome. Recurrence is defined as occurring in ≥1 pediatric samples and in ≥1 adult samples. TU mtDNA SNVs from adult samples were obtained from the TCMA dataset[23,38] by using a tumor VAF cutoff of 0.03. A list of all recurrent mtDNA SNVs (regardless of predicted impact) is provided in Table S1C. The mtDNA SNVs identified in the single-cell sequenced samples are highlighted in red font. **Abbreviations**: a.a., amino acid; ATP6, ATP synthase F0 subunit 6; COX, cytochrome c oxidase; CYB, cytochrome b; mtDNA, mitochondrial DNA; ND, NADH dehydrogenase; PCGP, Pediatric Cancer Genome Project; RNR1, 12S ribosomal RNA; rRNA, ribosomal RNA; SNV, single-nucleotide variant; TCMA, The Cancer Mitochondrial Atlas; TE, tRNA glutamic acid; TF, tRNA phenylalanine; TI, tRNA isoleucine; TL1, tRNA leucine 1; TM, tRNA methionine; TN, tRNA asparagine; tRNA, transfer RNA; TU, tumor-enriched.

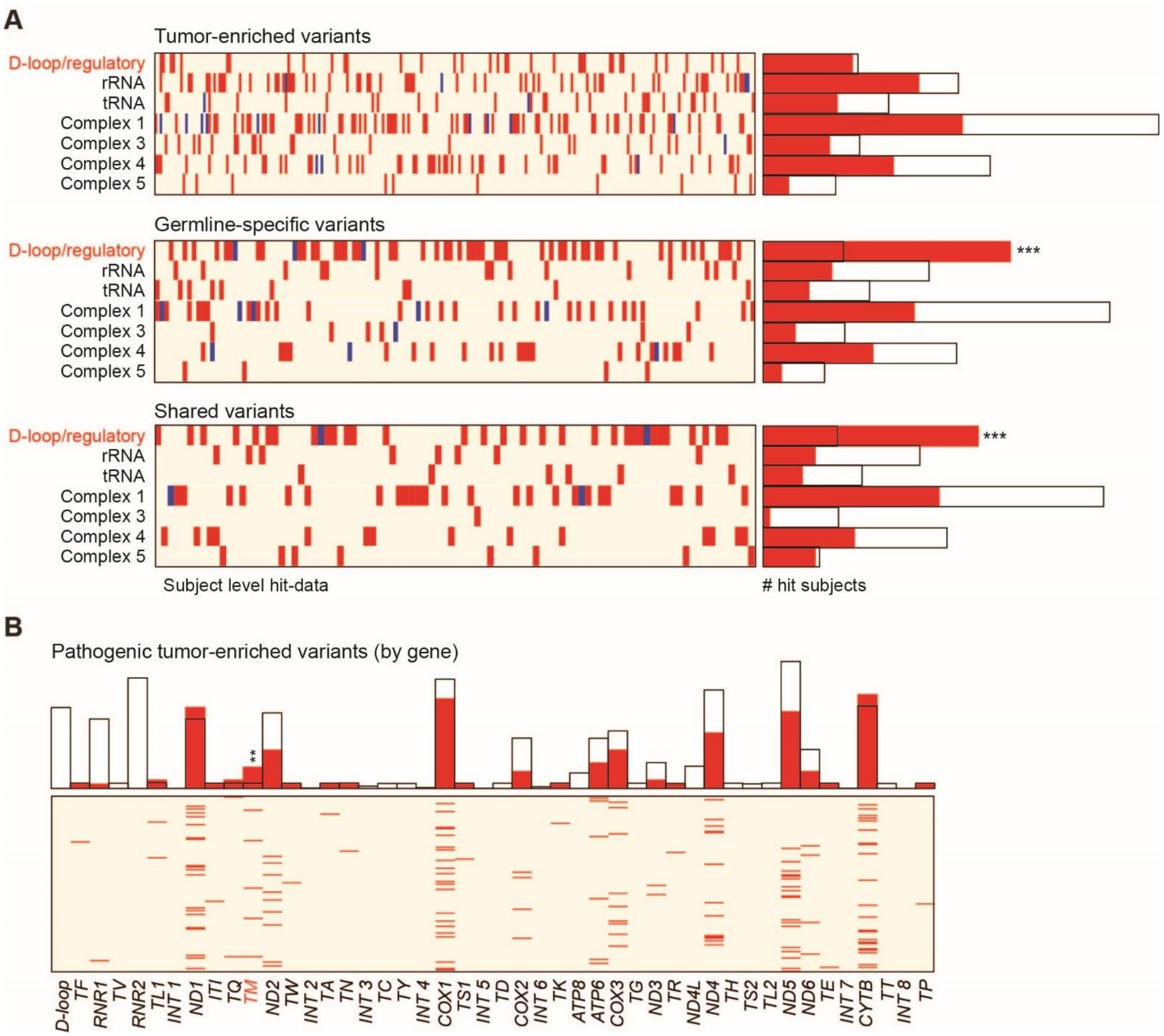


**Figure SN4. Genomic random interval analysis by SNV class and among pathogenic tumor-enriched variants.** (A) Results of GRIN analyses, stratified by somatic SNV class. Within the heatmap (left panel), rows represent (sets of) genes belonging to either respiratory chain complexes or noncoding regions (tRNA, rRNA, D-loop/regulatory). Corresponding subject-level hits are plotted by column: red bars indicate one variant, and blue bars indicate >1 variant. Bar plots of GRIN results (right panel) compare the numbers of subjects who were expected to have one variant per (set of) gene(s) under the null model (unfilled bars) and the observed number of subjects who had a variant in the indicated gene or set of genes (red bars). An overabundance of variants is indicated by the red bar extending farther to the right than the unfilled bar; underabundance of variants is indicated by the unfilled bar extending farther to the right than the red bar. *P*-values were determined by the probability distribution for the number of individuals with a mutation in each given region by chance (****P* ≤0.001). (B) GRIN analysis of pathogenic TU mutations within mtDNA genes. Within the heatmap (bottom panel), columns represent genes and rows represent subject-level hits (red bar = 1 variant). Bar plots of GRIN results (top panel) are as described in panel A (***P* ≤0.01). **Abbreviations:** *ATP6*, ATP synthase F0 subunit 6; *ATP8*, ATP synthase F0 subunit 8; *COX*, cytochrome c oxidase; *CYTB*, cytochrome b; *INT*, intergenic; *ND*, NADH dehydrogenase; *RNR1*, 12S ribosomal RNA; *RNR2*, 16S ribosomal RNA; SNV, single-nucleotide variant; *TA*, tRNA alanine; *TC*, tRNA cysteine; *TD*, aspartic acid; *TE*, tRNA glutamic acid; *TF*, tRNA phenylalanine; *TG*, tRNA glycine; *TH*, tRNA histidine; *TI*, tRNA isoleucine; *TK*, tRNA lysine; *TL1*, tRNA leucine 1; *TL2*, tRNA leucine 2; *TM*, tRNA methionine; *TN*, tRNA asparagine; *TP*, tRNA proline; *TQ*, tRNA glutamine; *TR*, tRNA arginine; *TS1*, tRNA serine 1; *TS2*, tRNA serine 2; *TT*, tRNA threonine; *TV*, tRNA valine; *TW*, tRNA tryptophan; *TY*, tRNA tyrosine.

### Supplementary Note 3. Distribution of mtDNA variants.

We generated a subject-level display of mitochondrial substitutions by location, further enumerated by SNV class and tumor type (i.e., blood, brain, or solid) (**Figure SN5**). As expected, this analysis indicated that INH variants were comparable in distribution and relative frequency to mtDNA variants identified in the general population as estimated by MitoMap[14]. In comparison, *de novo* GH and SH mtDNA variants were distributed throughout the genome, with relatively few recurrences among samples.

### Supplementary Note 4. Mutational signatures among *de novo* variants.

Similar to what has been reported in the adult TCGA cohort[18], transitional substitutions (T>C and C>T) accounted for the majority (92.3%) of all TU mutations, with 81.9% of C>T transitions occurring on the (H-strand, and 64.2% of T>C transitions occurring on the L-strand (**Figure SN6A,B**). In support of an endogenous, strand-asymmetric replication error being the main source of mtDNA mutations[18], we observed a reversal of the L- to H-strand ratio of C>T substitutions within the D-loop's origin of replication, relative to the rest of the mitochondrial genome (**Figure SN6A**). We also observed a similar reversal of L- to H-strand ratio of C>T substitutions within the origin of replication among the GH and SH variants (**Figure SN6C,D**). We did not find any evidence of a corresponding change to the L- to H-strand ratio of T>C substitutions within the same regions of the TU, GH, or SH variants, perhaps because the pediatric cohort had fewer samples than the adult cohort.

A closer examination of trinucleotide context revealed a pattern among pediatric TU SNVs (but not GH or SH SNVs) that approximated the benchmark mtDNA mutational signature observed in adult tumors (cosine similarity L-strand = 0.887, H-strand = 0.891 for TU SNVs)[18]. Computational modeling has suggested that the mtDNA mutational signature observed in tumor cells is similar to one that has been active in the germline during evolution, albeit with relaxation of the negative-selective pressure against functionally deleterious variants that has shaped the ancestral variants[18,19]. Thus, if the source of mutations is the same for all *de novo* variants, then any difference in mutational patterns between the GH or SH variants and the TU variants would suggest that the accumulation of GH or SH and TU variants in cells is subject to different constraints.

### Supplementary Note 5. Sources of variation between RNA and DNA VAFs.

Of the 50 variants with reduced RNA VAFs, 46 belonged to the INH group (**Figure 1B**), encoded synonymous or neutral amino acid changes in subunits of complex I or V, and were most likely a consequence of posttranscriptional RNA editing[20,21] that reverted variants to the reference sequence. Six of the 11 variants with elevated RNA VAFs were TU variants localized to MT-tRNAs, representing a significant enrichment (bootstrap $P = 0.0122$) of such allelic imbalances among MT-tRNA variants in the TU group (6 of 16) compared to those in all other mtDNA SNV groups (1 of 198). Allelic imbalances in somatic tRNA variants also have been reported in adult tumor samples and were noted to cause structural alterations that delay tRNA processing, thereby increasing the likelihood of pathogenicity[22,23]. The presence of the clinically relevant m.3243A>G mutation (*MT-TL1*) among our TU variants with elevated RNA VAFs (19% VAF difference, **Table S1B**, and **Figure SN7**) further substantiated this observation.

### Supplementary Note 6. Bootstrap evaluation of cancer subtypes' propensity for mtDNA mutations that were predicted to impart benign, pathogenic, or unknown function or occurred within the D-loop.

Cancer subtypes with fewer than 30 samples were excluded from the analysis. Across 10,000 bootstrapped samples, we found the greatest percentage of TU variants predicted to be pathogenic (Figure 1H) among the following tumor subtypes: B-ALL subtype, *E2A–PBX1* translocation (E2A; 5,316 bootstraps), Ewing sarcoma (EWS; 1,504 bootstraps), low-grade glioma (LGG; 1,360 bootstraps), *DUX4*-rearranged (DUX4r; 1,042 bootstraps), Philadelphia chromosome$^{+}$ ALL (PHALL; 209 bootstraps), B-ALL carrying the *ETV6–RUNX1* translocation (ETV; 97 bootstraps), high-grade glioma (HGG; 54 bootstraps), neuroblastoma (NBL; 50 bootstraps), ependymoma (EPD; 11 bootstraps), and hyperdiploid leukemia (HYPER; 10 bootstraps). Thus, we have 92.22% bootstrap confidence that E2A, EWS, LGG, or DUX4r are the cancer subtypes with the greatest percentage of TU variants deemed pathogenic. In addition, the EPD subtype had the greatest percentage of TU variants with an "unknown" role in 9,113 of 10,000 bootstraps.

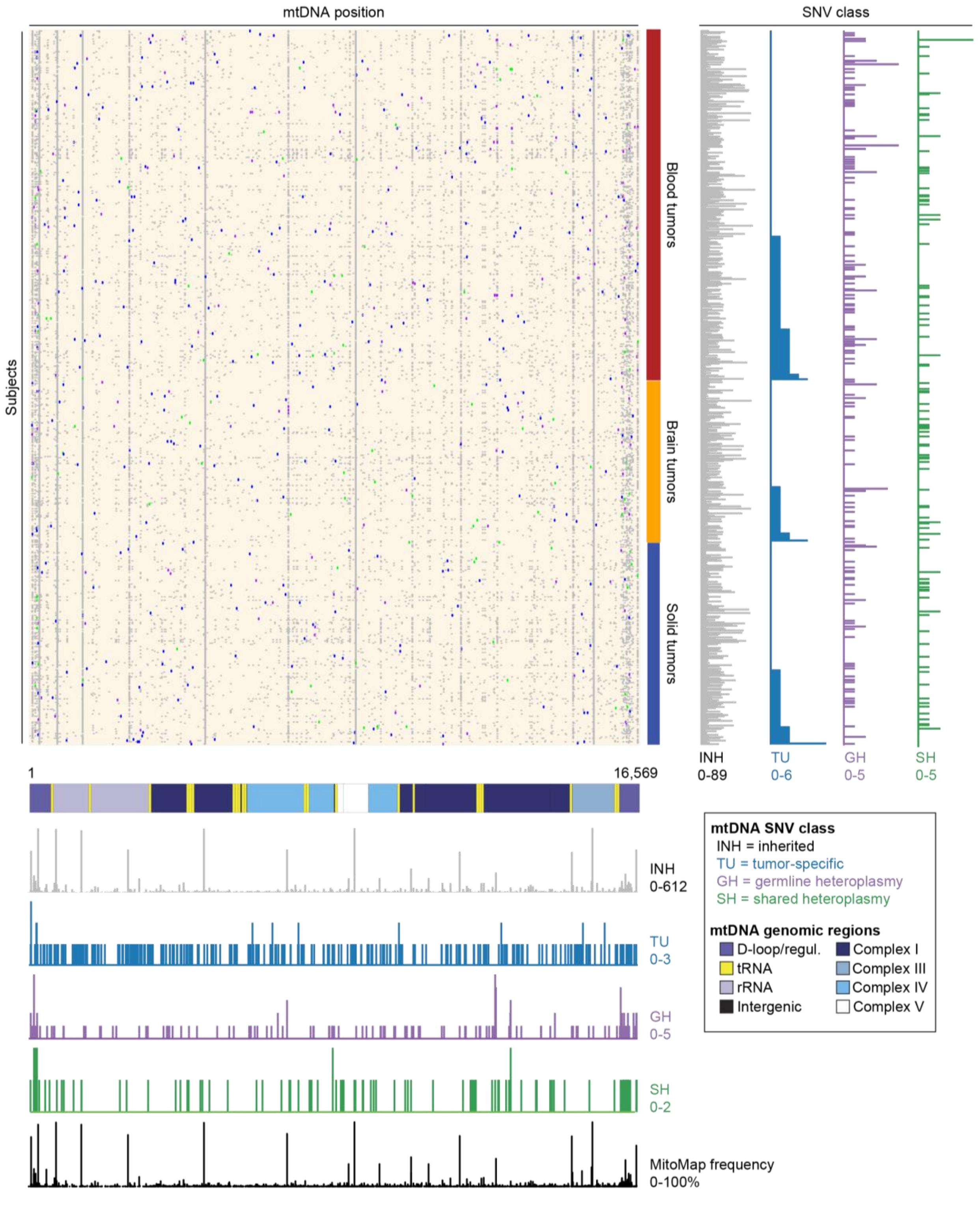


**Figure SN5. Landscape of mtDNA variants in pediatric patients with malignancies.** Heatmap (main panel) representing subject-level enumeration of mtDNA SNVs found among bulk tumors in pediatric patients (PCGP cohort[36]). Each row represents one case, and cases are grouped by tumor type (blood, brain, solid) and ordered by the total number of TU SNVs (vertical, blue histogram; right panel). The mtDNA SNVs are ordered along the x-axis by mtDNA bp positions, 1-16,569 (main panel), corresponding to the mitochondrial genome ideogram below the main panel. Distribution of variants by SNV class are similarly ordered by mtDNA bp position (bottom panel, below ideogram), with histogram height reflecting the variant count at each position. MitoMap[14] population frequencies (black histogram) are provided at the bottom and align with INH class variants (common polymorphisms). **Abbreviations:** bp, base pairs; INH, inherited germline heteroplasmy mtDNA, mitochondrial DNA; PCGP, Pediatric Cancer Genome Project; SNV, single-nucleotide variant TU, tumor-enriched.

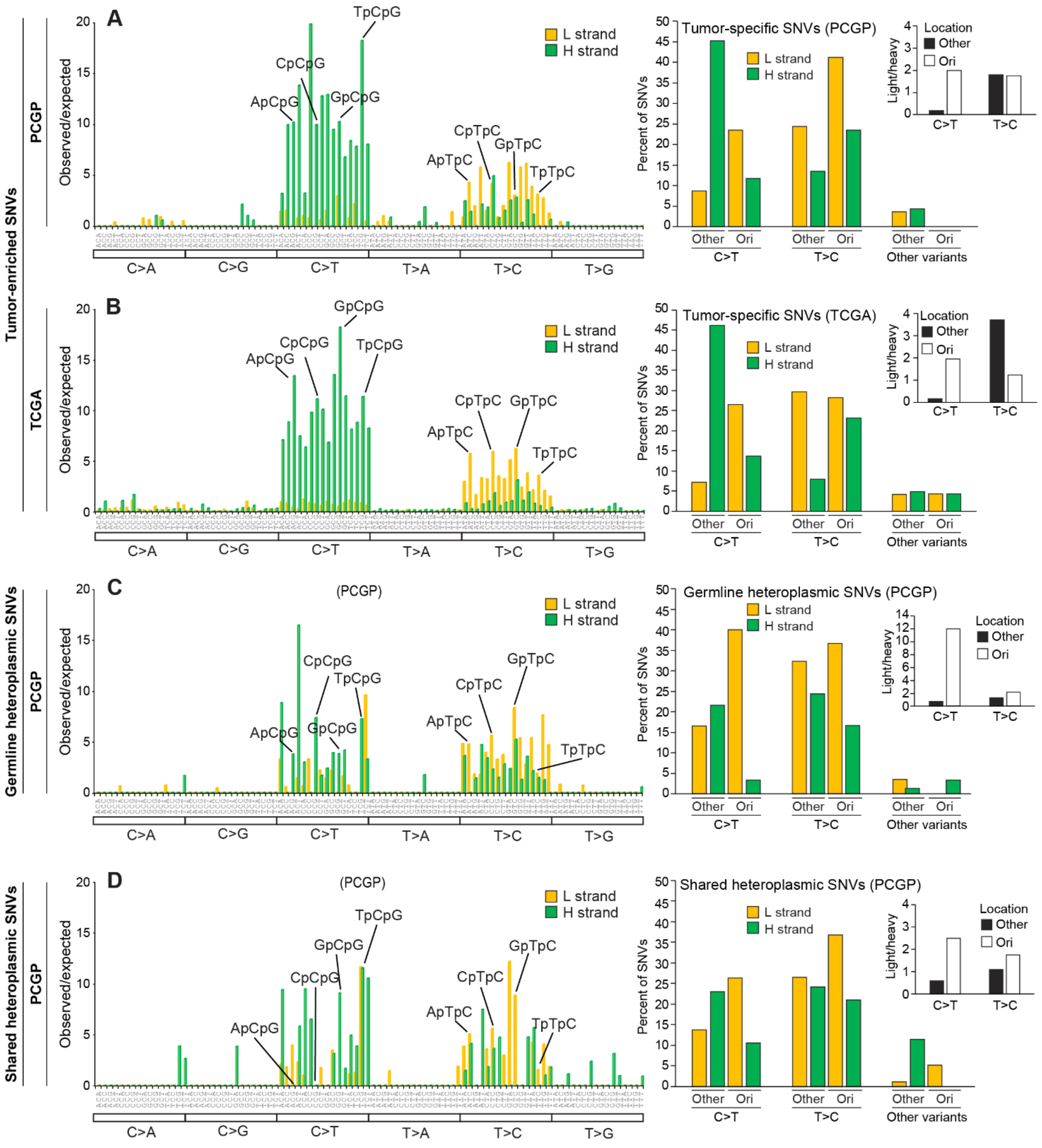


**Figure SN6. Origins of somatic mtDNA mutations.** Histograms (left panels) showing strand-specific trinucleotide sequence composition of the following SNV classes: (A) TU variants within the pediatric cohort (PCGP[36]); (B) TU variants within the adult cohort (TCGA); (C) GH variants within the pediatric cohort (PCGP[36]); (D) SH variants within the pediatric cohort (PCGP[36]). The heights of bars correspond to the ratio of observed *vs.* expected counts, with each of the 96 potential trinucleotide mutational combinations traversing the x-axis. Mutational signature analysis for PCGP *vs.* TCGA TU SNVs: L-strand cosine similarity = 0.887, H-strand = 0.891; for PCGP TU *vs.* PCGP germline: L-strand cosine similarity = 0.757, H-strand = 0.708. Right panel: actual strand-specific mutational counts were binned to highlight the major C>T and T>C substitution classes, and the distribution of these variants within the D-loop/regulatory regions of the mitochondrial genome (between origin of replication regions, $O_H$ through to Ori-b), *vs.* all other regions of the mitochondrial genome. Consistent with the analysis performed by Ju et al.[18], our bar graphs (right panel, inset) showing potential strand inversion are provided. The ratio of L/H strands between the critical D-loop region and the rest of the mitochondrial genome are binned by the major substitution classes. **Abbreviations:** GH, germline heteroplasmy; H, heavy; L, light; $O_H$, origin for H-strand DNA replication; Ori-b, bidirectional initiation origin for mtDNA replication; PCGP, Pediatric Cancer Genome Project; SH, shared heteroplasmy; SNV, single-nucleotide variant; TCGA, The Cancer Genome Atlas; TU, tumor-enriched.

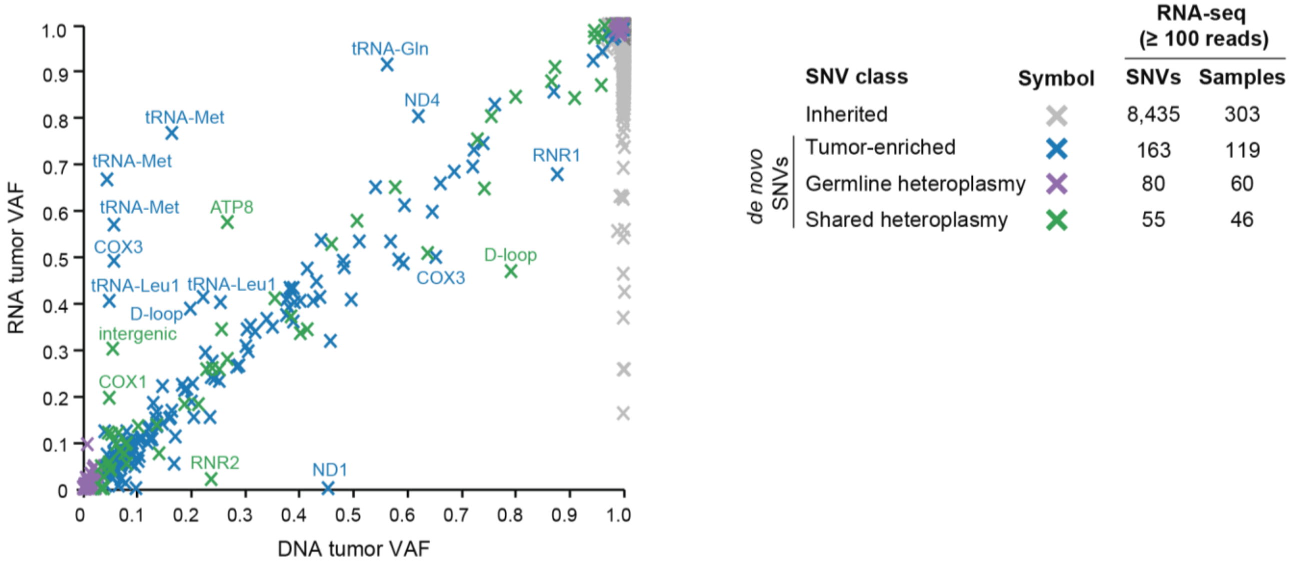


**Figure SN7. Validation of mtDNA variants by RNAseq.** RNAseq validation of 8,732 mitochondrial variants revealed transcriptional infidelity among a fraction of TU (blue) and INH (grey) SNVs. Two-dimensional scatter plot depicts RNAseq-derived VAFs and WGS-derived VAFs. Transcriptional VAFs were notably elevated in TU variants localizing to MT-tRNAs. SNV frequency and sample counts per SNV class are listed. Labeled points indicate TU, GH, or SH loci, where the VAF difference between RNAseq and WGS is >15%. **Abbreviations:** ATP8, ATP synthase F0 subunit 8; COX, cytochrome c oxidase; Gln, glutamate; INH, inherited germline heteroplasmy; Leu, leucine; Met, methionine; MT-tRNA, mitochondrial transfer RNA; ND, NADH dehydrogenase; RNR1, 12S ribosomal RNA; RNR2, 16S ribosomal RNA; SH, shared heteroplasmy; SNV, single nucleotide variant; TU, tumor-enriched; VAF, variant allele fraction; WGS, whole-genome sequencing.

### Supplementary Note 7. Single-cell characterization of SH and GH mutations in *MLL*-rearranged infant leukemia.

The two non-TU *de novo* variants in the *MLL*-rearranged infant leukemia (INF010) sample, a SH m.4674A>G (*MT-ND2*) and a GH m.95A>T (D-loop) substitution, have been noted in large population-based studies[15,24,25]. For the D-loop GH variant (m.95A>T), which was detected by bulk WGS at a VAF of 23.5% in the nontumor sample and a VAF of 1.7% in the tumor sample, only one of 19 sequenced cells from the leukemic sample harbored the variant at an appreciable level (39.3% VAF) (**Figure 2A**). These results indicate that the low tumor VAF originated from the presence of the variant in a very small proportion of cells (likely to be normal hematopoietic cells) in the diagnostic sample. Conversely, the SH variant (m.4674A>G *MT-ND2*), which was detected by bulk WGS at a VAF of 29.6% in the nontumor sample and a VAF of 26.6% in the tumor sample, was present in all 19 cells but at lower VAFs (IQR,5.3%-24.8% VAF) (**Figure 2A**).

### Supplementary Note 8. Functional implications of the p.G142E mutation in MT-CYB (complex III).

To understand the functional impact of the p.G142E (m.15171G>A) mutation on cytochrome b (MT-CYB) activity, we employed an orthogonal approach to computational analysis. Structural analysis showed that the G142 residue resides inside the ligand-binding site of MT-CYB. The endogenous ligand of MT-CYB, ubiquinol, enters the volume comprising residues of the cd1 and ef helices, termed the $Q_o$-binding site, which is distinct from the $Q_i$-binding site that binds quinone[26]. Substitution of the small, neutral glycine residue with the charged, long side chain glutamate alters the physicochemical character of the ligand-binding pocket of MT-CYB (**Figure SN8A**). Molecular Dynamics (MD) simulations of both the wild-type and the p.G142E mutant provided details on the possible mechanism of glutamate substitution. We found that the p.G142E reduced the cavity volume of the ligand-binding site (**Figure SN8B**). Owing to the introduction of the negative charge, the $Q_o$-binding site was also more hydrated because of the additional close-contact interactions with water molecules (**Figure SN8C**).

We next determined whether the reduced cavity volume and reduced hydrophobic character of the binding pocket are important for ligand binding. Unbiased MD simulations of MT-CYB in a membrane containing ubiquinol showed that binding of the latter followed a tail-first approach, whereby the tail of ubiquinol entered the $Q_o$-binding site, thereby stabilizing the ligand and enabling the headgroup to properly orient and presumably insert into the cavity (**Figure SN8D**). This is made possible by the hydrophobic character of the

$Q_o$-binding site, supporting our claim that the p.G142E mutation disfavors ligand entry due to the reduced cavity volume. Because ligand entry followed a tail-first approach, it disallowed the conformational change required for full-ligand entry, owing to the additional cost requirement to displace water.

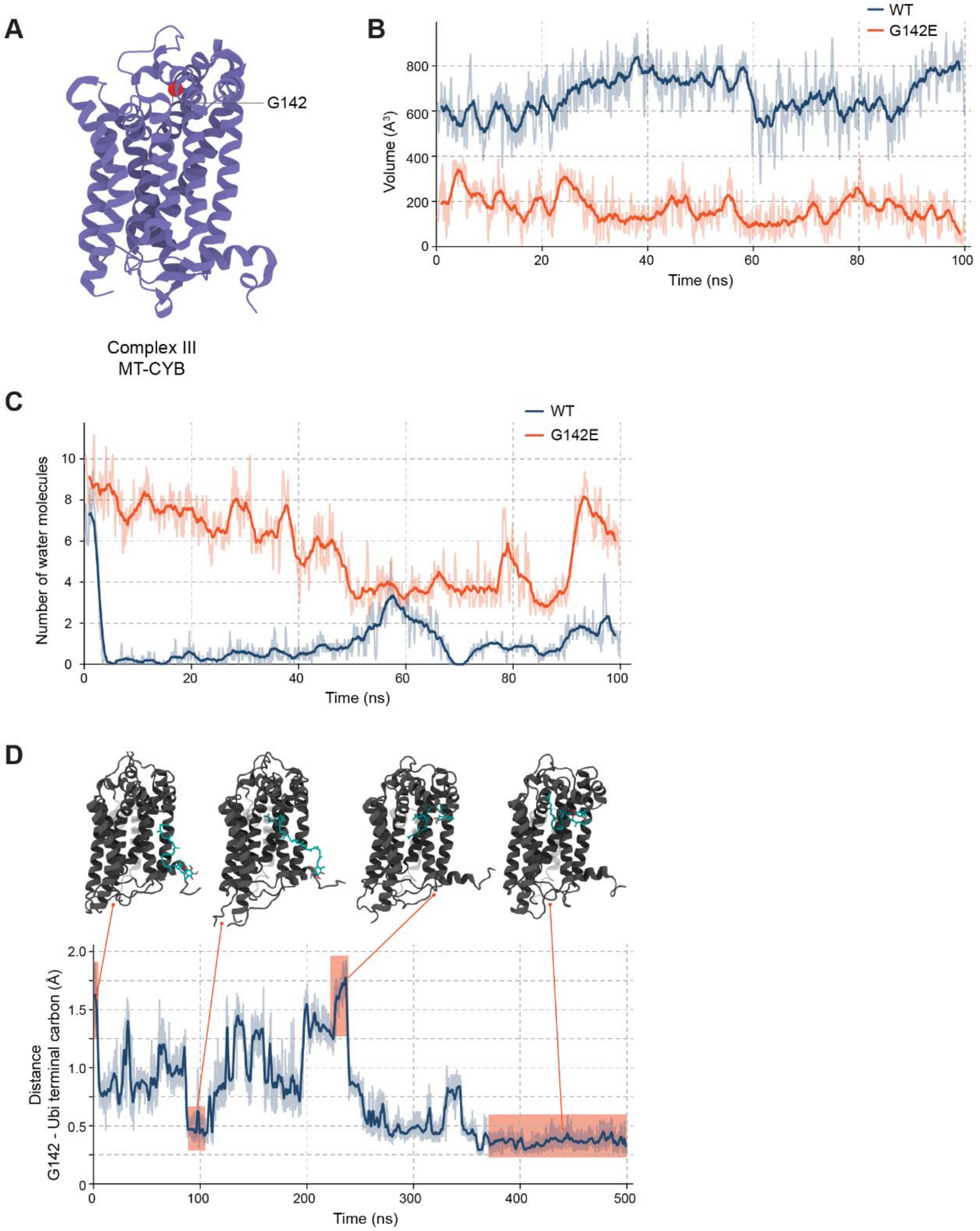


**Figure SN8. Predicted functional consequences of the p.G142E substitution, resulting from the m.15171G>A mutation (ETV027).** (A) Cytochrome *b* (MT-CYB) secondary structure. A red sphere is used to represent the location of G142. (B) Comparison of ligand binding cavity volume between the wild-type (WT) and G142E mutant. The introduction of a negatively charged glutamate residue leads to a noticeable reduction in cavity volume which is stable and maintained throughout the simulation. (C) G142E mutation leads to the hydration of the ligand binding pocket. The number of water molecules that are within 5Å of residue 142 are depicted. (D) Distance measurements between G142 residue and the terminal carbon atom of the ligand acyl chain, showing the ligand probe interactions with cavity residues and the resulting insertion of the ligand tail into the binding site, a position that is stable and maintained for the rest of the trajectory. This configuration likely puts the ligand headgroup in the required position to enter the pocket. **Abbreviations**: ns, nanosecond; $A^3$, $Angstroms^3$ as a unit of volume.

**Supplementary Note 9. Coverage of mtDNA-encoded transcripts by single-cell RNA sequencing and concordance with whole genome sequencing allele frequencies.**

Samples were chosen based on whether their mtDNA variants of interest were within the range of coverage imposed by the assay [i.e., maximal coverage within 300 bp of the poly(A) tail], as detailed in **Figure SN9A-C.**

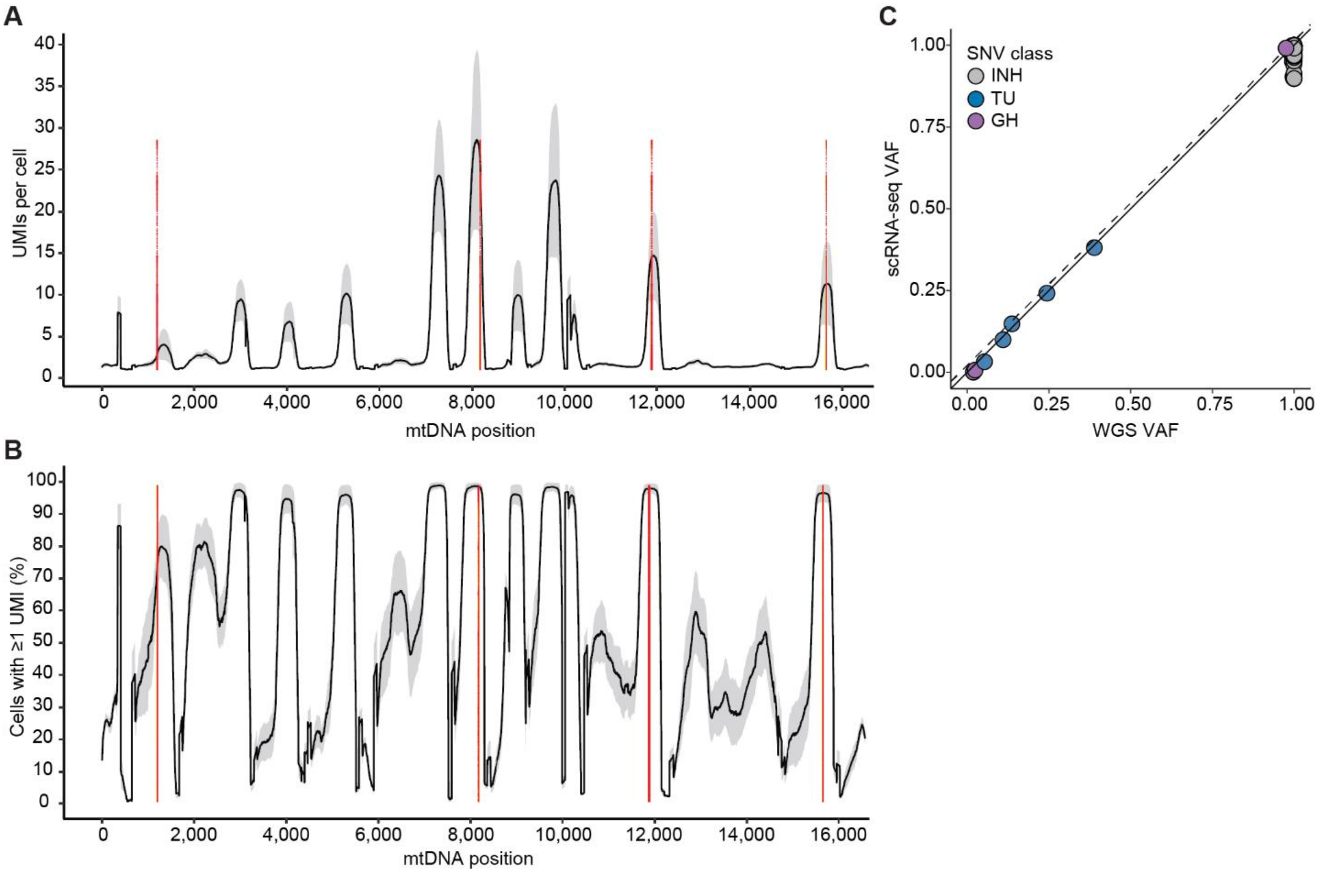


**Figure SN9. Coverage of mtDNA-encoded transcripts by single-cell RNA sequencing and concordance with whole genome sequencing allele frequencies.** (A-B), Line plots indicate the: (A) average number of unique molecular identifiers (UMIs) per cell per position and (B) average percentage of cells with at least one UMI per position using single cell data from three E2A-PBX$^+$ samples. In both plots, UMI base-pair (bp) positions are plotted as they occur along the mitochondrial genome (1-16,569 bp), and red lines indicate the bp position of tumor-enriched variants in the three samples selected for single-cell RNA sequencing characterization. Grey shaded areas reflect the standard deviation. (C) Concordance correlation plot demonstrating high concordance between VAFs from single-cell RNA sequencing (y-axis) and whole-genome sequencing (x-axis) platforms. Lin's concordance correlation coefficient: 0.99 (95% CI 0.99 − 0.99); dashed line represents the line of perfect concordance; solid line represents the reduced major axis. Mitochondrial variants are colored by single nucleotide variant (SNV) class (TU, tumor-enriched = blue; INH, inherited = grey; GH, germline = purple) and represent SNVs detected by scRNA-seq (among all three E2A-PBX cases) that had more than 1 UMI per cell. **Abbreviations**: UMI, unique molecular identifier; bp, base pair; VAF, variant allele fraction.

**Supplementary Note 10. Predicting prednisolone sensitivity by using bulk RNA seq and scRNAseq.**

Given that glucocorticoids are an essential component of chemotherapy for ALL, the initial prednisolone response is a strong prognostic indicator of overall response to therapy[27-30]. Genes that are typically downregulated in ALL in response to glucocorticoid treatment[31] were activated in cells harboring the m.11889G>A variant. Therefore, we examined whether the presence of this mtDNA variant influenced the predicted prednisolone response. Leveraging a discovery cohort of 161 patients with newly diagnosed pediatric B-ALL (St. Jude Total Therapy XV and XVI) for which both *ex vivo* prednisolone sensitivity ($LC_{50}$) and RNAseq data were available[32], we first identified the top 100 significantly upregulated ($P \leq 0.00069$) and the top 100 significantly downregulated ($P \leq 9.68 \times 10^{-6}$) differentially activated drivers that distinguished sensitive ($LC_{50}$ <0.1 µM) from resistant ($LC_{50}$ > 64 µM) samples (**Figure SN10A** and **Table S2Q**). These genes were used to calculate a PSP score for each sample, which successfully discriminated prednisolone-sensitive samples from prednisolone-resistant samples, regardless of protocol (Total XV, $P = 0.00012$ and Total XV1, $P = 4.8 \times 10^{-8}$, Student's *t*-test) (**Figure SN10B**). Subsequent testing against a validation cohort composed of 179 patients with B-ALL (n = 115 pediatric; n = 64 adults), for whom *ex vivo* prednisolone sensitivity values ($LC_{50}$) and gene expression data were available[32], confirmed the utility of our PSP score (**Figure SN10C** and **Table S2R**). This analysis revealed superior separation between sensitive and resistant groups among the pediatric samples ($P = 1.9 \times 10^{-5}$, Student's *t*-test,) as compared to adults ($P = 0.0012$, Student's *t*-test), a finding that was expected, given that the prednisolone-sensitivity biomarkers were developed within a pediatric discovery model and that adult patients are more prone to inferior treatment outcomes[33].

Utilizing these same biomarkers, we next calculated a PSP score for each cell captured by scRNAseq to determine whether there were significant differences between mutant-bearing and WT blasts from each of the three E2A–PBX1$^{+}$ samples analyzed. We found that cells harboring the m.11889G>A variant had significantly lower PSP scores (predicted to be more resistant to prednisolone) than their WT counterparts ($P = 5.6 \times 10^{-5}$) (**Figure SN10D**). In contrast, the presence or absence of the other two mtDNA variants had minimal impact on the PSP score (**Figure SN10D**). The m.11889G>A mutation was predicted to disrupt the function of MT-ND4, a key component of the OXPHOS complex I. Thus, we tested whether prednisolone sensitivity of B-ALL cells would be affected by rotenone, a highly specific complex I inhibitor that engages MT-ND4[34,35].

Rotenone treatment at low doses conferred a small degree of protection against prednisolone-induced cell death (**Figure SN10E**) and decreased the PSP score (**Figure SN10F**). Together, these results suggest that the mitochondrial dysfunction associated with select TU mtDNA mutations has the potential to alter cell state and therapy response, thereby influencing functional heterogeneity among leukemic blasts at the time of diagnosis.

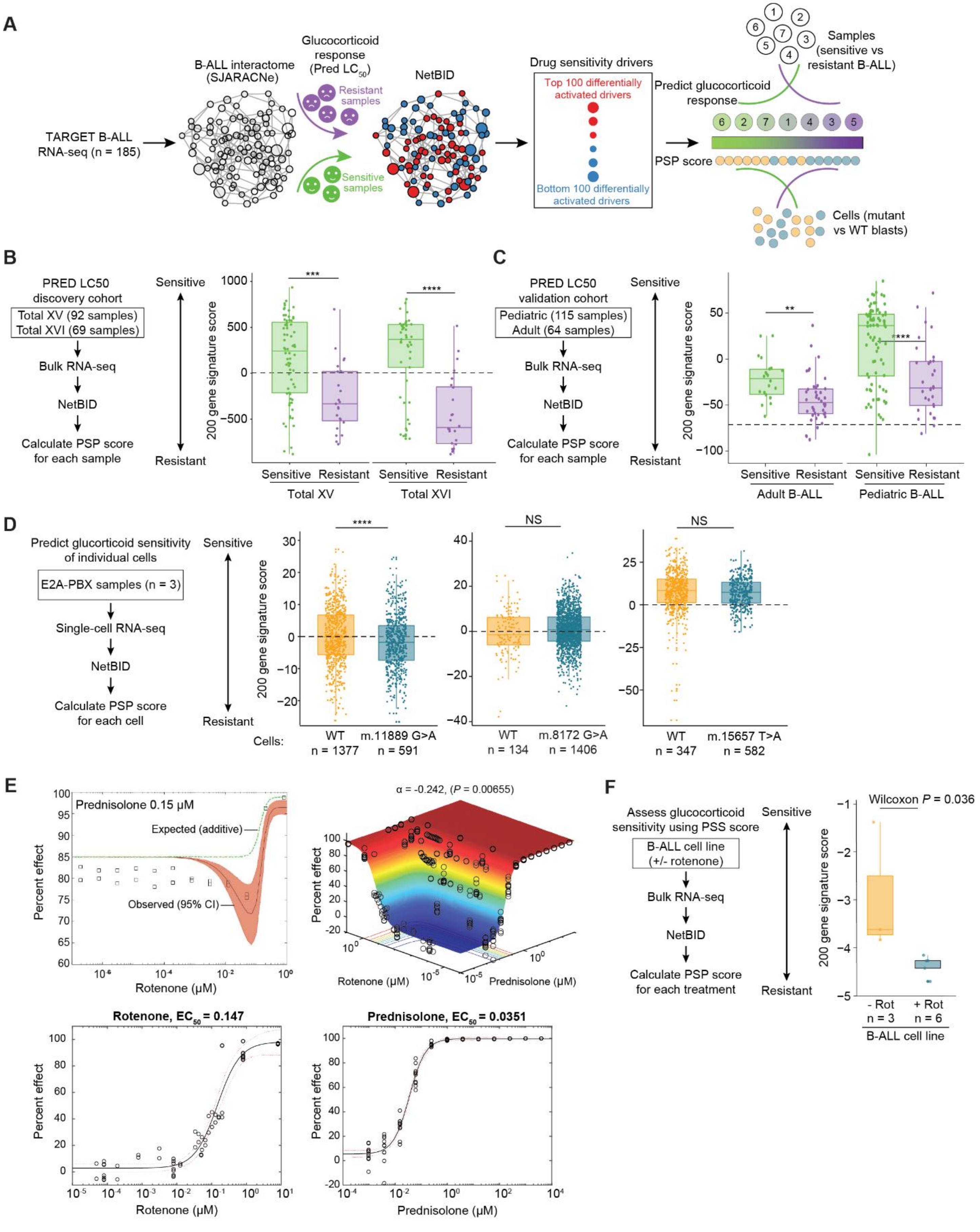


**Figure SN10. Tumor-enriched mtDNA variant influences sensitivity to glucocorticoids.** (A) Schematic for biomarker-based development of prednisolone sensitivity prediction (PSP) score. Using the SJARACNe algorithm, a B-ALL interactome was constructed from an RNAseq dataset comprising 185 pediatric B-ALL samples (TARGET cohort). The reverse-engineered regulatory network was then superimposed onto a NetBID analysis of gene expression and *ex vivo* prednisolone sensitivity ($LC_{50}$) data derived from a pharmacotyping discovery cohort of pediatric patients with newly diagnosed ALL (n = 203, St. Jude Total Therapy XV and XVI). A PSP score was then calculated for each sample by using the top 100 most upregulated ($P$ <0.00069 and top 100 most downregulated ($P <9.68 \times 10^{-6}$) differentially activated drivers that distinguished sensitive ($LC_{50}$ <0.1 µM) from resistant ($LC_{50}$ >64 µM) samples. Each PSP score represents a weighted mean of activities across all 200 drivers. (B) As expected, PSP scores discriminated between prednisolone-sensitive (green) and -resistant (purple) samples in the discovery cohort. Box plots highlighting median differences between resistant and sensitive groups (***$P \leq 0.001$; ****$P \leq 0.0001$; two-sided Student's *t*-test); black dashed line demarcates the null. (C) Analysis of PSP scores in patients from an independent validation cohort revealed superior separation between prednisolone-sensitive and -resistant pediatric samples, as compared to those from adults (**$P \leq 0.01$; ****$P \leq 0.0001$; two-sided Student's *t*-test); black dashed line demarcates the null. (D) Analysis of PSP scores in leukemic cells harboring wild-type (WT) mtDNA or the indicated mutation. Left panel: Boxplots reflect median PSP scores between WT (gold) and mutant-bearing (teal) leukemia cells, revealing that cells harboring the m.11889G>A mutation in *MT-ND4* were predicted to be more resistant to prednisolone than WT cells ($P = 5.6 \times 10^{-5}$, two-sided Wilcoxon signed-rank test) (left panel). When compared to their respective wild-type (i.e. harboring WT mtDNA) leukemia cells, no significant differences in PSP scores were detected between leukemia cells harboring either the m.15657T>A (middle panel) or the m.8172G>A (right panel) mutations. Black dashed lines demarcate the null. (E) Treatment of a B-ALL cell line (RS;411) with the complex I inhibitor rotenone recapitulates the prednisolone resistance associated with the m.11889G>A mutation. Increasing micromolar concentrations of rotenone confer a small degree of protection against cell death (percent effect = percent killed) in the presence of 0.15 µM prednisolone (upper left panel). Response surface modeling[39,40] (upper right panel), which provides a more comprehensive view of the combinatorial effects of drug treatment, indicates that rotenone has a small but significant antagonistic effect on prednisolone. Dose-response curves (lower panels) show individual effects of each agent. (F) Rotenone-induced resistance to prednisolone is predicted to occur in the RS;411 cell line, where rotenone-treated cells exhibit significantly lower PSP scores than do untreated cells ($P = 0.036$, two-sided Wilcoxon signed-rank test).

### Supplementary Note Tables

| Tumor Subtype | Tier 1[b] Coefficient | Tier 1 *P*-value | Tier 2[c] Coefficient | Tier 2 *P*-value | Tier 3[d] Coefficient | Tier 3 *P*-value |
|---|---|---|---|---|---|---|
| ACT | 0.16 | 0.5 | 0.18 | 0.44 | 0.18 | 0.44 |
| CBF | 0.15 | 0.69 | 0.43 | 0.22 | 0.36 | 0.31 |
| E2A | -0.17 | 0.48 | -0.21 | 0.37 | 0.15 | 0.54 |
| EPD | 0.25 | 0.13 | 0.09 | 0.57 | 0.15 | 0.37 |
| DUX4r | **0.43** | **0.02** | **0.42** | **0.02** | **0.48** | **0.01** |
| ETV | 0.22 | 0.12 | 0.18 | 0.2 | 0.2 | 0.16 |
| EWS | **0.53** | **0** | 0.3 | 0.08 | **0.38** | **0.03** |
| HGG | 0.27 | 0.12 | **0.35** | **0.04** | 0.26 | 0.13 |
| HYPER | -0.04 | 0.79 | 0.12 | 0.46 | 0.02 | 0.91 |
| HYPO | 0.4 | 0.09 | -0.03 | 0.91 | 0.09 | 0.72 |
| INF | -0.06 | 0.84 | 0.11 | 0.72 | 0.19 | 0.54 |
| LGG | 0.03 | 0.87 | 0.32 | 0.05 | 0.22 | 0.18 |
| MB | -0.11 | 0.63 | -0.21 | 0.34 | -0.23 | 0.31 |
| NBL | **0.28** | **0.04** | **0.31** | **0.02** | **0.28** | **0.04** |
| OS | 0.14 | 0.43 | 0.24 | 0.18 | 0.27 | 0.14 |
| PHL | 0.31 | 0.16 | 0.13 | 0.55 | 0.13 | 0.56 |
| PHL-Hyp | 0.55 | 0.1 | 0.11 | 0.76 | 0.11 | 0.76 |
| PH | **0.36** | **0.03** | 0.28 | 0.09 | 0.33 | 0.05 |
| RB | 0.46 | 0.15 | 0.27 | 0.42 | 0.46 | 0.16 |
| RHB | 0.41 | 0.27 | 0.41 | 0.27 | 0.41 | 0.27 |
| TALL | 0.1 | 0.75 | -0.07 | 0.83 | -0.07 | 0.83 |

**Table SN1**. **Correlations between mtDNA SNVs and annotated nuclear SNVs.[a]**

[a]List of samples with nuclear SNV data available. Subtypes with fewer than six samples were excluded from the analysis.
[b]Tier 1 includes coding synonymous, nonsynonymous, splice-site, and noncoding RNA variants.
[c]Tier 2 includes regulatory regions.
[d]Tier 3 includes intergenic and intronic regions.

**Abbreviations:** ACT, adrenocortical carcinoma; CBF, core-binding factor; DUX4r, *DUX4*-rearranged; E2A, *E2A–PBX1* translocation; EPD, ependymoma; ETV, *ETV6–RUNX1* translocation; EWS, Ewing sarcoma; HGG, high-grade glioma; HYPER, hyperdiploid; HYPO, hypodiploid; INF, infant ALL with/without *MLL* rearrangements; LGG, low-grade glioma; MB, medulloblastoma; NBL, neuroblastoma; OS, osteosarcoma; PHL, Philadelphia chromosome–like; PHL-Hyp, Philadelphia chromosome–like with either hyperdiploid or hypodiploid features; RB, retinoblastoma; RHB, rhabdomyosarcoma; TALL, T-cell acute lymphoblastic leukemia.

| SNV type | Predicted Impact | Location | Gene | BP | BP change | AA variant | Mutation Assessor score | Tumor Group | Tumor Subtype | Sample |
|---|---|---|---|---|---|---|---|---|---|---|
| SNV | D-loop | D-loop | noncoding | 72 | T>C | N/A | N/A | Blood | HYPER | SJHYPER007 |
| SNV | D-loop | D-loop | noncoding | 72 | T>C | N/A | N/A | Blood | INF | SJINF010 |
| SNV | D-loop | D-loop | noncoding | 72 | T>C | N/A | N/A | Brain | MB | SJMB029 |
| SNV | D-loop | D-loop | noncoding | 214 | A>G | N/A | N/A | Solid | NBL | SJNBL015 |
| SNV | D-loop | D-loop | noncoding | 214 | A>G | N/A | N/A | Solid | EWS | SJEWS010367 |
| SNV | D-loop | D-loop | noncoding | 228 | G>A | N/A | N/A | Blood | TALL | SJTALL006 |
| SNV | D-loop | D-loop | noncoding | 228 | G>A | N/A | N/A | Blood | PH | SJPHALL020044 |
| Stop-gain | Pathogenic | Complex I | *ND1* | 3842 | G>A | W179X | N/A | Blood | HYPO | SJHYPO026 |
| Stop-gain | Pathogenic | Complex I | *ND1* | 4156 | C>T | Q284X | N/A | Solid | MEL | SJMEL001002 |
| Missense | Pathogenic | Complex I | *ND2* | 5094 | A>G | I209V | 1.97 | Blood | HYPER | SJHYPER023 |
| Missense | Pathogenic | Complex I | *ND2* | 5095 | T>C | I209T | 2.09 | Blood | DUX4r | SJBALL020340 |
| Missense | Pathogenic | Complex IV | *COX1* | 6087 | G>A | A62T | 3.89 | Blood | HYPER | SJHYPER091 |
| Missense | Pathogenic | Complex IV | *COX1* | 6087 | G>A | A62T | 3.89 | Solid | RHB | SJRHB013 |
| Missense | Pathogenic | Complex IV | *COX1* | 6631 | T>C | V243A | 5.72 | Solid | ACT | SJACT005 |
| Missense | Pathogenic | Complex IV | *COX1* | 6631 | T>C | V243A | 5.72 | Blood | ETV | SJETV078 |
| Stop-gain | Pathogenic | Complex IV | *COX1* | 6708 | G>A | G269X | N/A | Blood | E2A | SJE2A059 |
| Missense | Benign | Complex IV | *COX1* | 6891 | A>G | S330G | 0.03 | Brain | LGG | SJLGG019 |
| Missense | Pathogenic | Complex IV | *COX1* | 6892 | G>A | S330N | 3.07 | Solid | NBL | SJNBL032 |
| Missense | Pathogenic | Complex IV | *COX1* | 7125 | C>T | T408A | 1.18 | Blood | PH | SJPHALL003 |
| Missense | Pathogenic | Complex IV | *COX1* | 7126 | A>G | T408I | 1.87 | Brain | HGG | SJHGG007 |
| Silent | Benign | Complex IV | *COX1* | 7337 | G>A | S478S | N/A | Solid | NBL | SJNBL187 |
| Silent | Benign | Complex IV | *COX1* | 7337 | G>A | S478S | N/A | Brain | HGG | SJHGG101 |
| Missense | Pathogenic | Complex IV | *COX1* | 7697 | G>A | V38I | 1.58 | Blood | PHL-Hyp | SJHYPER013 |
| Silent | Benign | Complex IV | *COX1* | 7699 | C>A | V38V | N/A | Solid | OS | SJOS001107 |
| Stop-gain | Pathogenic | Complex IV | *COX3* | 9450 | G>A | G82X | N/A | Solid | NBL | SJNBL003 |
| Missense | Benign | Complex I | *ND3* | 10068 | G>A | A4T | -0.83 | Solid | OS | SJOS004 |
| Missense | Benign | Complex I | *ND3* | 10068 | G>A | A4T | -0.83 | Solid | OS | SJOS009 |
| Stop-gain | Pathogenic | Complex I | *ND4* | 10806 | G>A | W16X | N/A | Solid | EWS | SJEWS001304 |
| Stop-gain | Pathogenic | Complex I | *ND5* | 12441 | T>A | Y35X | N/A | Solid | ACT | SJACT001 |
| Missense | Pathogenic | Complex I | *ND5* | 12851 | T>C | I172T | 3.15 | Solid | EWS | SJEWS010362 |
| Missense | Pathogenic | Complex I | *ND5* | 12851 | T>C | I172T | 3.15 | Blood | HYPO | SJHYPO056 |
| Stop-gain | Pathogenic | Complex I | *ND5* | 13414 | G>A | G360X | N/A | Blood | HYPER | SJHYPER020 |
| Stop-gain | Pathogenic | Complex I | *ND5* | 13556 | G>A | W407X | N/A | Solid | ACT | SJACT005 |
| Silent | Benign | Complex I | *ND5* | 13557 | A>G | W407W | N/A | Blood | CBF | SJCBF009 |
| Stop-gain | Pathogenic | Complex III | *CYTB* | 14888 | G>A | G48X | N/A | Solid | OS | SJOS011 |
| Silent | Benign | Complex III | *CYTB* | 15067 | T>C | F107F | N/A | Solid | RB | SJRB031 |
| Silent | Benign | Complex III | *CYTB* | 15067 | T>C | F107F | N/A | Blood | TALL | SJTALL013 |
| Stop-gain | Pathogenic | Complex III | *CYTB* | 15234 | G>A | W163X | N/A | Solid | OS | SJOS004 |
| Stop-gain | Pathogenic | Complex III | *CYTB* | 15240 | G>A | W165X | N/A | Solid | NBL | SJNBL031 |
| Missense | Pathogenic | Complex III | *CYTB* | 15656 | A>T | I304F | 1.13 | Blood | CBF | SJCBF012 |
| Missense | Pathogenic | Complex III | *CYTB* | 15657 | T>A | I304N | 3.55 | Blood | E2A | SJE2A025 |
| Missense | Pathogenic | Complex III | *CYTB* | 15657 | T>C | I304T | 2.05 | Blood | INF | SJINF019 |

**Table SN2**. **Recurrent and truncating mtDNA TU SNVs in the PCGP cohort.**

**Abbreviations:** AA, amino acid; ACT, adrenocortical carcinoma; BP, base pair; CBF, core-binding factor; DUX4r, *DUX4*-rearranged; E2A, *E2A–PBX1* translocation; ETV, *ETV6–RUNX1* translocation; EWS Ewing sarcoma; HGG, high-grade glioma; HYPER, hyperdiploid; HYPO, hypodiploid; INF, infant ALL with/without *MLL* rearrangements; LGG, low-grade glioma; MB, medulloblastoma; MEL, melanoma; N/A, not available; NLB, neuroblastoma; OS, osteosarcoma; PH, Philadelphia chromosome; PHL-Hyp, Philadelphia chromosome–like with either hyperdiploid or hypodiploid features; RB, retinoblastoma; RHB, rhabdomyosarcoma; SNV, single-nucleotide variant; TALL, T-cell acute lymphoblastic leukemia; TU, tumor-enriched.
